# Estimating the Financial Impact of Gene Therapy*

**DOI:** 10.1101/2020.10.27.20220871

**Authors:** Chi Heem Wong, Dexin Li, Nina Wang, Jonathan Gruber, Rena Conti, Andrew W. Lo

## Abstract

We assess the potential financial impact of future gene therapies by identifying the 109 late-stage gene therapy clinical trials currently underway, estimating the prevalence and incidence of their corresponding diseases, developing novel mathematical models of the increase in quality-adjusted life years for each approved gene therapy, and simulating the launch prices and the expected spending of these therapies over a 15-year time horizon. The results of our simulation suggest that an expected total of 1.09 million patients will be treated by gene therapy from January 2020 to December 2034. The expected peak annual spending on these therapies is $25.3 billion, and the total spending from January 2020 to December 2034 is $306 billion. We decompose their annual estimated spending by treated age group as a proxy for U.S. insurance type, and consider the tradeoffs of various methods of payment for these therapies to ensure patient access to their expected benefits.

## 1 Introduction

Gene therapy is a new class of medical treatment that alters part of a patient’s genome through the replacement, deletion, or insertion of genetic material to treat a disease. While still in its infancy, gene therapy has demonstrated immense potential to treat and even cure previously intractable diseases. The introduction of voretigene neparvovec (marketed as Luxturna^®^) for inherited retinal disease and onasemnogene abeparvovec-xioi (marketed as Zolgensma^®^) for spinal muscular atrophy (SMA) in the U.S. have improved the lives of patients [115, 124]. Yet the price per treatment of $425,000 per eye for Luxturna, and $2.1 million per patient for Zolgensma, have raised concerns regarding affordability among budget-constrained payers and patients alike.

Stakeholders have expressed concerns that gene therapy will be too expensive for individual patients to afford on their own, especially if they continue to be priced at more than 30 times the median household income of $61,937 [104], (equivalently, at several multiples of the average U.S. home mortgage of $354,400 [126]). Insurance coverage for gene therapy also varies by policy type. Many insurance policies do not cover access to gene therapy, or they impose very restrictive policies to limit the number of patients who might be treated [78, 156]. Widespread underinsurance in the U.S. [117]—requiring substantial out-of-pocket costs in the form of deductibles and coinsurance payments—may place gene therapy out of reach for American patients who might benefit from treatment. Many health plans, especially those facing fixed annual budgets, including state Medicaid policies, some employer-offered plans, and insurance offered on the federal and state-sponsored exchanges, have warned they may not be able or willing to absorb the additional spending should a greater number of people become eligible for expensive gene therapy once it reaches the market [159]. While some spending on novel gene therapies would likely be paid for by Medicare, the taxpayer-supported health insurance for Americans over the age of 65, other spending on these treatments might have an impact on the wages that private corporations pay to their employees [70].

In this paper, we estimate the potential fiscal impact of gene therapy on the U.S. market. To do so, we create a new model to estimate the future number of gene therapy approvals, the size of their potential patient populations, and the prices of these future treatments. We begin by surveying the clinical trial databases for late-stage gene therapy trials, defined here as phase 2/3 or phase 3, and compile the prevalence and incidence of the diseases targeted in these trials from a meta-analysis of published sources. We develop a novel method to estimate the price of each gene therapy under consideration by calculating the expected quality-adjusted life years gained for each therapy in the relevant patient population. Combining these results and previously published probabilities of technical success by therapeutic area, we simulate whether a disease will have an approved gene therapy over the next fifteen years, the expected number of treated patients and the expected spending from January 2020 to December 2034. Throughout this paper, the expected number of treated patients refers to the pool of eligible patients that remains after accounting for market penetration and the treatment schedule, explained in more detail in Section 2.4.

Our results suggest that an estimated total of 1.09 million patients will be treated with gene therapy by the end of December 2034. The number of patients receiving gene therapies annually will peak at 94,696 patients in 2025 before declining to 65,612 at the end of our time period. The annual spending across all expected products and patients is expected to reach $25.3 billion in 2026. The cumulative spending on treating these patients in the 15-year period is estimated to be $306 billion, or $241 billion when discounted by 3% per annum to 2020 dollars. We decompose the expected spending by patient age group as a proxy for insurance type. For the expected spending on these therapies that is not covered by Medicare, we consider several alternate ways of financing.

## 2 Simulation Design

A critical element in our simulation analysis is the number of gene therapies that will receive regulatory approval over the next few years. Therefore, we begin with a brief review of the approval process in the U.S.

Since the passing of the Food, Drug, and Cosmetic Act in 1938, pharmaceuticals developed by companies have to be reviewed by the Food and Drug Administration (FDA) for safety and efficacy before they can be marketed in the U.S. The application for marketing approval differs slightly by the type of therapy: New Drug Applications (NDAs) are for small molecules, and Biologics License Applications (BLAs) are for biologics. Gene therapy is considered a biologic product, hence the BLA designation applies.

Clinical investigations in human subjects typically take place in three phases—phases 1, 2 and 3—before marketing approvals are sought. Phase 1 trials are designed to investigate the dosage and safety of the treatment, while phase 2 trials attempt to detect early signs of efficacy and possible side effects in a relatively small sample of patients. Phase 3 trials are intended to demonstrate a statistically significant treatment effect when compared to the best standard of care in a broader population of patients. Some clinical trials combine multiple phases into a single design, with the phase numbers separated by a slash. For example, a phase 2/3 trial combines elements of phase 2 and phase 3 investigations into a single trial design in order to reduce the overall development time and cost, and maximize the participation of subjects with orphan disease willing to participate in trials. The clinical development of therapeutics is a tedious and costly process that may span decades and cost billions of dollars, with the bulk of the cost and time spent conducting phase 3 clinical trials [63, 84]. The process is also very risky, with only 13.8% of therapeutic development programs entering phase 1 reaching approval [166].

To estimate the financial impact of gene therapies on the U.S. healthcare system, we first identify all existing late-stage clinical trials of gene therapies, simulate their successes or failures from phase 2/3 or 3 to approval, then estimate the spending on the successful ones by summing the product of their expected prices and number of patients, as outlined in Figure 1. By using simulation analysis rather than purely deterministic methods, we are able to capture the inherent uncertainty in costs, revenues, and other parameters of this new therapeutic class.

**Figure 1:**
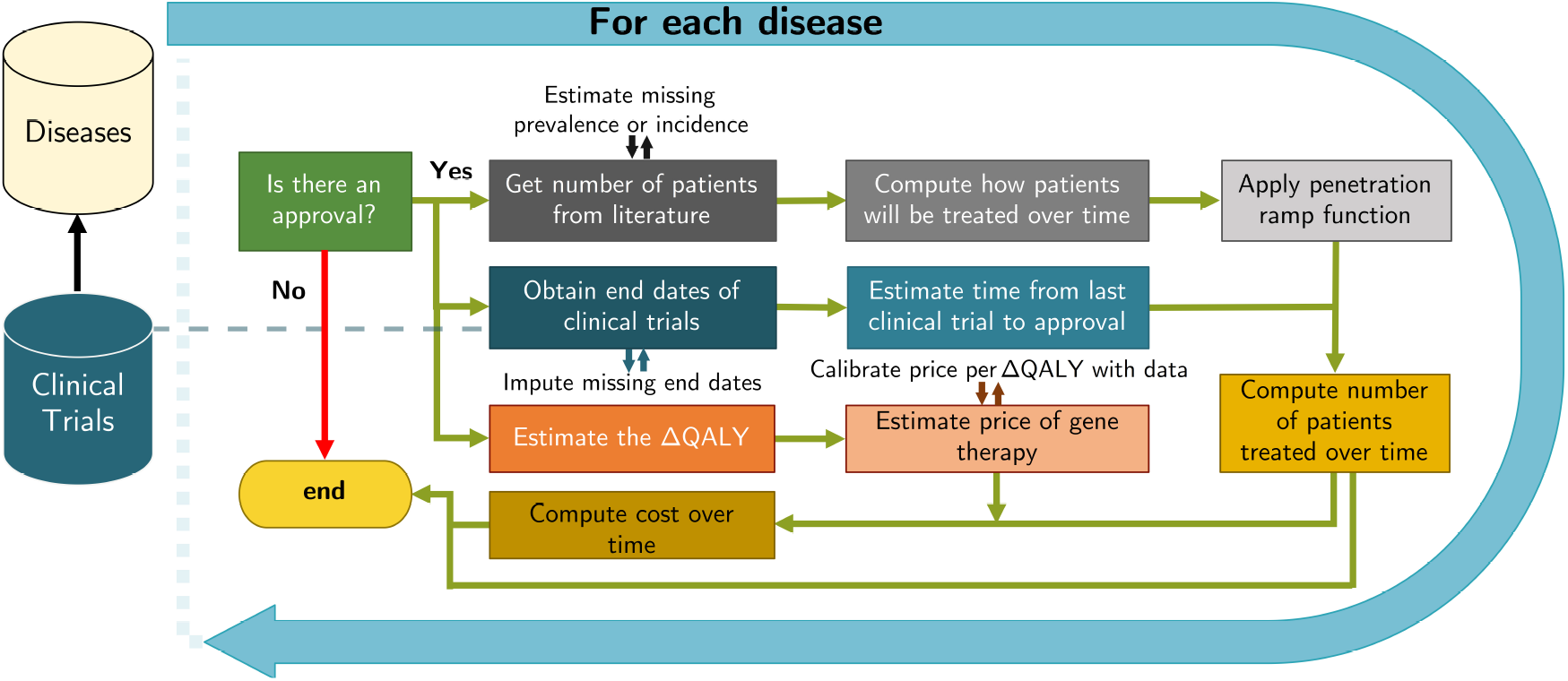
A flowchart showing the performance of the simulation. After extracting the information on each disease from the clinical trial databases, we simulate whether the disease will obtain an approval. If it fails to do so, the simulation will end for this disease in this iteration. Otherwise, we will estimate the expected number of patients to be treated, compute the corresponding cost of treatment, and store the results. At each step of the computation, we source data from literature and impute missing information.

We organize the simulation into the following five distinct modules, and describe each of these in some detail: (1) identifying the number of gene therapies currently in clinical trials; (2) estimating the probabilities of success of these trials; (3) estimating the time to approval; (4) simulating the expected number of patients treated by these therapies if approved; and (5) estimating the expected market prices of the approved therapies. We describe the first four modules in Sections 2.1–2.4. Given the importance and potential controversies surrounding the pricing of gene therapies, we devote a standalone section to this issue in Section 3.

### 2.1 Clinical Trial Data

We use clinical trial metadata from the Citeline TrialTrove database and the U.S. National Library of Medicine’s ClinicalTrials.gov database to determine the number of gene therapies currently under development and their potential number of patients.

We download data from the Citeline database, isolating trials tagged with ‘gene therapy’ under the ‘therapeutic class’ field. We supplement this information by searching for trials on the *clinicaltrials*.*gov* main page using the key words ‘gene therapy’, then reading the trial description to determine if the trial is in fact related to a gene therapy. All database queries were made before May 31, 2019. Clinical trials from both sources are merged before filtering for clinical trials that are in either phase 2/3 or phase 3 of the development process and are not known to be compassionate uses of the treatment.^1^ The outcomes of compassionate use are rarely used as data points in the clinical development process. Even though adverse events from compassionate use are reported to the FDA and may, in rare cases, be used to characterize the risk and benefits of a therapy, the FDA is cognizant that these uses often occur outside of clinical trial settings, and has almost never given an unfavorable decision to a product labeling because of an adverse outcome of compassionate use [88, 116]. We include clinical trials without U.S. trial site in our dataset because it is currently possible for the FDA, as empowered by Federal administrative law 21 CFR Part 312.120, to grant marketing approval using evidence from foreign clinical trials [68].

Our filtering criteria are intended to remove trial entries unrelated to the clinical development process, and to isolate gene therapies that are most likely to seek regulatory approval in the U.S. in the near future.

We remove repeated entries, and identify the diseases and therapeutic areas targeted by each gene therapy. Each clinical trial entry in our dataset contains a brief title of the trial, its clinical phase, the disease being targeted, the start and end dates of the clinical trial, the therapy name, and the companies involved in the clinical trial.

This process yields 109 trials investigating 57 distinct diseases, listed in Table A1 in Supplementary Materials. We classify the diseases into three categories: cancer (oncology),^2^ rare disease, and general disease. The distribution of disease and the clinical trials by category and therapeutic area are shown in Table 1 and Table 2. The majority of trials and diseases are in the area of oncology, followed by rare diseases. These therapeutic areas are notoriously risky. Only 3.1% and 6.2% of the drug development programs in oncology and rare diseases go from Phase 1 to approval, respectively, compared to the baseline of 13.8% across all drugs and indications [166].

**Table 1:** Count of number of clinical trials by category and therapeutic area.

| Therapeutic Area | Cancer | General | Rare Disease | Subtotal |
| --- | --- | --- | --- | --- |
| Autoimmune/Inflammation |  | 3 | 2 | 5 |
| Cardiovascular | - | 15 | 1 | 16 |
| CNS | - | 3 | 7 | 10 |
| Metabolic/Endocrinology | - | 3 | 15 | 18 |
| Oncology | 52 | - | 1 | 53 |
| Ophthalmology | - | - | 7 | 7 |
| Subtotal | 52 | 24 | 33 | 109 |

**Table 2:**
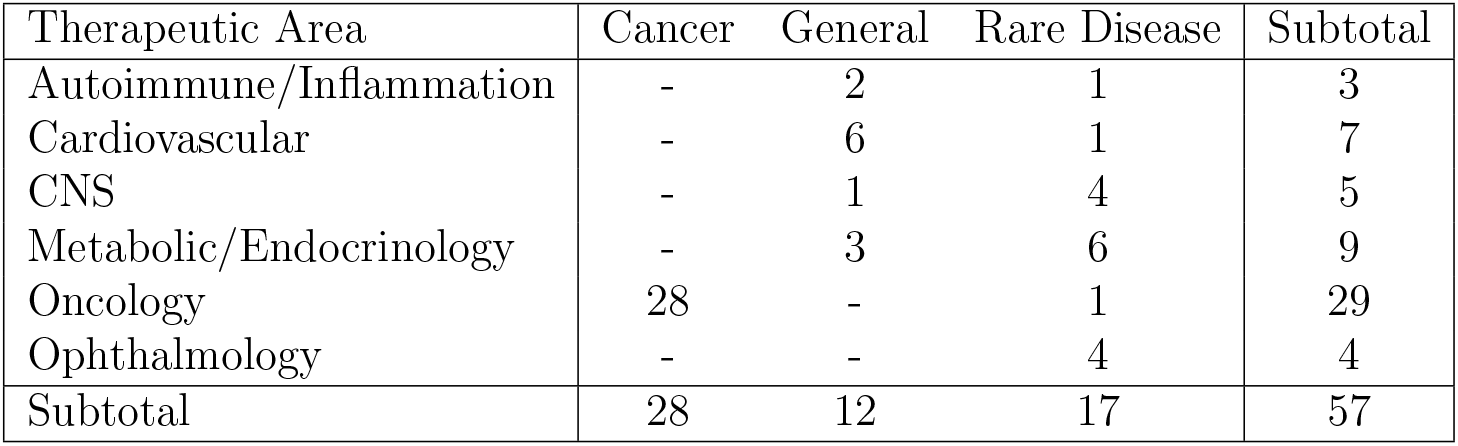
Count of number of diseases by category and therapeutic area.

| Therapeutic Area | Cancer | General | Rare Disease | Subtotal |
| --- | --- | --- | --- | --- |
| Autoimmune/Inflammation | - | 2 | 1 | 3 |
| Cardiovascular | - | 6 | 1 | 7 |
| CNS | - | 1 | 4 | 5 |
| Metabolic/Endocrinology | - | 3 | 6 | 9 |
| Oncology | 28 | - | 1 | 29 |
| Ophthalmology | - | - | 4 | 4 |
| Subtotal | 28 | 12 | 17 | 57 |

### 2.2 Probability of Success Estimates

We define a gene therapy development program as the set of clinical trials by a sponsor testing a therapeutic for a disease. We consider whether a gene therapy will be developed for a disease by simulating correlated coin flips’ for each gene therapy program, and observing if there is at least one approval.

Our computational method assumes that clinical trials are always perfectly correlated within the same development program. This is logical, since the FDA requires findings from at least two pivotal trials in the BLA review process[171]^3^. Our assumption can also be justified for clinical trials run by the same sponsor that target the same disease, but for different patient segments. We reason that the sponsors are risking multiple expensive late-stage trials for the same disease, thus have confidence that the treatment will work on all patient sub-populations, and therefore any marketing licensing approval (or denial) will be similar for all the patient segments.

It can be argued that different gene therapy treatments for a disease are highly correlated, since they operate on similar platforms (e.g. CAR-T or in-vivo gene delivery using adeno-associated virus vectors), even though different gene sequences may be targeted. To reflect this, we consider a correlation of 90% between development programs in our simulations. Our sensitivity analysis, however, demonstrates that these computations are insensitive to this parameter (see Section 4.3).

The phase 3 to approval probability of success (PoS_3*A*_) for each disease is informed by prior studies on the probabilities of success of drug development programs by therapeutic area from the MIT Laboratory of Financial Engineering’s Project ALPHA website [97]. These estimates for the probabilities of success are derived from over 55,000 drug development programs between January 2000 and January 2020, and computed using the path-by-path methods as introduced in Wong et al. [166]. The PoS_3*A*_ values used in our simulations are given in Table 3 and the mapping of diseases to therapeutic areas is shown in Table A2.

**Table 3:** The probability of success of drug development programs from phase 3 to approval (PoS_3*A*_), categorized by therapeutic area. We assume that the probability of success for gene therapy follows a similar pattern.

| Therapeutic Area | $\text{PoS}_{3A}$ (%) |
| --- | --- |
| Autoimmune/Inflammation | 48.5 |
| Cardiovascular | 50.1 |
| Central Nervous System (CNS) | 37.0 |
| Metabolic/Endocrinology | 45.7 |
| Oncology | 28.5 |
| Ophthalmology | 45.9 |

### 2.3 Time to Approval

We also require an estimate of the time to approval for gene therapy treatments in order to assess the patient impact and cost over time. Typically, companies submit their Biologics License Application (BLA) to the U.S. Food and Drug Administration (FDA) some time after the end of the clinical trial period. We assume that the time between the end of the last clinical trial for the disease and the submission of the BLA is a variable drawn from a triangular distribution between 0 and 365 days, with a median of 182.5 days. This is informed by the practical knowledge that it takes an average of 6 months to prepare the documents for the BLA submission [166].

In addition, there will be a lag time between the submission of the BLA and the decision of the FDA. The FDA has 60 days to decide if it will follow up on a BLA filing [91], and it can take another 10 months to deliver its verdict [90]. This implies the maximum possible time between BLA submission and FDA approval will be 12 months. We thus assume that the time between the BLA submission and the FDA decision is drawn from a triangular distribution between 0 and 365 days, with a median of 182.5 days. Our assumptions are also valid for therapies that use the priority review pathways.

We also assume that the BLA will be filed only after the last clinical trial for a disease has ended. Trials with missing declared end dates will have their end dates imputed by adding random durations to the trial start date, drawn from a gamma distribution fitted to clinical trials with complete date information in our data (see Figure 2).

**Figure 2:**
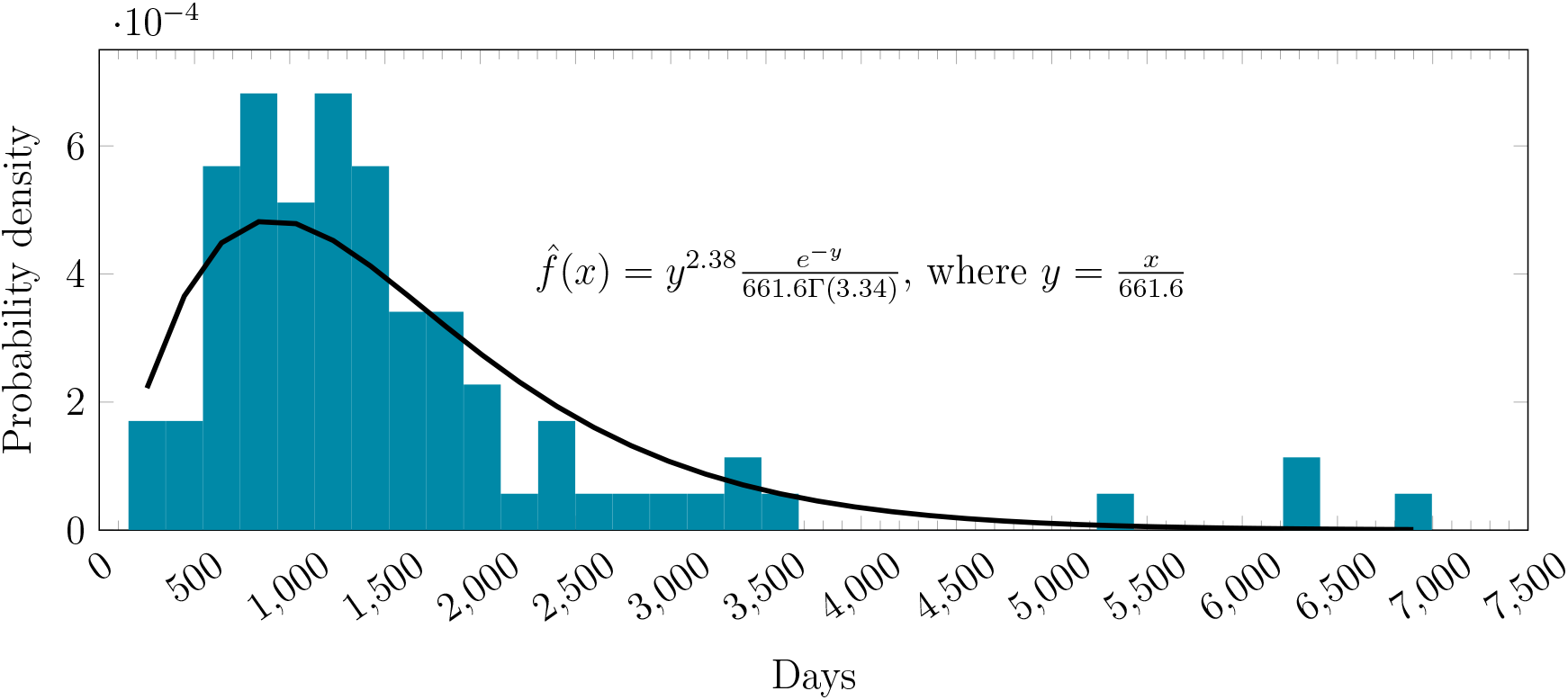
The empirical distribution of duration against our fitted gamma distribution.

Diseases with a prior approved therapy will automatically be considered to be approved as of January 1st 2020. For some diseases, the last clinical trial will have ended more than three years ago (i.e., before January 2017). For diseases that match this criterion, we treat them as though they have failed.

### 2.4 Number of Patients

The second module simulates the number of new and existing patients that will be treated over time, conditioned on the disease receiving an approved gene therapy.

We consider only the superset of the patient segments listed in the clinical trials for each disease. For example, if there are two clinical trials, one targeting ‘patients above the age of 40’ and the other targeting ‘patients above the age of 18’, we only consider the latter when estimating the patient population for the disease. If insufficient information about the sub-population is given, we assume that all the patients with that disease are eligible. The proportion of patients who are eligible for treatment and are willing to do so will be taken into account later, as we will explain in Section 2.4.

#### Incidence and Prevalence

For the number of currently affected patients and the number of new patients per year for each indication, we source medical journals and online data repositories, such as the Surveillance, Epidemiology, and End Results (SEER) website and *cancer*.*net*. If we are able to find an estimated patient population, we cite it directly. Otherwise, we multiply the prevalence and incidence rates by the population of the U.S., which we take to be 327.7 million [74]. When necessary, we also make the assumption that the female to male ratio is 1:1.

In cases in which we are able to find estimates for the disease incidence but not the prevalence, we combine the incidence of the disease (i.e., *i* new patients a year) and the disease survival rate (i.e., *p*% of the people with a disease will be alive after *k* years) to obtain the steady-state estimate of the prevalence (*j*) using Equation Equation 1. The incidence can also be estimate from the prevalence by rearranging Equation Equation 1 to yield Equation 2.

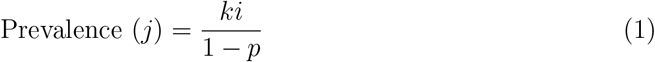

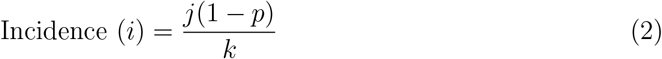

The equations can be derived by assuming that the number of patients will be constant through the years at a level *j*. Since *ki* new patients are added over *k* years and *j*(1 − *p*) patients that will die over the same period, *ki* = *j*(1 − *p*) for the number of patients to be constant over time. Rearranging this equation will yield Equation 1 and Equation 2. The number of patients for each disease are presented in Table A3 in the appendix. We adjust these estimates to avoid double-counting in cases of overlapping patient populations, e.g., the number of patients for ‘Spinal Muscular Atrophy’ is the difference between ‘Spinal Muscular Atrophy’ and ‘Spinal Muscular Atrophy I’ (a sub-category of the former).

#### Treatment of Patients over Time

In our simulation, we assume that newly diagnosed patients are treated immediately upon diagnosis. We further assume that the proportion of existing patients who seek treatment do so in such a way that the existing stock of patient declines exponentially, with a half-life of *λ*. Mathematically, the proportion of existing patients that seek treatment between time *t* and *t* + *δ* after approval is given by *E*(*t, δ, λ*), where:

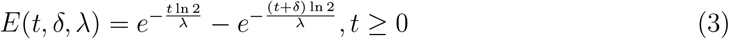

In the face of limited information, we assume that 25% of the existing stock of patients will seek treatment in the first year of our simulation. This requires that the half-life be set to 28.91 months, which in turn implies that 95% of all patients who are diagnosed prior to the approval of the gene therapy want treatments within 10.5 years. Admittedly, this is just an assumption, and we perform a sensitivity analysis to determine its impact on our results in Section 4.3. Not everyone who seeks treatment will be given one; the effective number of patients treated is determined by the patient penetration rate, as we shall describe next.

#### Patient Penetration

It is unlikely that all the patients under consideration will receive gene therapy treatments. This may be due to ineligibility, lack of awareness of the treatment, or simply lack of interest in gene therapy, among many other reasons. We term the percentage of the patients that receive gene therapy treatments the ‘patient penetration rate’, and model it using a ramp function, *ρ*(*t*, Θ_*max*_, *T*_*max*_). The ramp function is frequently used by the industry to model the rate of adoption of a product or technology [142], and is given by:

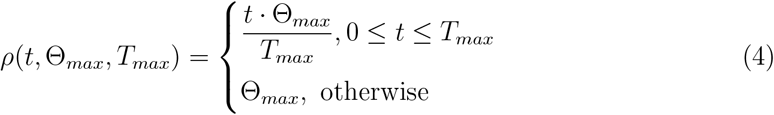

An illustration of the ramp function is given in Figure 3. Θ_*max*_ and *T*_*max*_ are assumed to follow Gaussian distributions 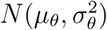 and 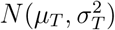, respectively. The parameter settings are listed in Tables 4 and 5. When setting *μ*_*θ*_ and *μ*_*T*_, we need to take into the account the nature of the diseases. At one extreme, we have rare diseases, which are often life-threatening, and affect a relatively small number of people. Faced with these prospects of survival, more patients are willing to enroll in new treatments quickly after they are approved. In addition, since the number of patients is relatively small, insurers are more willing to cover these therapies and manufacturers are more able to cope with a larger proportion of patients. Given this, we assign a high value of 40% for *μ*_*θ*_, and a low value of 6 months for *μ*_*T*_.

**Table 4:** Parameter settings for 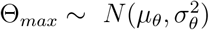.

| Classification | $\mu_\theta$ | $\sigma_\theta^2$ |
| --- | --- | --- |
| General | 0.01 | 0.002 |
| Rare Diseases | 0.4 | 0.08 |
| Cancer | 0.1 | 0.02 |

**Table 5:**
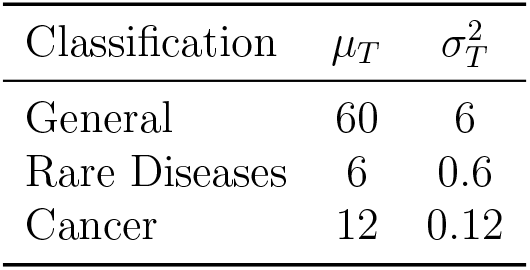
Parameter settings for 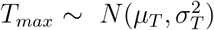. We consider the severity of the disease and the number of patients when making the assumptions.

**Figure 3:**
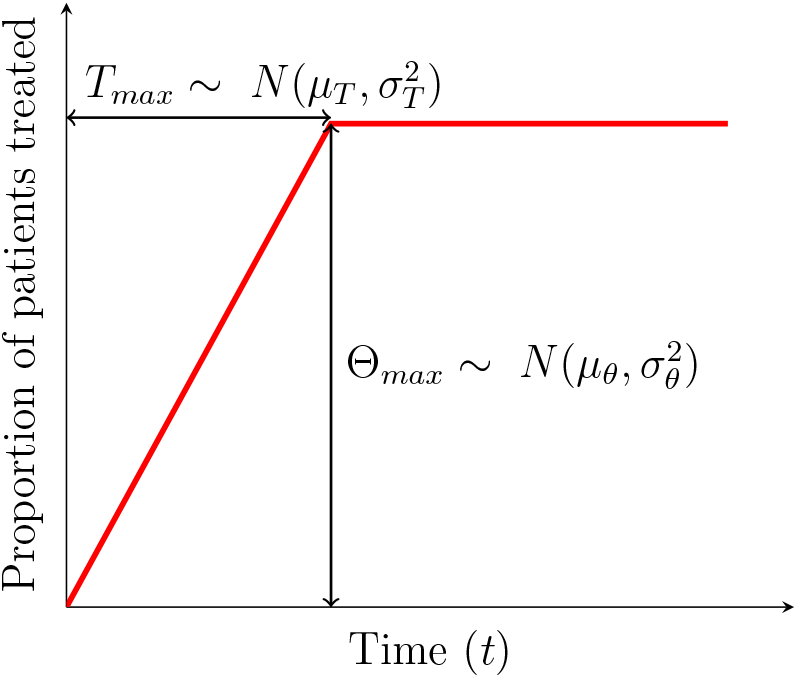
An illustration of the ramp function used to model the patient penetration rate over time.

On the other hand, general diseases are seldom deadly, and affect a large number of patients, possibly even in the millions. Since an acceptable standard of care is available for these conditions, patients may be less inclined to use new treatments due to fear of the unfamiliar. In addition, it is often financially difficult for insurance companies to cover so many patients. We thus assume that the maximum penetration rate will be 1%, and the ramp-up period, 5 years.

As an intermediate case, cancers have characteristics that fall between these two extremes, but in general, they are more similar to rare diseases. We therefore assign values of 10% and 12 months to the maximum penetration rate and ramp-up period, respectively. All variances are set to 10% of the means to model moderate uncertainty in our numbers. They do not affect our mean estimates of the number of impacted patients or spending on gene therapy.

The net number of patients to be treated for the disease at time *t* after the approval of a gene therapy is given by:

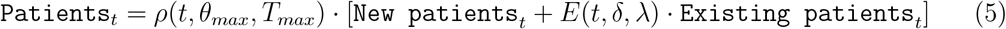

We do not consider the effect of market competition among different therapies for the same disease and patient groups on the number of treated patients. In our model, there is only one approval per disease, and a fraction of the eligible patients will receive that treatment.

## 3 Pricing

The cost to the healthcare system of providing the gene therapy for a disease for all patients being treated at time *t* after approval is given by *C*(*t*), where

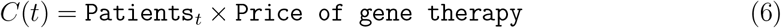

The price of each treatment is crucial to computing the expected total spending, and a source of considerable controversy because of the high price of gene therapies relative to many conventional therapeutics. The Institute for Clinical and Economic Review (ICER)— an independent nonprofit organization that aims to evaluate the clinical and economic value of healthcare innovation—has advocated pricing drugs and gene therapies by the relative risk and benefit to the patient. This is typically done by comparing the quality-adjusted life years (QALY) with and without the treatment, then multiplying the change in QALY (ΔQALY) by a constant, typically set between $50,000 and $150,000 per ΔQALY [135].

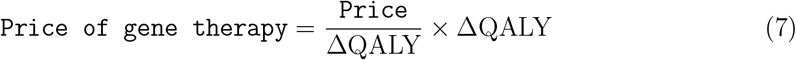

ICER has published reports containing its estimates of QALY gained by patients with vision loss associated with biallelic RPE65-mediated retinal disease following treatment with Luxturna^®^ [124], and with SMA Type I following treatment with Zolgensma^®^ [82]. These reports compute ΔQALY using the results of clinical trials to make informed estimates about the potential improvements in the quality of life and life expectancy of the patients.

While ICER’s methods are considered by some stakeholders to be the gold standard for this type of calculation, replicating its methods for all the clinical trials under consideration is not feasible in this paper, given the fact that all the clinical trials in our analysis are still pending. As an alternative, we develop a mathematical model to estimate the expected increase in QALY for each disease in our sample.

### 3.1 Estimating ΔQALY

We consider a representative patient who is expected to live to the age of *x* with a probability of *l*(*x*). The function *l*(*x*) is also known as the survival curve of the population. The patient enjoys a quality of life, *f* (*s, x*), that is dependent on his age, *x*, and his state of health, *s*. The expected QALY of a typical person in the baseline state of *s*_0_ (the ‘healthy’ state) can be computed by integrating *l*(*x*)*f* (*s*_0_, *x*) over *x*.

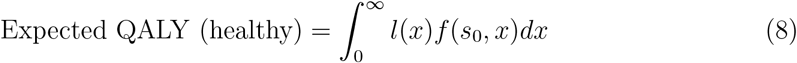

Suppose that the patient is afflicted with a disease at time *a*, which changes his survival curve after time *a* from *l*(*x*) to 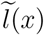. Likewise, his quality of life after diagnosis changes from *f* (*s*_0_, *x*) to *f* (*s*_*d*_, *x*). This patient will then have an expected QALY of:

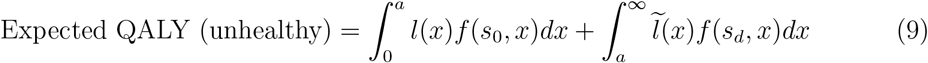

The change in the expected QALY due to the disease can then be expressed as:

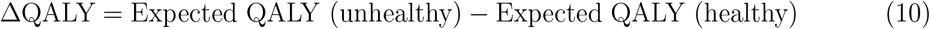

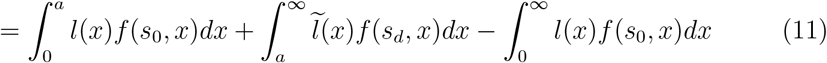

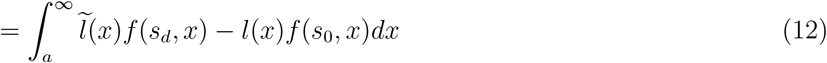

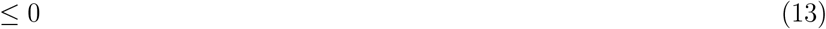

It is customary in the literature to incorporate time preferences into the model. This is done by multiplying the integrand by the discount factor, *r*(*x* − *a*). There is a normalization term *l*(*a*) to reflect conditional survival to age *x*.

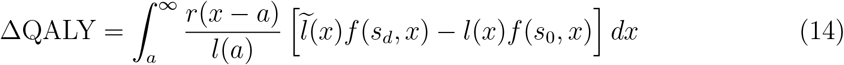

If the distribution of age when the patient population contracts the disease is given by *A*(*a*), then the expected decrease in QALY over the patient population is given by:

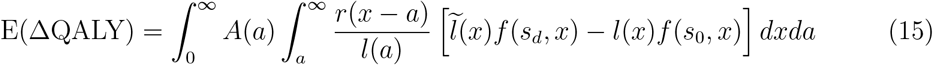

Equation 15 is a general formula that accounts for the expected value of the changes in QALY between two states of health using only three variables: the time of disease onset, and the utility of the two health states. By making the relevant substitutions, we can also apply this formula to compute the expected changes in QALY given a gene therapy (*gt*) and an alternative treatment (*alt*).

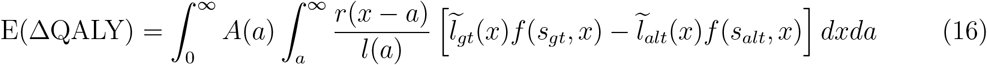

While death and patient statistics can be collected to determine *l*(*x*) and *A*(*a*) empirically, determining *f* (*s, x*) and 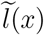 is challenging. Therefore, we use simple functions to modify these variables. In particular, we assume that being afflicted with a severe disease will modify the survival curve by a multiplicative factor, *D*(*t*). That is, the survival curve of a patient after he is diagnosed at age *a* is given by:

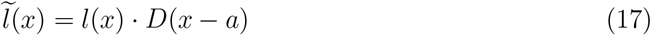

This functional form assumes that the disease is age-agnostic, and affects the survival curve only through the time elapsed since the patient has been diagnosed. For example, if the disease does not affect mortality (e.g. blindness), then *D*(*x* − *a*) = 1 for all *x* − *a >* 0. On the other hand, if the condition causes death immediately, then *D*(*x* − *a*) = 0 for all *x* − *a >* 0.

For the utility function, *f* (*s, x*), we assume that it can be decomposed into two multiplicative factors, one dependent only on age, *f*_*a*_(*x*), and the other dependent only on the state of health, *f*_*h*_(*s*):

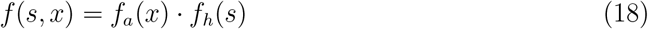

Assuming that Equations 17 and 18 hold, Equation 16 can be simplified to:

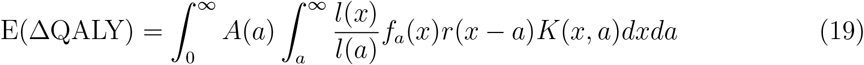

where *K*(*x, a*) is the change in the quality-adjusted life years:

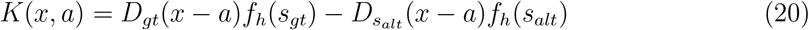

### 3.2 Calibration of ΔQALY

For each of these variables, we attempt to obtain empirical values from the literature as much as possible. When necessary, we interpolate values, briefly explaining our assumptions and the data collection methods for the inputs to the model.

For the age-dependent QoL, *f*_*a*_(*x*), we extract the general population utility values from Institute for Clinical and Economic Review [115] and fit a linear model across the data. The QoL values and the fitted model are shown in Figure 4.

**Figure 4:**
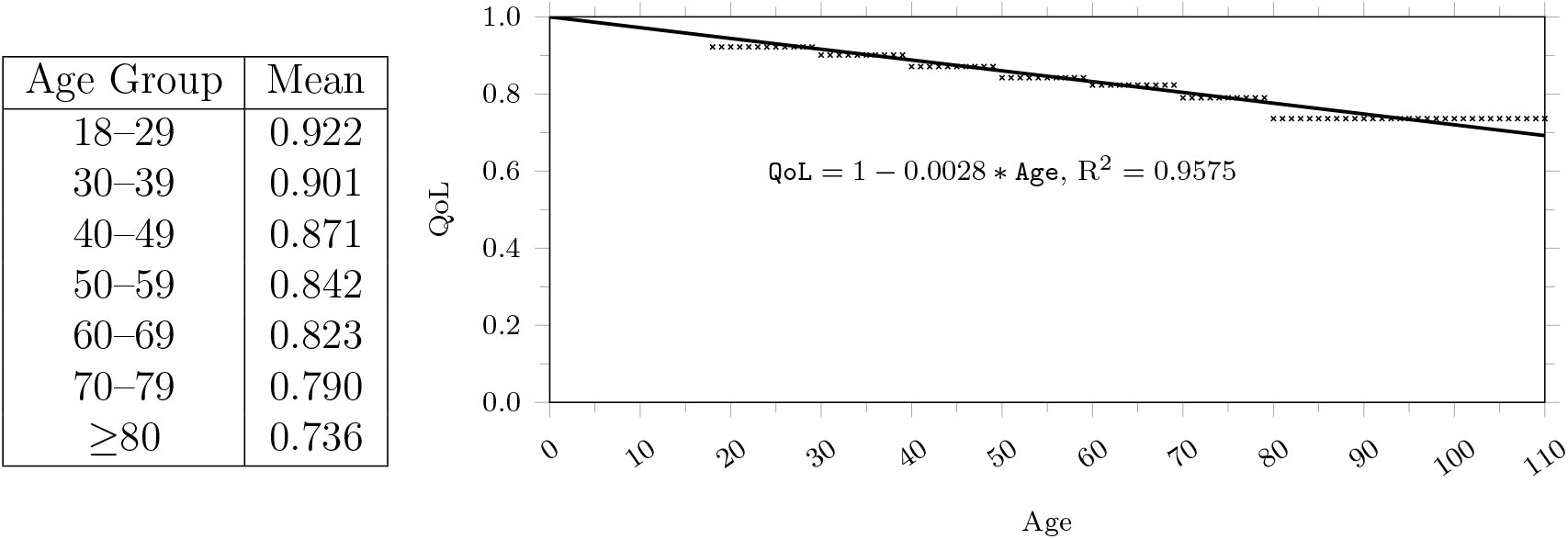
Age-dependent QoL, *f*_*a*_(*x*). The values extracted from ICER’s SMA final report [115] are replicated in the table on the left and are presented as crosses in the figure on the right. A linear line, with its intercept set to 1, is fitted with the data points.

Since we are unable to know the patient outcomes for these potential gene therapies ahead of their approval, we assume that the gene therapy treatments will restore a person’s survivability to that of a normal individual. This implies that *D*_*gt*_(*x* − *a*) = 1. To estimate the impact of a disease on patient survivability, we model its survival curve, *D*_*alt*_(*x* − *a*), using the exponential survival curve shown in Equation 21. In the equation, *λ* is the force of mortality, and *μ* is the normalization factor. We estimate *λ* and *μ* by matching the function to *T* -year survival rates, which are the proportions of the patients (*k*) who will be alive after *T* years, from data. The parameter values and their sources are listed in Table A4 in the Supplementary Material.

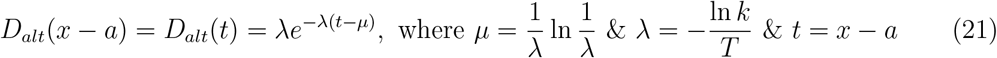

The health-related quality of life variables, *f*_*h*_(*s*_*gt*_) & *f*_*h*_(*s*_*alt*_), are treated separately, depending on the disease classification. For cancerous indications, we assume that the quality of life of the patients is not affected by the disease. For non-cancerous indications, we source the medical literature for the available quality of life (QoL) estimates. We use the QoL for the typical disease condition to approximate the ‘before treatment’ QoL, *f*_*h*_(*s*_*alt*_), and use the best possible outcome for each condition as the ‘post-treatment’ QoL, *f*_*h*_(*s*_*gt*_). We interpolate the missing values using linear regressions of the sourced QoLs against disease severity. To do this, we first give scores, *ζ*, ranging from one to five for each disease, based on our perception of disease severity. We then fit a line of *f*_*h*_(*s*_*alt*_) against *ζ* in order to estimate the missing ‘before treatment’ QoL values, *f*_*h*_(*s*_*alt*_) (see Figure 5). We define ΔQoL = *f*_*h*_(*s*_*gt*_) − *f*_*h*_(*s*_*alt*_). Separately, we regress ΔQoL against *ζ* to interpolate the change in QoL (see Figure 6). Given ΔQoL and *f*_*h*_(*s*_*alt*_), we can then estimate the missing values of *f*_*h*_(*s*_*gt*_). Our estimated values are reported in Table A5 in the Supplementary Material.

**Figure 5:**
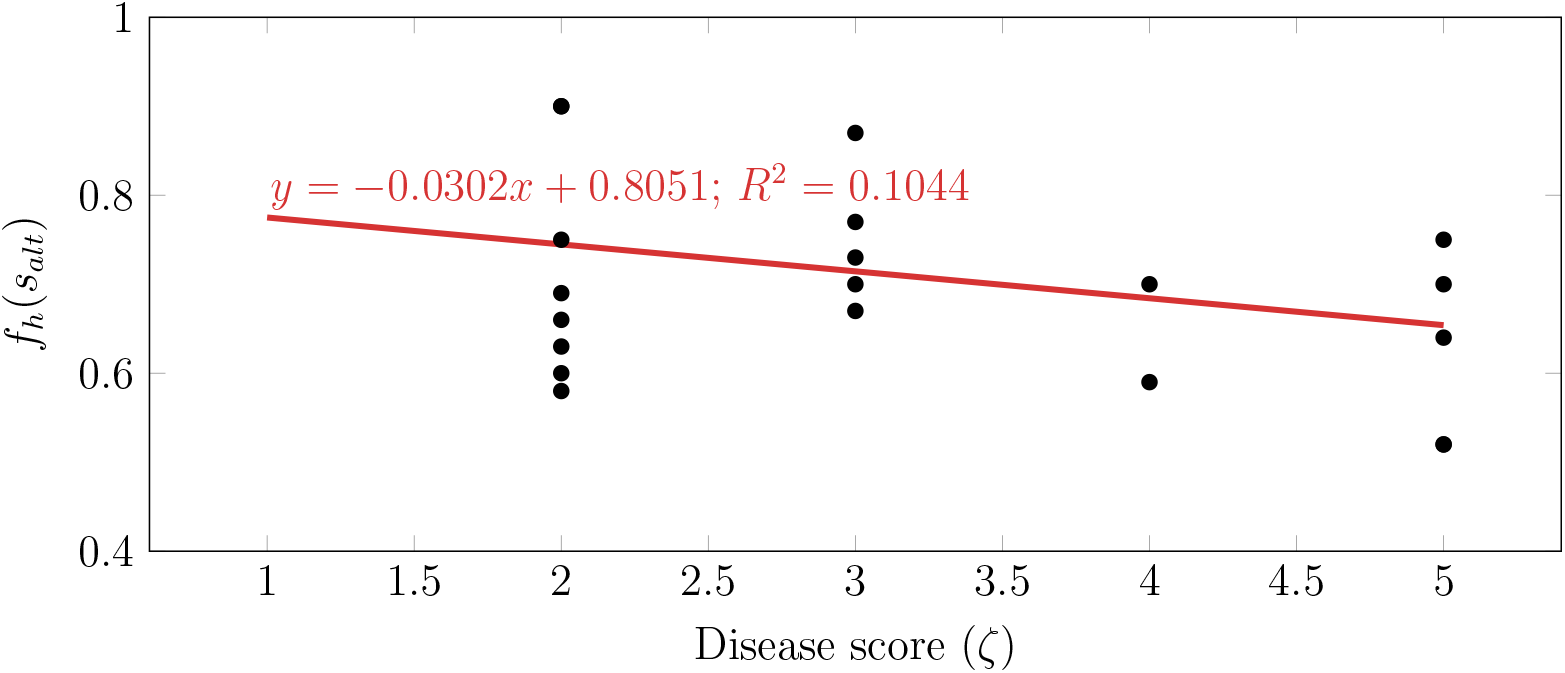
Scatter plot of *f*_*h*_(*s*_*alt*_) against disease score (*ζ*)

**Figure 6:**
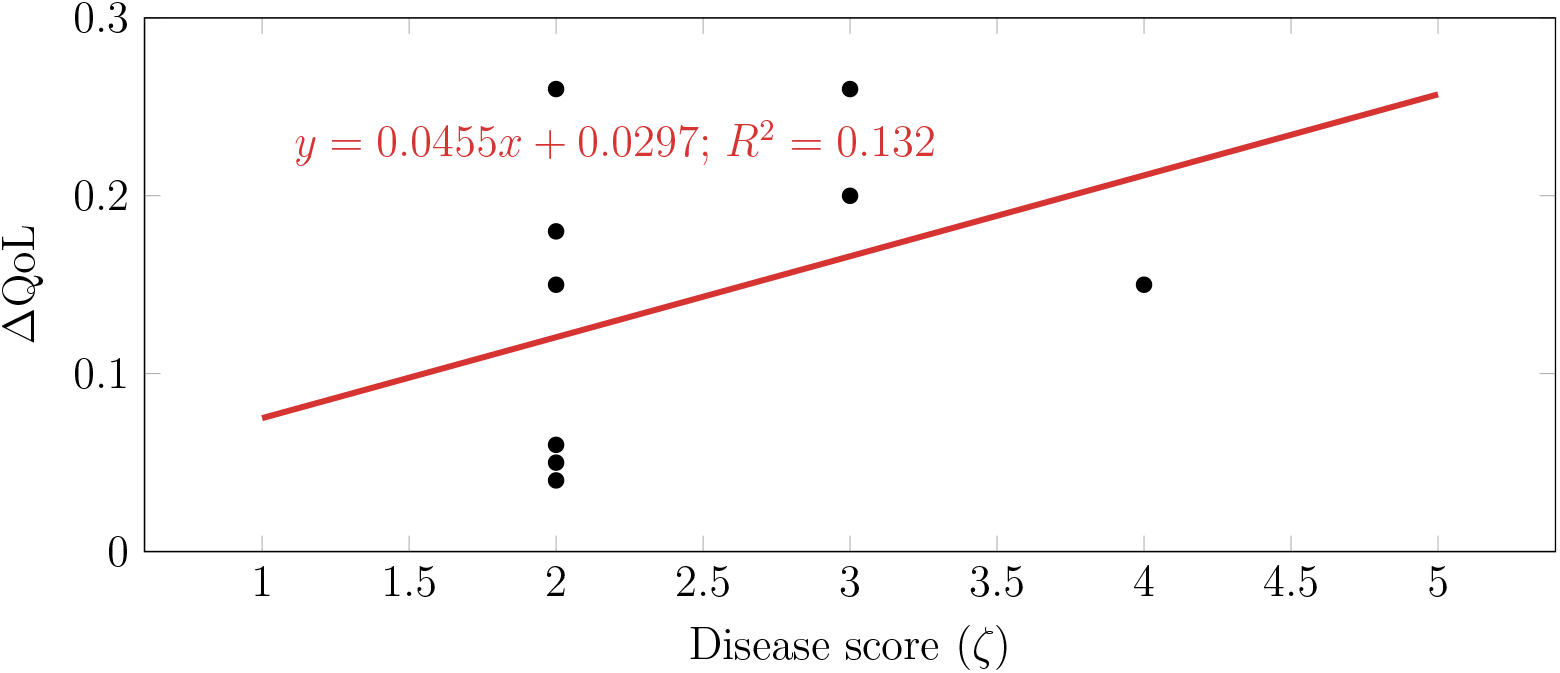
Scatter plot of ΔQoL against disease score (*ζ*)

We are able to extract the distribution of the age of cancer onset from the National Cancer Institute’s Surveillance, Epidemiology, and End Results (SEER) Program website. However, empirical age distributions are practically nonexistent for non-cancerous diseases. To overcome this lack, we search the literature for the average age of diagnosis of each disease, and fit a triangle distribution for each disease using the optimization program shown in Figure 7.

**Figure 7:**
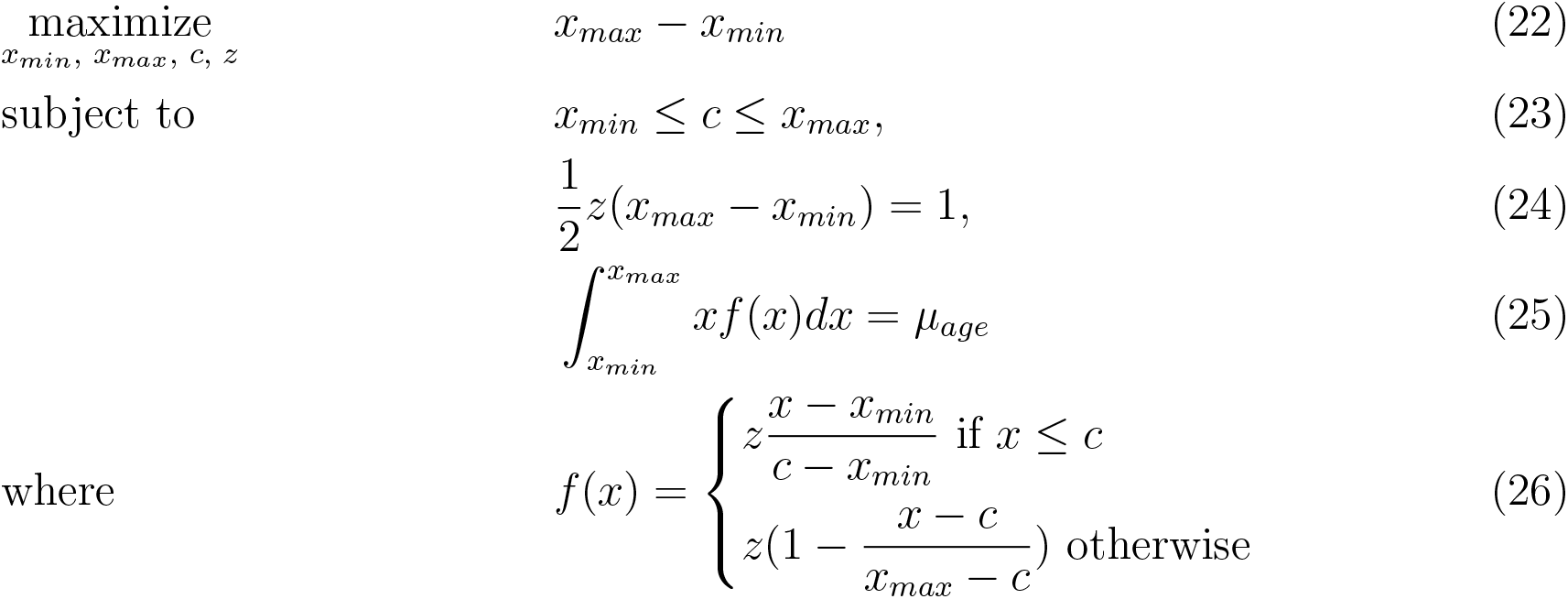
An optimization program to obtain the triangle distribution given the average age of diagnosis, *μ*_*age*_. *x*_*min*_ and *x*_*max*_ are coordinates of the base of the triangle. *c* and *z* are the mode and height of triangle.

This program maximizes the domain’s interval (Equation 22) while imposing the requirement that the distribution’s mode, *c*, has to be in the domain (Equation 23). In addition, the area under the curve has to be equal to 1 (Equation 24), and the mean of the distribution has to be equal to the average age (Equation 25).

We experimented with an impulse function and an uniform distribution to model the age distribution, but these functions created unrealistic scenarios. Modeling the age distribution with the impulse function, while simple, will force Equation 15 to collapse into a single point, and lose any nuance in the QALY gained by patients of different ages. On the other hand, estimating a uniform distribution from the average age creates distributions with narrow support. The distributions from our optimization program have a wider base of support and avoid sharp changes in density. We illustrate this with some examples in Figure A1 in the Supplementary Materials.

We assume a 3% per annum discount rate, as suggested by ICER for high-impact single or short-term therapy (SST) [92].

### 3.3 Price per ΔQALY

To estimate as realistic a market price of gene therapy as possible, we calibrate our assumed price per ΔQALY with the 4 data points currently available: Zolgensma, priced at $2.1 million per patient [132], Luxturna, priced at $0.425 million per eye treated [157], Kymriah, priced at $0.475 million for a one-time dose [67], and Yescarta, priced at $0.373 million for a one-time dose [67]. Separately, Zynteglo, sold at a cost of 1.6 million Euros (approximately $1.8 million), has been approved in the European Union. The data points are listed in Table 6.

**Table 6:** Diseases under consideration, approved gene therapy treatments used as proxy, prices of approved treatments, countries/areas in which treatments have been approved, and computed expected change in QALY.

| Disease | Approved treatment | Country approved | List price | E(ΔQALY) |
| --- | --- | --- | --- | --- |
| Beta-Thalassemia | Zynteglo | E.U. | 1.8M | 4.58 |
| Diffuse Large B Cell Lymphoma (DLBCL) | Yescarta | U.S. | 0.373M | 6.19 |
| Leber Congenital Amaurosis due to RPE65 Mutations | Luxturna | U.S. | 0.425M | 4.63 |
| Leukemia (Acute Lymphoblastic) | Kymriah | U.S. | 0.475M | 13.02 |
| Spinal Muscular Atrophy Type 1 | Zolgensma | U.S. | 2.125M | 20.56 |

We calibrate the price per ΔQALY by minimizing the mean-squared error (MSE) between the estimated price given the expected change in QALY and the actual price. We report the mean absolute percentage error (MAPE) between the estimated price and the actual price in addition to the MSE. We note that Zolgensma, Zynteglo, and Luxturna are gene replacement therapies for rare diseases, while Kymriah and Yescarta are chimeric antigen receptor T-cell (CAR-T) therapies indicated for cancers. As such, we perform two separate calibrations, one for rare diseases and the other for cancerous indications. We assume that the price per ΔQALY for general diseases is identical to that for cancerous indications.

Considering only the therapies approved in the United States, we estimate a price per E(ΔQALY) of $101,663 (MSE: 2.18*×*10^9^, MAPE: 11.2%) for rare diseases and $40,797 (MSE: 1.77*×*10^1^0, MAPE: 44.2%) for other diseases. Using all the data points, the price per E(ΔQALY) for rare diseases increases to $114,781 (MSE: 1.70*×*10^12^, MAPE: 108%). In this paper, we use the former for our calculations since it has a smaller mean-squared error and better reflects prices in the U.S., our focus. This value will give us estimates of $2.09M per patient for Zolgensma and $0.470M per eye for Luxturna.

Our calibrated price per E(ΔQALY) for cancerous indications is just slightly below ICER’s $50,000 to $100,000 range for ‘intermediate care value’. The higher price per E(ΔQALY) for rare diseases also reaffirms the general belief that developers of treatments for rare diseases should be compensated more for their elevated R&D risk and the low financial prospects of serving a small population of patients. It is assumed that the clinical cost of delivering the gene therapy is a negligible fraction of the overall cost of development (though they are considerably higher than the delivery cost of conventional therapeutics). It is also highly likely that the outside option cost will be similar.

The expected increases in QALY computed by our model are close to those provided by the ICER reports for the treatments [115, 124]. For example, we estimate that treatments for Spinal Muscular Atrophy Type 1 and Leber Congenital Amaurosis due to RPE65 Mutations provide 20.56 and 4.63 incremental QALYs, whereas ICER estimates Zolgensma and Luxturna to provide 12.23 to 26.58 and 1.3 to 2.7 incremental QALYs^4^, respectively. We have deliberately applied the same methods and assumptions used for the all other diseases to estimate the expected changes in QALY for Spinal Muscular Atrophy Type 1 and Leber Congenital Amaurosis due to RPE65 Mutations even though we could have obtained these numbers directly from ICER reports. By doing so, our price per ΔQALY calibration will correct for potential biases in our data, and our price estimates will be more realistic.

Our estimated change in QALY, the price per unit change in QALY, and the estimated price of therapy for each disease are shown in Table 7.

**Table 7:** Estimated ΔQALY, assumed price per ΔQALY and estimated price of gene therapies per disease. Prices are given to 3 significant figures for display in this table.

| Disease | $\Delta QALY$ | $\frac{\text{Cost}}{\Delta QALY}$ (\$) | Price (\$) |
| --- | --- | --- | --- |
| **General Diseases:** |  |  |  |
| Arteriosclerosis Obliterans | 2.96 | 41K | 121K |
| Critical Limb Ischemia | 7.32 | 41K | 299K |
| Degenerative Arthritis | 3.53 | 41K | 144K |
| Diabetic Foot Ulcers | 7.92 | 41K | 323K |
| Diabetic Peripheral Neuropathy | 3.95 | 41K | 161K |
| Heart Failure | 6.92 | 41K | 282K |

Table 7 – continued from previous page
| Disease | $\Delta QALY$ | $\frac{\text{Cost}}{\Delta QALY}$ (\$) | Price (\$) |
| --- | --- | --- | --- |
| Knee Osteoarthritis with Kellgren & Lawrence Grade 3 | 10.62 | 41K | 433K |
| Parkinson's Disease | 8.26 | 41K | 337K |
| Peripheral Artery Disease | 4.52 | 41K | 184K |
| Refractory Angina due to Myocardial Ischemia (AFFIRM) | 3.80 | 41K | 155K |
| Stable Angina | 3.85 | 41K | 157K |
| **Rare Diseases:** |  |  |  |
| Beta-Thalassemia | 4.58 | 102K | 466K |
| Cerebral Adrenoleukodystrophy (CALD) | 20.33 | 102K | 2.07M |
| Choroideremia | 4.24 | 102K | 431K |
| Cystic Fibrosis | 13.20 | 102K | 1.34M |
| Ewing's Sarcoma | 14.04 | 102K | 1.43M |
| Hemophilia A | 11.18 | 102K | 1.14M |
| Hemophilia B | 10.63 | 102K | 1.08M |
| Leber Congenital Amaurosis due to RPE65 Mutations | 4.63 | 102K | 470K |
| Leber Hereditary Optic Neuropathy | 3.97 | 102K | 404K |
| Lipoprotein Lipase Deficiency (LPLD) | 5.74 | 102K | 584K |
| Metachromatic Leukodystrophy | 21.06 | 102K | 2.14M |
| Mucopolysaccharidosis Type IIIa | 16.27 | 102K | 1.65M |
| Recessive Dystrophic Epidermolysis Bullosa | 5.89 | 102K | 599K |
| Retinitis Pigmentosa | 3.28 | 102K | 333K |
| Sickle Cell Anemia | 7.36 | 102K | 748K |
| Spinal Muscular Atrophy | 19.23 | 102K | 1.96M |
| Spinal Muscular Atrophy Type 1 | 20.56 | 102K | 2.09M |
| **Cancer:** |  |  |  |
| B-Cell Non-Hodgkin's Lymphoma | 4.90 | 41K | 200K |
| BCG Unresponsive NMIBC | 2.86 | 41K | 117K |
| Bladder Cancer, in situ concurrent with Papillary Tumors | 0.66 | 41K | 26.9K |
| Diffuse Large B Cell Lymphoma (DLBCL) | 6.19 | 41K | 253K |
| Head and Neck Cancer | 6.13 | 41K | 250K |
| Hepatocellular Carcinoma | 9.30 | 41K | 380K |
| High-Grade Glioma | 12.56 | 41K | 512K |
| Leukemia (Acute Lymphoblastic) | 13.04 | 41K | 532K |
| Leukemia (Acute Myelogenous) | 8.55 | 41K | 349K |
| Lymphoma | 4.90 | 41K | 200K |
| Melanoma (Locally Advanced Cutaneous) | 6.23 | 41K | 254K |
| Melanoma (Metastatic) | 9.22 | 41K | 376K |

Table 7 – continued from previous page
| Disease | $\Delta QALY$ | $\frac{\text{Cost}}{\Delta QALY}$ (\$) | Price (\$) |
| --- | --- | --- | --- |
| Multiple Myeloma (Newly Diagnosed) | 5.90 | 41K | 241K |
| Nasopharyngeal Carcinoma | 5.20 | 41K | 212K |
| NSC Lung Cancer | 7.04 | 41K | 287K |
| NSC Lung Cancer Stage 3 | 6.52 | 41K | 266K |
| Oral Cancer (Advanced) | 8.21 | 41K | 335K |
| Ovarian Cancer (Platinum-Resistant) | 10.83 | 41K | 442K |
| Ovarian Cancer, Primary Peritoneal Cavity Cancer | 7.93 | 41K | 324K |
| Pancreatic Cancer (Locally Advanced) | 7.64 | 41K | 312K |
| Prostate Cancer | 0.42 | 41K | 17.1K |
| Prostate Cancer (Localized) | 0.42 | 41K | 17.1K |
| Prostate Cancer (Metastatic Hormone-Refractory) | 7.75 | 41K | 316K |
| Prostate Cancer (Newly Diagnosed) | 0.99 | 41K | 40.4K |
| Recurrent Glioblastoma | 12.55 | 41K | 512K |
| Relapsed and Refractory Multiple Myeloma (RRMM) | 8.33 | 41K | 340K |
| Squamous Cell Cancer of Head and Neck or Esophagus | 5.51 | 41K | 225K |
| Synovial Sarcoma | 8.65 | 41K | 353K |

## 4 Results

All the equations are discretized from their continuous form in our computations. When solving integrals using the trapezoidal rule to obtain the ΔQALY, we use strip widths of 1 year for an age range from 0 to 110 years old, the resolution offered by the life tables. When simulating the number of patients and the cost over time, we use time intervals of 1 month. Our code is implemented on Python 3.6 backed by Numpy, and executed on a single 2.2GHz CPU core. Pseudo-code and further details of computation can be found in Section A8 in the Supplementary Materials.

We perform 1,000,000 iterations of the simulation to compute the mean number of patients and the total spending. With this number of iterations, one can expect the computed mean to be within 1.89% of the true mean 95% of the time (see Section A7 in the Supplementary Materials). We also report the 5th and 95th percentiles of the computed values as our upper and lower bounds respectively.

In the following section, we define a ‘minor’ to be a patient below the age of 18 and an ‘elderly’ patient to be one who is older than 62 years old. The remainder of the patients are labeled as ‘adults’.

### 4.1 Expected Number of Approvals and Patients

Our simulations indicate that the expected number of gene therapies approved between January 2020 and January 2034 is 18.3, with a 90% confidence interval of [14.0, 23.0] (see Figure 8).

**Figure 8:**
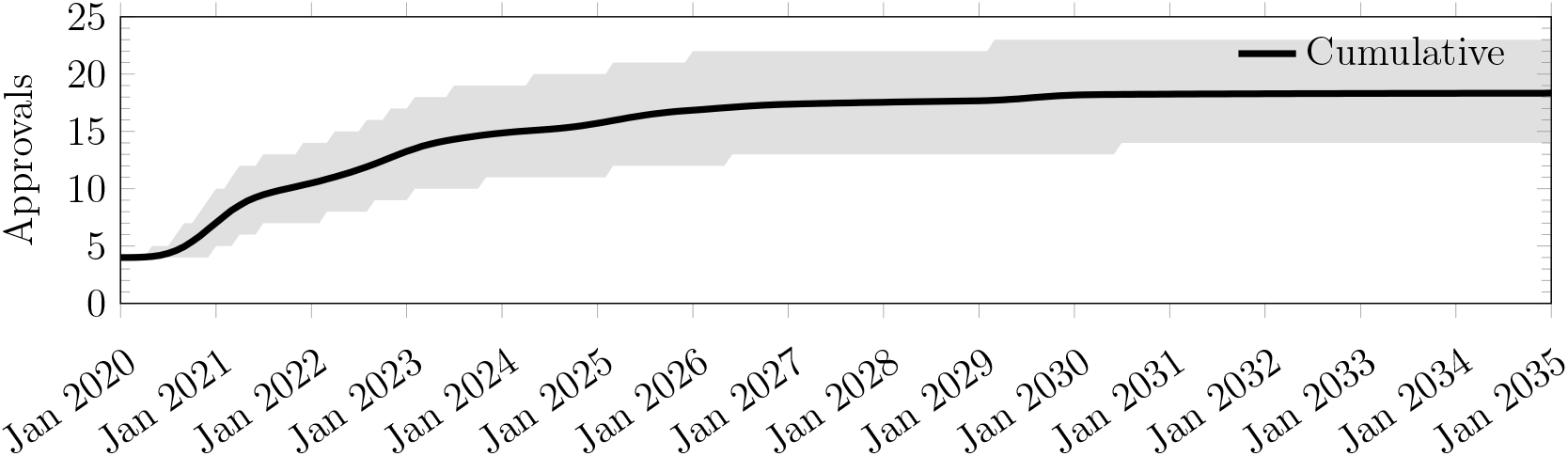
Cumulative number of approvals between January 2020 and December 2034, obtained from 1,000,000 simulation runs.

Table 8 shows the annual number of patients over time by age groups. Our simulations expect the number of patients treated to grow from 16,244 in 2020 to 94,696 in 2025 before declining to 65,612 in 2034. The decline can be attributed to the declining stock of existing patients as they are treated, and the fact that we do not consider new development programs launched in the future. The proportions of patients who are minors, adults and elderly are 17.9%, 35.4%, and 46.7% respectively.

**Table 8:** Expected annual number of patients treated by gene therapy between 2020 and 2035, conditioned on the age group and patient type. ‘Minor’, ‘adult’ and ‘elderly’ are defined to be ‘below the age of 18’, ‘between the ages of 18 and 62’, and ‘greater than 62 years old’, respectively.

| Year | Existing | Minor<br>New | Subtotal | Existing | Adult<br>New | Subtotal | Existing | Elderly<br>New | Subtotal | Total |
| --- | --- | --- | --- | --- | --- | --- | --- | --- | --- | --- |
| 2020 | 1,630<br>(1,283, 1,995) | 417<br>(322, 522) | 2,047<br>(1,614, 2,500) | 3,682<br>(2,557, 4,943) | 1,657<br>(1,080, 2,454) | 5,339<br>(3,670, 7,237) | 6,006<br>(3,928, 8,282) | 2,853<br>(1,813, 4,142) | 8,859<br>(5,785, 12,250) | 16,244<br>(11,349, 21,685) |
| 2021 | 2,833<br>(2,001, 4,357) | 1,294<br>(681, 1,950) | 4,127<br>(2,770, 6,326) | 9,921<br>(5,857, 14,948) | 6,399<br>(2,970, 11,725) | 16,320<br>(9,380, 24,834) | 14,278<br>(8,578, 21,422) | 9,540<br>(4,783, 17,438) | 23,818<br>(14,050, 36,234) | 44,265<br>(26,876, 65,911) |
| 2022 | 3,832<br>(2,000, 8,352) | 3,235<br>(947, 14,178) | 7,067<br>(3,224, 22,601) | 16,809<br>(6,965, 31,751) | 11,041<br>(4,364, 19,680) | 27,849<br>(12,843, 47,995) | 23,112<br>(9,343, 46,360) | 15,513<br>(6,561, 28,346) | 38,625<br>(17,258, 69,494) | 73,543<br>(35,001, 126,974) |
| 2023 | 4,722<br>(2,001, 11,596) | 5,612<br>(1,243, 25,879) | 10,334<br>(3,614, 37,353) | 19,364<br>(7,340, 36,474) | 13,890<br>(5,999, 23,487) | 33,254<br>(15,161, 56,955) | 25,862<br>(9,222, 52,202) | 19,031<br>(8,407, 32,988) | 44,893<br>(19,347, 80,954) | 88,482<br>(41,055, 151,872) |
| 2024 | 4,922<br>(1,832, 11,745) | 7,734<br>(1,490, 29,056) | 12,656<br>(3,681, 40,322) | 18,580<br>(7,284, 32,906) | 16,230<br>(7,683, 26,300) | 34,810<br>(16,794, 56,353) | 24,122<br>(8,817, 45,862) | 21,781<br>(10,261, 36,233) | 45,902<br>(21,011, 77,784) | 93,371<br>(45,504, 151,799) |
| 2025 | 5,235<br>(1,865, 11,736) | 9,621<br>(1,741, 30,664) | 14,856<br>(3,996, 41,541) | 16,570<br>(6,585, 28,364) | 18,159<br>(9,026, 28,683) | 34,728<br>(17,320, 54,531) | 21,115<br>(7,795, 38,698) | 23,994<br>(11,748, 38,878) | 45,110<br>(21,451, 73,552) | 94,696<br>(47,833, 148,985) |
| 2026 | 4,998<br>(1,667, 11,079) | 11,086<br>(1,918, 31,601) | 16,085<br>(3,996, 41,494) | 13,653<br>(5,511, 23,012) | 19,350<br>(9,839, 30,163) | 33,003<br>(16,868, 50,992) | 17,220<br>(6,444, 30,948) | 25,370<br>(12,692, 40,529) | 42,592<br>(20,887, 67,946) | 91,682<br>(47,432, 141,917) |
| 2027 | 4,246<br>(1,323, 9,859) | 12,120<br>(2,004, 32,129) | 16,366<br>(3,676, 40,559) | 10,687<br>(4,403, 17,871) | 19,915<br>(10,254, 30,847) | 30,604<br>(15,948, 46,843) | 13,402<br>(5,129, 23,798) | 26,032<br>(13,206, 41,300) | 39,433<br>(19,847, 62,104) | 86,401<br>(45,218, 132,708) |
| 2028 | 3,522<br>(1,024, 8,638) | 12,842<br>(2,052, 32,455) | 16,364<br>(3,369, 39,508) | 8,203<br>(3,402, 13,700) | 20,129<br>(10,409, 31,105) | 28,332<br>(14,899, 43,197) | 10,215<br>(3,945, 18,068) | 26,242<br>(13,367, 41,550) | 36,457<br>(18,583, 57,103) | 81,153<br>(42,510, 124,357) |
| 2029 | 2,978<br>(851, 7,552) | 13,373<br>(2,114, 32,684) | 16,351<br>(3,217, 38,620) | 6,259<br>(2,594, 10,485) | 20,219<br>(10,480, 31,214) | 26,478<br>(13,969, 40,339) | 7,744<br>(3,002, 13,682) | 26,315<br>(13,432, 41,632) | 34,059<br>(17,475, 53,231) | 76,888<br>(40,221, 117,723) |
| 2030 | 2,656<br>(712, 6,628) | 13,807<br>(2,222, 32,909) | 16,463<br>(3,228, 38,011) | 4,764<br>(1,969, 8,027) | 20,276<br>(10,526, 31,277) | 25,040<br>(13,223, 38,155) | 5,863<br>(2,275, 10,366) | 26,361<br>(13,473, 41,682) | 32,224<br>(16,597, 50,343) | 73,726<br>(38,538, 112,779) |
| 2031 | 2,116<br>(540, 5,433) | 14,039<br>(2,241, 32,998) | 16,154<br>(3,032, 37,097) | 3,620<br>(1,490, 6,146) | 20,313<br>(10,558, 31,324) | 23,933<br>(12,635, 36,496) | 4,434<br>(1,720, 7,852) | 26,391<br>(13,501, 41,715) | 30,826<br>(15,896, 48,190) | 70,914<br>(36,887, 108,603) |
| 2032 | 1,654<br>(408, 4,332) | 14,185<br>(2,251, 33,049) | 15,840<br>(2,861, 36,273) | 2,746<br>(1,125, 4,698) | 20,338<br>(10,577, 31,355) | 23,084<br>(12,174, 35,255) | 3,349<br>(1,298, 5,938) | 26,411<br>(13,518, 41,736) | 29,759<br>(15,350, 46,585) | 68,684<br>(35,539, 105,370) |
| 2033 | 1,286<br>(308, 3,400) | 14,282<br>(2,257, 33,084) | 15,567<br>(2,726, 35,611) | 2,079<br>(847, 3,575) | 20,354<br>(10,591, 31,374) | 22,434<br>(11,813, 34,313) | 2,528<br>(978, 4,482) | 26,424<br>(13,529, 41,751) | 28,953<br>(14,926, 45,369) | 66,952<br>(34,479, 102,883) |
| 2034 | 993<br>(232, 2,644) | 14,344<br>(2,262, 33,106) | 15,337<br>(2,619, 35,063) | 1,572<br>(638, 2,713) | 20,364<br>(10,601, 31,387) | 21,937<br>(11,533, 33,602) | 1,906<br>(735, 3,379) | 26,432<br>(13,536, 41,761) | 28,338<br>(14,597, 44,479) | 65,612<br>(33,666, 100,944) |

We show the number of patients treated by month in Figure 9a. We can see that our simulations expect the number of patients treated to peak at around 7911 (CI: [3978, 12477]) per month in Jul 2025 before declining to 5424 (CI: [2778, 8350]) by December 2034. The monthly number of existing patients treated exceeds the monthly number of newly-diagnosed patients treated until Sep 2024, when this trend is expected to reverse (see Figure 9b). Only 7% of all patients treated in December 2034 are preexisting patients. Cancer patients are expected to form the biggest group of patients receiving gene therapy treatments, simply due to the number of cancer indications being targeted. We expect the relative proportions of cancer, general disease, and rare disease patients to be 48.0%, 30.0%, and 22.0%, respectively, in December 2034. The cumulative number of patients to be treated is expected to be 1.09 million (CI: [0.595M, 1.66M]) by the end of December 2034 (see Figure 9c).

**Figure 9:**
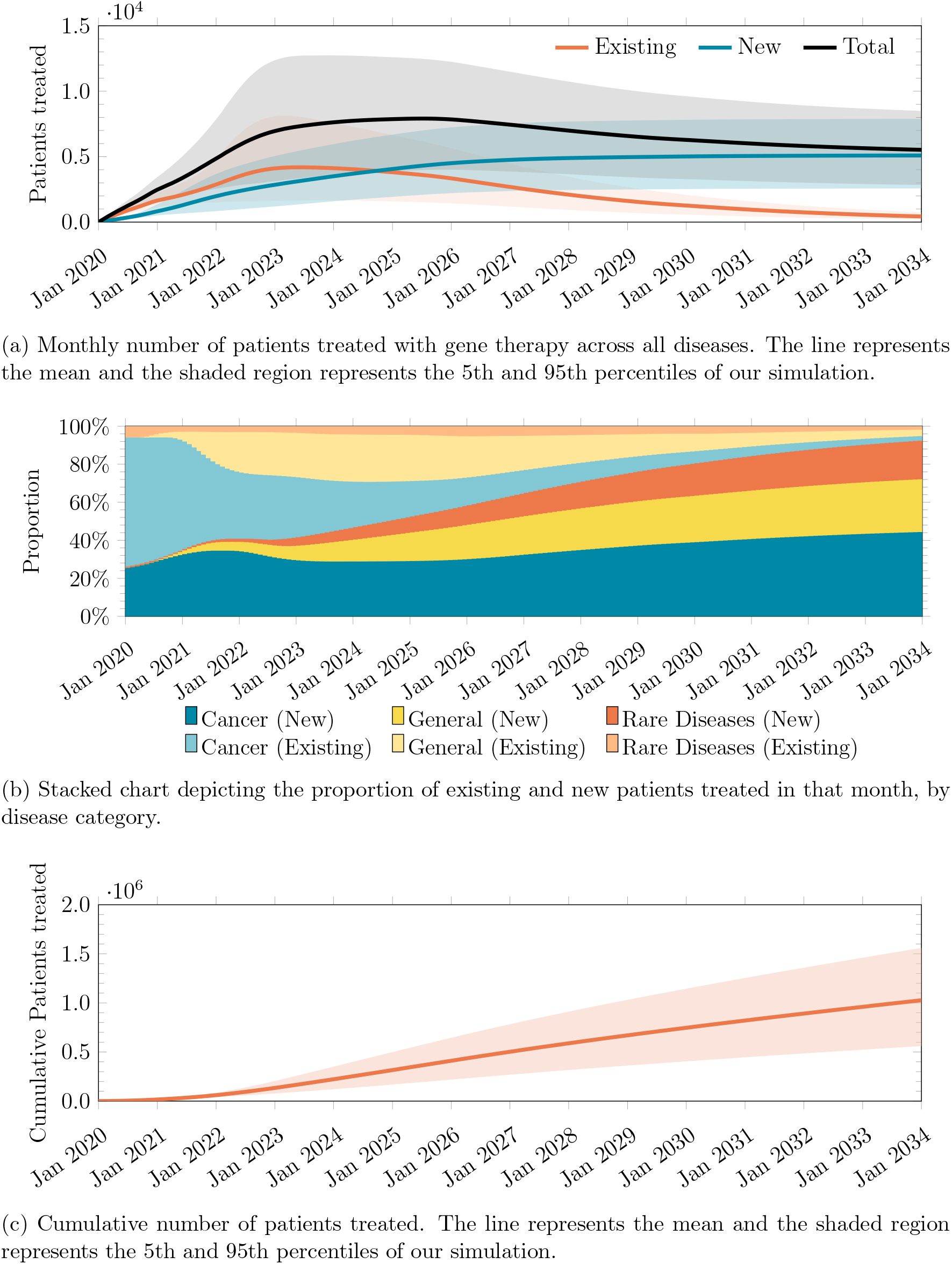
Number of patients treated between January 2020 and December 2034, obtained from 1,000,000 simulation runs.

### 4.2 Expected Spending

We expect an increase in spending up to $2.11 billion per month (CI: [1.01B, 3.88B]) in Apr 2026, before decreasing slowly to a steady-state rate of $1.62 billion (CI: [0.624B, 2.9B]) per month (see Figure 10a). We emphasize that the total spending eventually declines because our simulations analyze a fixed stock of innovations, and do not account for new development programs that may be launched in the future. Treating existing cancer patients initially consumes over 45.6% of the total monthly expenditure, but declines to only 0.99% by December 2034. In contrast, the proportion of spending on new patients in the ‘general disease’ and ‘rare disease’ groups will increase from 0.0% and 4.26%, respectively, in Feb 2020 to 21.2% and 46.2% by December 2034. The monthly spending on treating existing patients shall exceed the monthly spending on treating newly diagnosed patients in Nov 2023. The cumulative discounted spending on treating patients with approved gene therapy products is expected to reach $241 billion (CI: [123B, 402B]) by December 2034, 15 years after the start of our simulation.

**Figure 10:**
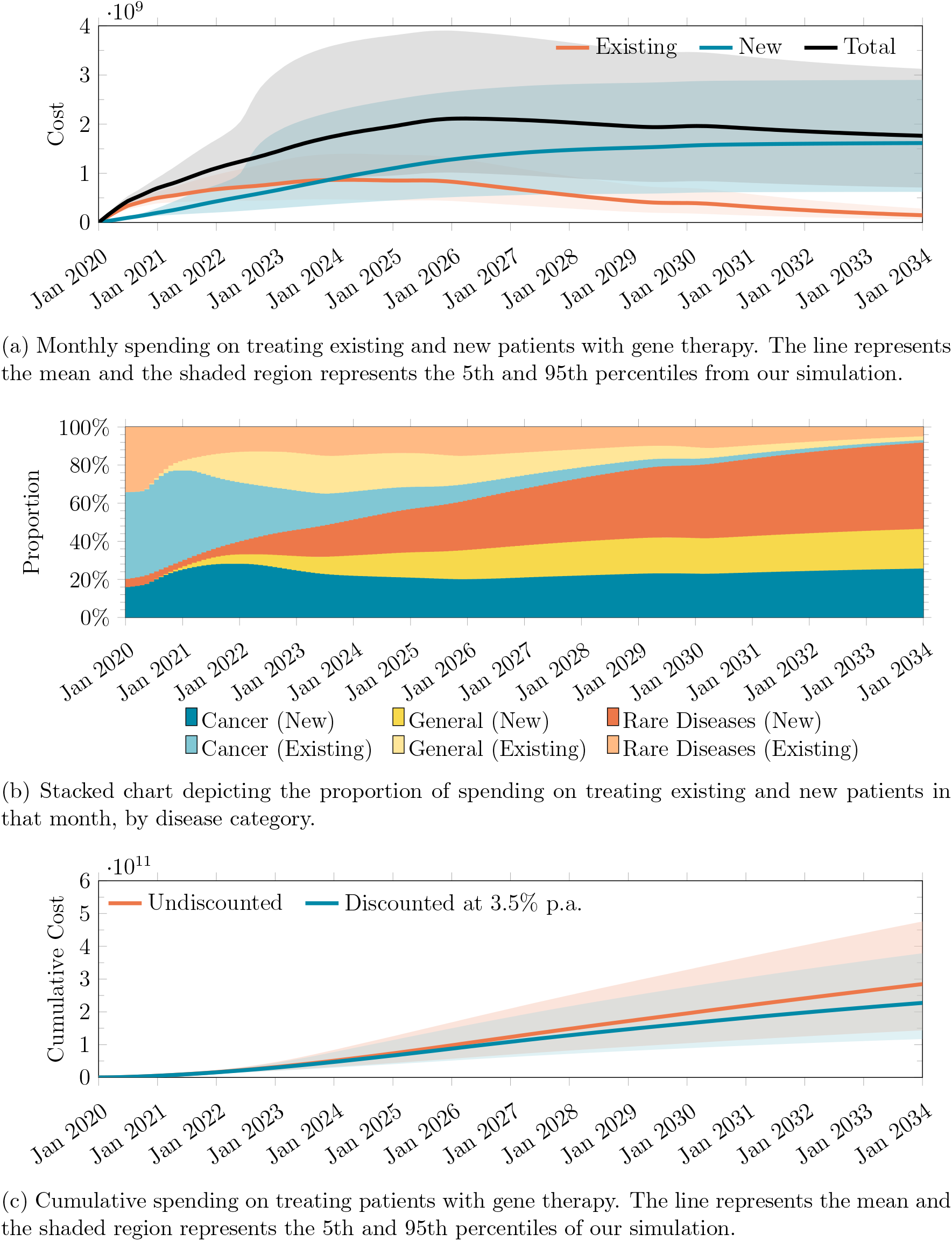
Spending on gene therapy between January 2020 and December 2034, obtained from 1,000,000 simulation runs.

In terms of annual spending on approved gene therapies, we expect that $5.15 billion will be spent in 2020, increasing to $25.3B in 2026 before declining to $21.0B in 2034 (see Table 9). Minors, adults and the elderly will consume 43.2%, 26.0%, and 30.9%, respectively, of the total spending. In the U.S., all elderly people are covered by Medicare. It is also estimated that two in five children and one in seven adults in the U.S. are covered by Medicaid [99]. The remainder of the spending is expected to come from private sources such as employer-provided or private insurance. Using these proportions, we estimate that the expected annual spending by Medicare, Medicaid ^5^ and private sources respectively may reach $8.1, $5.44, and $12.2 billion (see Table 10). We discuss the implications of these estimates in Section 4.4.

**Table 9:** Expected annual spending on gene therapy between 2020 and 2035, conditioned on the age group and patient type. ‘Minor’, ‘adult’ and ‘elderly’ are defined to be ‘below the age of 18’, ‘between the ages of 18 and 62’, and ‘greater than 62 years old’ respectively. Numbers in billions.

| Year | Existing | Minor<br>New | Subtotal | Existing | Adult<br>New | Subtotal | Existing | Elderly<br>New | Subtotal | Total |
| --- | --- | --- | --- | --- | --- | --- | --- | --- | --- | --- |
| 2020 | 1.77<br>(1.31, 2.25) | 0.32<br>(0.25, 0.40) | 2.10<br>(1.55, 2.65) | 0.91<br>(0.67, 1.18) | 0.37<br>(0.26, 0.53) | 1.29<br>(0.93, 1.69) | 1.20<br>(0.79, 1.65) | 0.57<br>(0.36, 0.80) | 1.77<br>(1.15, 2.45) | 5.15<br>(4.00, 6.39) |
| 2021 | 2.23<br>(1.60, 3.19) | 0.72<br>(0.42, 0.86) | 2.94<br>(2.03, 4.06) | 2.19<br>(1.43, 3.16) | 1.31<br>(0.68, 2.33) | 3.51<br>(2.17, 5.31) | 2.76<br>(1.73, 3.98) | 1.80<br>(0.96, 3.42) | 4.56<br>(2.77, 7.20) | 11.01<br>(7.47, 15.85) |
| 2022 | 2.37<br>(1.39, 5.76) | 1.79<br>(0.50, 9.98) | 4.16<br>(1.97, 15.78) | 2.91<br>(1.63, 4.41) | 2.09<br>(0.94, 3.70) | 5.01<br>(2.72, 7.76) | 3.52<br>(1.92, 5.30) | 2.70<br>(1.31, 5.03) | 6.22<br>(3.39, 9.99) | 15.38<br>(8.76, 27.15) |
| 2023 | 2.91<br>(1.26, 8.02) | 3.33<br>(0.60, 18.48) | 6.25<br>(1.97, 26.43) | 3.37<br>(1.80, 5.12) | 2.67<br>(1.30, 4.42) | 6.04<br>(3.34, 9.10) | 3.79<br>(1.97, 5.77) | 3.34<br>(1.71, 5.81) | 7.13<br>(3.88, 11.05) | 19.41<br>(10.30, 39.86) |
| 2024 | 3.06<br>(1.06, 8.12) | 4.79<br>(0.67, 20.70) | 7.85<br>(1.84, 28.46) | 3.46<br>(1.80, 5.27) | 3.26<br>(1.66, 5.17) | 6.72<br>(3.75, 9.99) | 3.84<br>(1.94, 5.89) | 4.02<br>(2.11, 6.65) | 7.86<br>(4.31, 11.92) | 22.43<br>(11.27, 44.10) |
| 2025 | 3.50<br>(0.98, 8.37) | 6.11<br>(0.77, 21.79) | 9.61<br>(1.95, 29.48) | 3.18<br>(1.62, 4.88) | 3.76<br>(1.96, 5.81) | 6.94<br>(3.85, 10.27) | 3.51<br>(1.74, 5.43) | 4.59<br>(2.42, 7.36) | 8.10<br>(4.44, 12.19) | 24.65<br>(12.04, 46.08) |
| 2026 | 3.49<br>(0.88, 8.08) | 7.15<br>(0.85, 22.43) | 10.64<br>(1.94, 29.57) | 2.67<br>(1.35, 4.12) | 4.08<br>(2.14, 6.25) | 6.75<br>(3.74, 10.00) | 2.94<br>(1.46, 4.53) | 4.96<br>(2.62, 7.84) | 7.89<br>(4.33, 11.85) | 25.28<br>(11.98, 45.94) |
| 2027 | 2.97<br>(0.70, 7.18) | 7.89<br>(0.88, 22.79) | 10.86<br>(1.77, 28.88) | 2.14<br>(1.08, 3.32) | 4.27<br>(2.24, 6.51) | 6.40<br>(3.53, 9.49) | 2.34<br>(1.17, 3.59) | 5.17<br>(2.74, 8.11) | 7.51<br>(4.13, 11.27) | 24.77<br>(11.28, 44.52) |
| 2028 | 2.47<br>(0.54, 6.30) | 8.42<br>(0.89, 23.01) | 10.89<br>(1.62, 28.11) | 1.66<br>(0.83, 2.63) | 4.34<br>(2.28, 6.61) | 6.00<br>(3.29, 8.93) | 1.80<br>(0.90, 2.75) | 5.24<br>(2.78, 8.20) | 7.04<br>(3.87, 10.61) | 23.92<br>(10.48, 42.84) |
| 2029 | 2.25<br>(0.48, 5.71) | 8.86<br>(0.95, 23.22) | 11.10<br>(1.62, 27.70) | 1.28<br>(0.64, 2.07) | 4.36<br>(2.30, 6.64) | 5.64<br>(3.08, 8.42) | 1.37<br>(0.69, 2.10) | 5.26<br>(2.79, 8.22) | 6.63<br>(3.64, 10.04) | 23.37<br>(10.03, 41.59) |
| 2030 | 2.37<br>(0.39, 5.52) | 9.31<br>(1.02, 23.53) | 11.68<br>(1.58, 27.89) | 0.98<br>(0.48, 1.62) | 4.38<br>(2.31, 6.66) | 5.36<br>(2.91, 8.02) | 1.04<br>(0.52, 1.60) | 5.27<br>(2.80, 8.24) | 6.31<br>(3.45, 9.60) | 23.35<br>(9.98, 41.08) |
| 2031 | 1.90<br>(0.30, 4.51) | 9.49<br>(1.03, 23.61) | 11.39<br>(1.47, 27.11) | 0.75<br>(0.36, 1.26) | 4.39<br>(2.32, 6.67) | 5.14<br>(2.78, 7.71) | 0.79<br>(0.39, 1.22) | 5.28<br>(2.81, 8.25) | 6.07<br>(3.31, 9.28) | 22.59<br>(9.47, 39.80) |
| 2032 | 1.47<br>(0.22, 3.57) | 9.60<br>(1.03, 23.64) | 11.07<br>(1.38, 26.37) | 0.57<br>(0.28, 0.97) | 4.40<br>(2.32, 6.68) | 4.97<br>(2.68, 7.47) | 0.60<br>(0.30, 0.93) | 5.28<br>(2.82, 8.25) | 5.88<br>(3.20, 9.03) | 21.92<br>(9.02, 38.69) |
| 2033 | 1.14<br>(0.17, 2.78) | 9.67<br>(1.04, 23.67) | 10.81<br>(1.30, 25.78) | 0.43<br>(0.21, 0.74) | 4.40<br>(2.33, 6.69) | 4.84<br>(2.60, 7.29) | 0.45<br>(0.22, 0.70) | 5.29<br>(2.82, 8.25) | 5.74<br>(3.11, 8.84) | 21.38<br>(8.67, 37.83) |
| 2034 | 0.87<br>(0.13, 2.15) | 9.72<br>(1.04, 23.68) | 10.59<br>(1.24, 25.31) | 0.33<br>(0.16, 0.57) | 4.41<br>(2.33, 6.69) | 4.73<br>(2.53, 7.15) | 0.34<br>(0.17, 0.53) | 5.29<br>(2.82, 8.26) | 5.63<br>(3.04, 8.70) | 20.95<br>(8.40, 37.13) |

**Table 10:** Expected annual spending on gene therapy between 2020 and 2035 by funding source. Numbers in billions.

|  | Medicare | Medicaid | Private |
| --- | --- | --- | --- |
| 2020 | 1.77<br>(1.15, 2.45) | 1.02<br>(0.79, 1.26) | 2.36<br>(1.87, 2.87) |
| 2021 | 4.56<br>(2.77, 7.20) | 1.68<br>(1.20, 2.28) | 4.77<br>(3.29, 6.78) |
| 2022 | 6.22<br>(3.39, 9.99) | 2.38<br>(1.27, 7.02) | 6.79<br>(3.79, 13.75) |
| 2023 | 7.13<br>(3.88, 11.05) | 3.36<br>(1.40, 11.44) | 8.92<br>(4.46, 21.11) |
| 2024 | 7.86<br>(4.31, 11.92) | 4.10<br>(1.43, 12.37) | 10.47<br>(4.84, 23.19) |
| 2025 | 8.10<br>(4.44, 12.19) | 4.83<br>(1.55, 12.82) | 11.71<br>(5.22, 24.14) |
| 2026 | 7.89<br>(4.33, 11.85) | 5.22<br>(1.55, 12.83) | 12.17<br>(5.20, 24.08) |
| 2027 | 7.51<br>(4.13, 11.27) | 5.26<br>(1.43, 12.50) | 12.01<br>(4.86, 23.37) |
| 2028 | 7.04<br>(3.87, 10.61) | 5.21<br>(1.32, 12.14) | 11.68<br>(4.50, 22.55) |
| 2029 | 6.63<br>(3.64, 10.04) | 5.25<br>(1.28, 11.92) | 11.50<br>(4.32, 21.97) |
| 2030 | 6.31<br>(3.45, 9.60) | 5.44<br>(1.26, 11.96) | 11.60<br>(4.33, 21.81) |
| 2031 | 6.07<br>(3.31, 9.28) | 5.29<br>(1.19, 11.61) | 11.24<br>(4.09, 21.13) |
| 2032 | 5.88<br>(3.20, 9.03) | 5.14<br>(1.13, 11.29) | 10.90<br>(3.89, 20.54) |
| 2033 | 5.74<br>(3.11, 8.84) | 5.01<br>(1.08, 11.03) | 10.63<br>(3.73, 20.06) |
| 2034 | 5.63<br>(3.04, 8.70) | 4.91<br>(1.04, 10.83) | 10.41<br>(3.60, 19.69) |

The total expected increase in QALY over these 15 years is 5.59 million (see Figure 11), which translates to an average increase of 5.12 in QALY per patient. This comes at an average 2020 present value cost of $43,110 per unit change in QALY.

**Figure 11:**
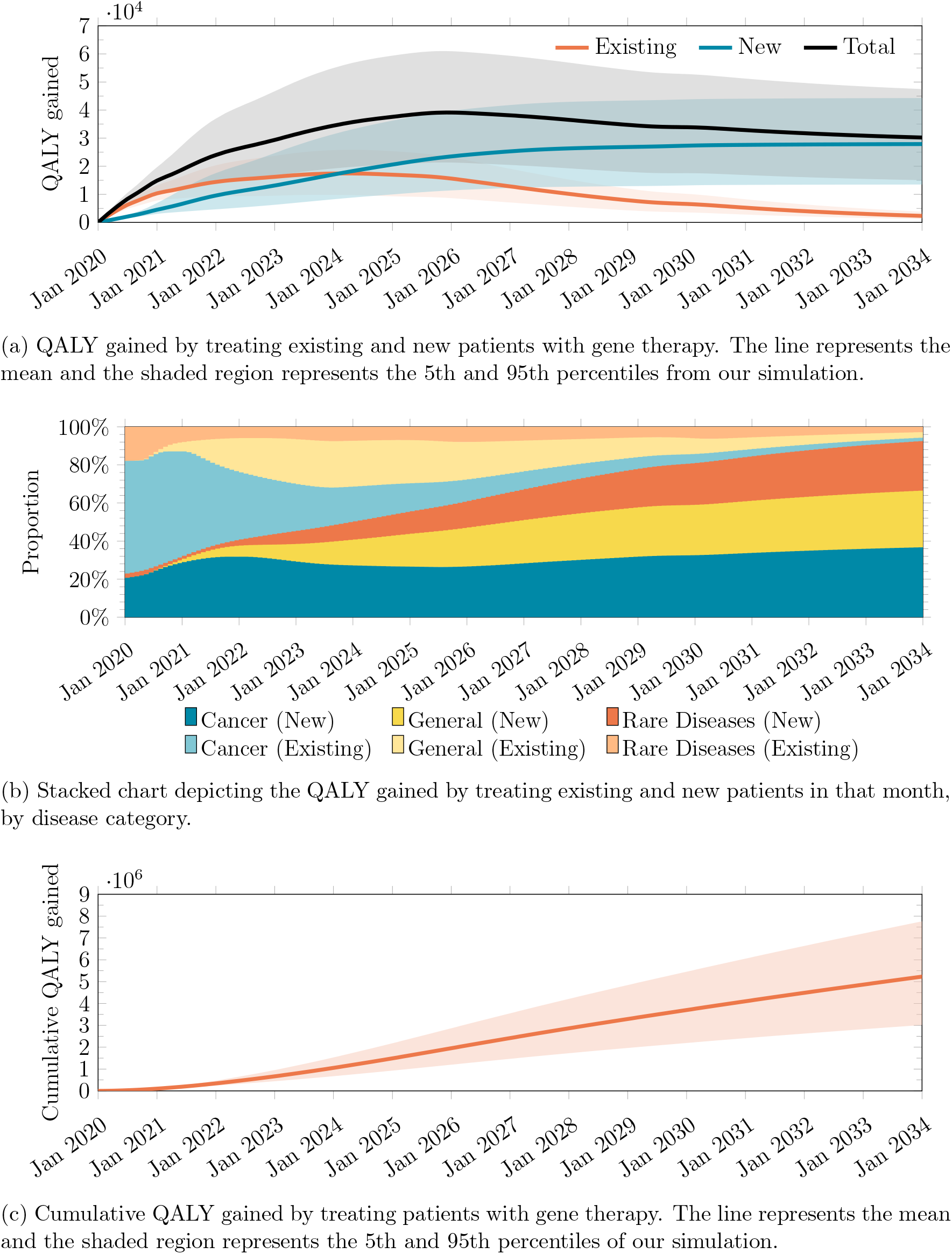
Expected ΔQALY made possible by gene therapy treatments between January 2020 and December 2034, obtained from 1,000,000 simulation runs.

### 4.3 Sensitivity Analysis

To test the sensitivity of our results to initial conditions, we simulate *±*20% changes in the following variables, analyzing their impact on our results.

1. The maximum penetration rate in the ramp function, Θ_*max*_
2. The time to maximum penetration rate in the ramp function, *T*_*max*_
3. The amount of QALY gained in each disease
4. The price per ΔQALY
5. The phase-3-to-approval probability of success (PoS_3*A*_)
6. The number of new patients of each disease
7. The number of existing patients of each disease
8. The time from phase 3 to BLA
9. The time from BLA to approval

For each of these factors, we consider its impact on the the peak monthly spending and the cumulative spending from January 2020 to December 2034 of patient treatment. We also look at how the variables change the timing of the peak monthly spending.

As can be seen from Figure 12, the percentage change in the discounted cumulative spending and the maximum monthly spending on treating all patients with gene therapy scale linearly with the percentage change in several variables: the maximum penetration rate (Θ_*max*_) and the QALY gained (ΔQALY), the price per ΔQALY. Increasing or decreasing the transition probability from phase 3 to approval, or the number of new or existing patients only leads to sublinear increases or decreases in the discounted cumulative spending and the maximum monthly spending. However, changing the time variables, such as the number of days from phase 3 to BLA, from BLA to approval, or the ramp-up period (*T*_*max*_), induce a small change in the opposite direction.

**Figure 12:**
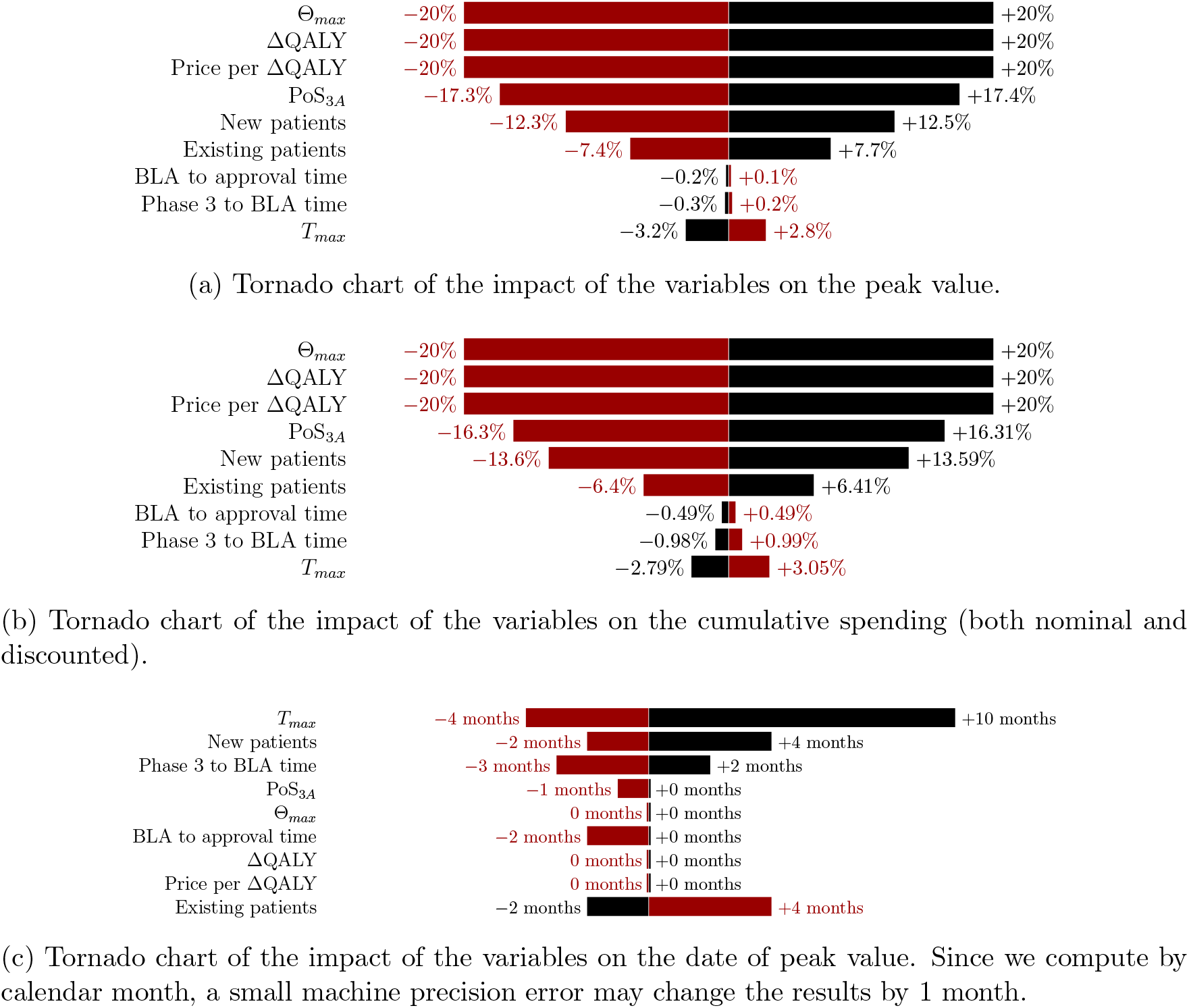
Tornado charts showing the sensitivity of the variables on the different metrics. The black bars represent the effect of increasing the variable by 20% and the red bars represent the effect of decreasing the variable by 20%.

Introducing perturbations of 20% in the probability of success, the number of new patients, the number of days from Phase 3 to BLA or from BLA to approval, or the time to maximum penetration rate in the ramp function (*T*_*max*_) will change the date of the peak monthly spending in the same direction as the perturbation, by up to 10 months. Increasing or decreasing the number of existing patients, on the other hand, will cause a shift of up to 4 months in the date of peak spending in the opposite direction. Perturbing the maximum penetration rate (Θ_*max*_), the QALY gained (ΔQALY), and the price per ΔQALY will not change the date of peak spending.

We also study the effect of changing the correlation between development programs. Changing the correlation from our assumed value of 0.9 to 0 (i.e., perfectly uncorrelated) increases the mean discounted cumulative spending by 3.4%, from $241 billion to $245 billion. Increasing the correlation to 1.0 instead will decrease the mean discounted cumulative spending by 0.4% to $236 billion.

We vary the proportion of existing patients seeking treatment in the first year – which determines the *λ* parameter in Equation 3 – and observe that mean discounted cumulative spending changes by between -32% and +0.08% (see Figure 13). We can expect the results to differ by less than 5% from the baseline if the proportion of existing patients seeking treatments in the first year is between 8% and 45%.

**Figure 13:**
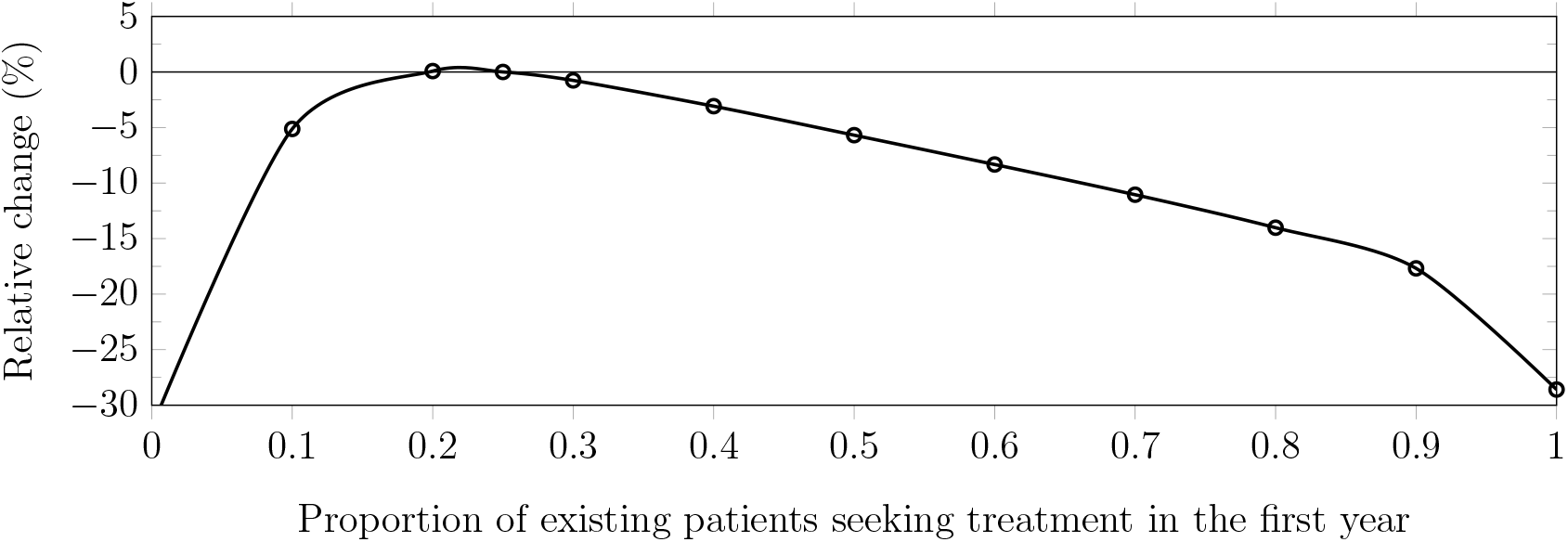
Percentage change in the discounted cumulative spending compared to the baseline when the proportion of existing patients seeking treatment in the first year changes.

Our study is independent of the results by Quinn et al. [142], who estimated that 341,775 patients will have been treated with gene therapy by December 2030, and increases by approximately 50,000 per year in the steady state. The authors of this report did not attempt to quantify the cost of providing gene therapy to these patients. In contrast, our simulation expects that about 820,425 patients will be treated by the end of December 2030, with a steady-state increase of around 61,170 per year in the long run.

Some of the differences between our estimates and this other report are due to differences in sample inclusion criteria and the use of different data for patient prevalence and incidence of disease. For example, Quinn et al. [142] considers “durable” gene therapies under all phases of clinical investigation whereas we consider any therapy with late-stage clinical trial(s). Furthermore, they assume that the ‘potentially treatable pool in oncology is entirely incident—there is no prevalence’. Another difference arises from our decision to start with the broadest range of patients and then deflate these numbers through the penetration rate, rather than attempting to estimate the prevalence and incidence for each patient segment. If we removed existing oncology patients from our simulation, the cumulative number of patients treated by December 2030 becomes 666,895, approximately 1.95 times the estimate in Quinn et al. [142]. We can obtain similar patient estimates to Quinn et al. [142] simply by reducing our penetration rates by 48.8%, which will lower our estimated cumulative spending on gene therapy between January 2020 and December 2034 to $149 billion.

### 4.4 Discussion

We estimate that 1.09 million patients are to be treated with gene therapy by the end of December 2034, spending up to $25.3B annually. These estimates are likely to be lower bounds since our simulation employs conservative assumptions about the speed and volume of gene therapy development. Specifically, we consider only late-stage gene therapy development programs, defined as those already in phase 2/3 or phase 3, and do not account for the possibility that a program in phase 1 or phase 2 may be fast-tracked or granted accelerated approval. We also do not account for any new gene therapies programs being added and approved between 2020 and 2034.

A potential criticism of our approach is that estimating the cost of gene therapies solely based on the change in QALY will overestimate the aggregate spending in the U.S. For example, we do not take into account the potential cost savings to gene therapies due to avoiding multiple costly therapeutic sessions over time based on the current standard of care, or to the recovery of the opportunity cost of caregivers. We have omitted the clinical costs of delivering gene therapy in our analysis, which are often higher than conventional therapeutics due to the need for inpatient hospital care. While there are cases where gene therapy is predicted to provide net cost-savings in treatment after accounting for the direct medical cost (e.g., valoctocogene roxaparvovec for the management of hemophilia A [80]), there is not yet evidence showing that gene therapy will result in net long-term cost savings. In addition, research has shown that new medical technologies generally raise health costs, and that cost-increasing changes in treatments outweigh cost-saving changes the majority of the time [102]. We also do not consider any markup that happens under the prevalent ‘buy-and-bill’ process in the U.S. These considerations suggest that our approach is indeed conservative, and that our estimates are likely to be lower bounds for realized costs over the next 15 years. Nonetheless, we have taken care to calibrate our price per ΔQALY using actual prices for approved therapeutics and estimating QALYs for those diseases, thereby allowing us to produce price estimates that closely track past data.

Another potential criticism is that we fail to consider the possibility that having multiple gene therapies for the same disease may lower the prices of the therapies. However, there is no analogous evidence that the presence of multiple brand-name drugs in the same class lowers the list prices of the drugs [149].

Based on our assumptions, the annual spending on gene therapy will average $20.4 billion and may reach $25.3 billion in 2026. This is well above the $819 million threshold recommended by ICER for its annual potential budget impact [93]. The cumulative spending on all future gene therapies from from January 2020 to January 2034 will be approximately $306 billion, or $241 billion when discounted at a cost of capital of 3% per annum over the next 15 years. We estimate the cost of gene therapy to average $43,110 per unit QALY, several times the average annual expenditure of $16,346 for American cancer patients between 2010 and 2014 [138].

However, when viewed from the broader perspective of aggregate U.S. spending, these figures seem less daunting. In 2018, the U.S. tax revenue was $3.33 trillion, of which individual income tax and payroll tax revenues were $1.68 and $1.17 trillion, respectively [136]. If the average spending of $20.4 billion were to be fully funded through income and payroll taxes, an increase of 0.612% would cover the expense. From a purely budgetary perspective, universal access to gene therapy should be feasible if taxpayers are willing to pay for it. This is probably the simplest and most direct way to provide access to gene therapy to all Americans, though it may not be politically feasible.

Since all elderly patients are covered by Medicare, we estimate that the program would need to increase its annual budget by up to $7.89 billion, or 1.1% of its 2018 spending of

$750.2 billion [19]. Funding this increase would require either an increase in payroll taxes or a reduction in other expenditures.

We estimate that annual gene therapy spending by Medicaid may reach $5.44 billion. This is approximately 0.9% of its 2018 spending of $597.4 billion [19]. Since Medicaid must be provided to all eligible Americans without any preset cap, managing this increase will require either raising funds from state and federal governments to pay for these additional costs, or cutting benefits.

Annual spending by minors and adults who are ineligible for Medicare or Medicaid—and therefore must rely on private insurers—is predicted to reach $12.2 billion. This spending poses a significant challenge for insurers and companies, who face annual budgets and competing priorities. In order to manage spending, many insurance policies might choose not to cover spending on gene therapy, or impose very restrictive policies to limit the number of potential patients who might be treated [156]. Many private insurers are already warning they may not be able or willing to absorb the additional spending should a greater number of people become eligible for expensive gene therapy treatments once new ones reach the market [159].

Many novel methods to finance gene therapy treatments through the existing healthcare infrastructure have been proposed, such as outcome-based payments, whereby the manufacturers would be paid only if the patients achieve predefined outcomes after treatment [72]. We note that both Zolgensma and Luxturna have offered outcome-based payment methods to payers. There have also been proposals to allow mortgage-like payments, and even performance-based annuity payments, as ways to finance gene therapy treatments [133]. In September 2019, Cigna, one of the largest U.S. health insurance companies, announced a program called Embarc Benefit Protection in which employers, health plans, and unions pay a monthly per-member premium that provides members with access to the two FDA-approved gene therapies, Luxturna and Zolgensma, at no out-of-pocket costs to them if their physicians authorize treatment. At the time of writing, Luxturna is not provided under

Embarc Benefit Protection, and it is unclear if the program is in effect. Cigna hopes to keep the monthly cost of the program to below $1 per member [164], but if our simulations are accurate, this will be financially infeasible.

A more ambitious proposal involves creating a national and possibly international gene therapy reinsurance company that performs a similar function to Embarc, but which serves a large number of primary health insurance providers. By allowing multiple primary insurers to cede the specific risk of gene therapy patients to the reinsurer, these risks can be diversified over a much larger pool members, lowering the cost of capital. The capital required for such a reinsurer can be raised through securitization techniques as described in [133], who simulated such a structure, and concluded that the returns to investors would be quite attractive under a broad range of assumptions. However, their simulations were not specifically calibrated for gene therapy, hence our framework may provide a useful complement to their analysis.

Also, it may be more cost-effective for the reinsurer to assume the responsibility of delivering the gene therapies it reinsures through nationally distributed Centers of Excellence (CoEs). This may seem too far afield for a reinsurance company, but the ability to have direct control over the quality of delivery, and to be able to collect data on the performance of these therapies over time, are two compelling reasons for the reinsurer to take this on. The data collected from these centers will be critical, not only for assessing the actuarial risk of reinsurance, but also for implementing performance-related contractual agreements, e.g., if a gene therapy ceases to be effective, then any remaining payments for the therapy will be cancelled.

An additional benefit of a single reinsurer to manage the risk and responsibility of delivering gene therapy is the ability of that reinsurer to avoid the adverse selection problem that often plagues individual insurers [71]. This problem arises when some insurers are willing to pay for gene therapy treatments while others are not, leading patients who require gene therapies to enroll en masse with those insurers providing coverage. Since these policies will likely have higher premiums to cover the high cost of gene therapy, patients have an incentive to leave the policy after receiving the treatment, leaving the insurers to pay the remaining cost without being able to recover the expenses. If a single reinsurer can aggregate this risk across a large pool of gene therapy patients and coordinate payouts across all the insurers, this adverse selection problem can be greatly mitigated, or altogether avoided. The viability of such a reinsurance vehicle would depend critically on the various parameters of the modules in our simulation, as well as the ability to engage with the largest health insurer of all, the U.S. government.

## 5 Conclusion

In this paper, we estimate the number of patients who will be treated by gene therapy between January 2020 and December 2034 using various data sources. We also develop a mathematical model to estimate the cost of these gene therapies, and calibrated the model to yield realistic cost estimates. It is our hope that this study, and our estimates of the potential financial impact of gene therapy in the U.S., will clarify some of the unknowns surrounding the impact of this new class of treatment, and allow policymakers, healthcare providers, insurance companies and patients alike to make more informed financial decisions about the future of this important therapeutic class.

## Data Availability

All data used in the manuscript are either commercial databases licensed by our respective universities or publicly available data. We intend to make our software available with an open-source license.

https://projectalpha.mit.edu

## A1 Current Gene Therapy Clinical Trials

As mentioned in the main paper, we list the clinical trials that are used in this study in the following table.

**Table A1:** List of clinical trials used in this study. ‘TT’ and ‘CT’ indicates ‘TrialTrove’ and ‘*clinicaltrials*.*gov* ’ respectively.

| **Trial Title** | **Disease** | **Sponsors** | **Source** |
| --- | --- | --- | --- |
| Randomized, double-blind, placebo-controlled study of AMG0(HGF plasmid) for patients with arteriosclerosis obliterans | Arteriosclerosis Obliterans | AnGes | TT |
| Tisagenlecleucel Versus Standard of Care in Adult Patients With Relapsed or Refractory Aggressive B-cell Non-Hodgkin Lymphoma: A Randomized, Open Label, Phase III Trial (BELINDA) | B-Cell Non-Hodgkin’s Lymphoma | Novartis | TT |
| A Global Randomized Multicenter Phase III Trial of JCAR017 Compared to Standard of Care in Adult Subjects With High-risk, Second-line, Transplant-eligible Relapsed or Refractory Aggressive B-cell Non-Hodgkin Lymphomas (TRANSFORM). | B-Cell Non-Hodgkin’s Lymphoma | Celgene | TT |
| A Phase III, Open Label Study to Evaluate the Safety and Efficacy of INSTILADRIN (rAd-IFN)/Syn3) Administered Intravesically to Patients With High Grade, BCG Unresponsive Non-Muscle Invasive Bladder Cancer (NMIBC) | BCG Unresponsive NMIBC | FKD Therapeutics | TT |
| A Phase III Study of BC-819 in Patients with Bladder Cancer who Failed Initial Treatment of BCG | BCG Unresponsive NMIBC | Anchiano Therapeutics | TT |
| Continued on next page |  |  |  |

Table A1 – continued from previous page
| **Trial Title** | **Disease** | **Sponsors** | **Source** |
| --- | --- | --- | --- |
| A Phase 3 Single Arm Study Evaluating the Efficacy and Safety of Gene Therapy in Subjects With Transfusion-dependent beta-Thalassemia, Who do Not Have a beta0/beta0 Genotype, by Transplantation of Autologous CD34+ Stem Cells Transduced Ex Vivo With a Lentiviral betaA-T87Q-Globin Vector in Subjects < or = 50 Years of Age | Beta-Thalassemia | bluebird bio | TT |
| A Phase 3 Single Arm Study Evaluating the Efficacy and Safety of Gene Therapy in Subjects With Transfusion-dependent beta-Thalassemia, Who Have a beta0/beta0 Genotype, by Transplantation of Autologous CD34+ Stem Cells Transduced Ex Vivo With a Lentiviral betaA-T87Q-Globin Vector in Subjects < or = 50 Years of Age | Beta-Thalassemia | bluebird bio | TT |
| An Integrated Phase II/III, Open Label, Randomized and Controlled Study of the Safety and Efficacy of CG0070 Adenovirus Vector Expressing GM-CSF in Patients With Non-Muscle Invasive Bladder Cancer With Carcinoma In Situ Disease Who Have Failed BCG Bladder Oncolytic virus for Non-muscle invasive bladder cancer Disease (BOND) | Bladder Cancer, in situ concurrent with Papillary Tumors | Cold Genesys | TT |
| A Phase 2/3 Study of the Efficacy and Safety of Hematopoietic Stem Cells Transduced With Lenti-D Lentiviral Vector for the Treatment of Cerebral Adrenoleukodystrophy (CALD) | Cerebral Adrenoleukodystrophy (CALD) | bluebird bio | CT |
| Efficacy and Safety of AAV2-REP1 for the Treatment of Choroideremia | Choroideremia | Nightstar Therapeutics | CT |

Table A1 – continued from previous page
| **Trial Title** | **Disease** | **Sponsors** | **Source** |
| --- | --- | --- | --- |
| A Phase 3 Double-Blind, Randomized, Placebo-Controlled Study to Evaluate the Safety and Efficacy of AMG0 in Subjects With Critical Limb Ischemia Efficacy and Safety of AMG0 in Subjects With Critical Limb Ischemia (AGILITY) | Critical Limb Ischemia | AnGes | TT |
| Safety and Efficacy of Recombinant Adeno-Associated Virus Containing the CFTR Gene in the Treatment of Cystic Fibrosis | Cystic Fibrosis | Targeted Genetics Corporation/ Cystic Fibrosis Foundation Therapeutics | CT |
| A Placebo Controlled, Double-blind, Randomized, Parallel-group, Multi-center Phase III study to determine the Efficacy and Safety of TissueGene-C in Patients with Degenerative Arthritis | Degenerative Arthritis | Kolon Life Science | TT |
| Safety and Efficacy Study of Pl-VEGF165 to Treat Diabetic Foot Syndrome | Diabetic Foot Syndrome | Human Stem Cells Institute | TT |
| A Phase III, Double-blind, Randomized, Placebo-controlled, Multicenter Study to Assess the Safety and Efficacy of VM202 to Treat Chronic Nonhealing Foot Ulcers in Diabetic Patients With Concomitant Peripheral Arterial Disease (PAD) | Diabetic Foot Ulcers | Helixmith | TT |
| A Phase III, Double-Blind, Randomized, Placebo-Controlled, Multicenter Study to Assess the Safety and Efficacy of VM202 in Subjects With Painful Diabetic Peripheral Neuropathy | Diabetic Peripheral Neuropathy | Helixmith | TT |
| A Phase III, Randomized, Open-Label Study Evaluating Efficacy of Axicabtagene Ciloleucel Versus Standard of Care Therapy in Subjects With Relapsed/Refractory Diffuse Large B Cell Lymphoma | Diffuse Large B Cell Lymphoma (DLBCL) | Gilead Sciences/Kite Pharma | TT |
| Continued on next page |  |  |  |

Table A1 – continued from previous page
| Trial Title | Disease | Sponsors | Source |
| --- | --- | --- | --- |
| A Multi-center Phase III, Randomized, Open-Label Trial of Vigil (Bi-shRNAfurin and GMCSF Augmented Autologous Tumor Cell Immunotherapy) in Combination With Irinotecan and Temozolomide as a Second-Line Regimen for Ewing's Sarcoma | Ewing's Sarcoma | Gradalis | TT |
| A Phase III Study of INGN 241 in Combination with Radiation Therapy in Patients with Advanced Solid Tumors and Head and neck cancer. | Head and Neck Cancer | Introgen Therapeutics | TT |
| An Open-Label, Randomized, Multi-Center Phase III Clinical Trial Comparing E10A Plus Chemotherapy And Chemotherapy Alone For Treatment Of Head And Neck Cancer | Head and Neck Cancer | Marsala Biotech | TT |
| A Randomized, Open-label, Multi-center Phase III Study Designed to Evaluate the Safety and Efficacy of E10A in Patients With Recurrent/Unresectable Squamous Cell Carcinoma of the Head and Neck Region | Head and Neck Cancer | Guangzhou Double Bioproducts Co. | TT |
| A Phase III, Pivotal, Randomized, Placebo-controlled, Double-Blind, Multicenter Study to Evaluate RT-100 AC6 Gene Transfer in Patients with Heart Failure and Reduced Left Ventricular Ejection Fraction;Heart Failure with Reduced Left Ventricular Ejection Fraction: One-time Gene Transfer Using RT-100 Intracoronary Administration of Adenovirus 5 encoding Human AC6 (FLOURISH) | Heart Failure | Renova Therapeutics | TT |
| Continued on next page |  |  |  |

Table A1 – continued from previous page
| **Trial Title** | **Disease** | **Sponsors** | **Source** |
| --- | --- | --- | --- |
| A Phase 3 Open-Label, Single-Arm Study To Evaluate The Efficacy and Safety of BMN 270, an Adeno-Associated Virus Vector-Mediated Gene Transfer of Human Factor VIII in Hemophilia A Patients With Residual FVIII Levels = 1 IU/dL Receiving Prophylactic FVIII Infusions | Hemophilia A | BioMarin | TT |
| Phase 3 Study To Evaluate Efficacy/Safety of Valoctocogene Roxaparvovec an AAV Vector-Mediated Gene Transfer of hFVIII at a Dose of 4E13vg/kg in Hemophilia A Patients With Residual FVIII Levels < or = 1IU/dL Receiving Prophylactic FVIII Infusions | Hemophilia A | BioMarin | TT |
| A Phase III Run In trial to Evaluate SPK-8011 in Patients with Hemophilia A | Hemophilia A | Roche/Spark Therapeutics | TT |
| An open-label, single-dose, multi-center, multi-national, Phase III pivotal trial to investigate efficacy and safety of AMT-061 in severe or moderately severe hemophilia B; HOPE-B: Trial of AMT-061 in Severe or Moderately Severe Hemophilia B Patients; Phase III, Open-label, Single-dose, Multi-center, Multinational Trial Investigating a Serotype 5 Adeno-associated Viral Vector Containing the Padua Variant of a Codon-optimized Human Factor IX Gene (AAV5-hFIXco-Padua, AMT-061) Administered to Adult Subjects With Severe or Moderately Severe Hemophilia B | Hemophilia B | uniQure | TT |
| Continued on next page |  |  |  |

Table A1 – continued from previous page
| Trial Title | Disease | Sponsors | Source |
| --- | --- | --- | --- |
| A Pivotal Phase III Study of PF-06838435 in Patients with Hemophilia B; Phase 3, Open Label, Single Arm Study To Evaluate Efficacy And Safety Of Fix Gene Transfer With Pf-06838435 (Raav-Spark100-Hfix-Padua) In Adult Male Participants With Moderately Severe To Severe Hemophilia B (Fix:C < or =2%) | Hemophilia B | Pfizer | TT |
| A Pivotal Phase III Study to Evalaute AMT-060 in Patients with Hemophilia B | Hemophilia B | uniQure | TT |
| Multicenter Randomized Controlled Trial of Adenovirus-mediated Adjuvant Gene Therapy Improving Outcome of Liver Transplantation in Patients With Advanced Hepatocellular Carcinoma | Hepatocellular Carcinoma | Wuhan Tiandakang Bio-Tech Engineering Co./ Shenzhen Tiandakang Gene Engineering Co. | TT |
| A Phase III Randomized, Open-Label Study Comparing Pexa Vec (Vaccinia GM CSF / Thymidine Kinase-Deactivated Virus) Followed by Sorafenib Versus Sorafenib in Patients With Advanced Hepatocellular Carcinoma (HCC) Without Prior Systemic Therapy | Hepatocellular Carcinoma | Transgene/ Sillajen Biotherapeutics /Jennerex/ Lees Pharmaceutical | TT |
| Phase III Prospective, Open-Label, Parallel-Group, Randomized, Multicenter Trial Comparing the Efficacy of Surgery, Radiation, and Injection of Murine Cells Producing Herpes Simplex Thymidine Kinase Vector Followed by Intravenous Ganciclovir Against the Efficacy of Surgery and Radiation in the Treatment of Newly Diagnosed, Previously Untreated Glioblastoma Multiforme | High-Grade Glioma | Novartis/Sandoz | TT |

Table A1 – continued from previous page
| **Trial Title** | **Disease** | **Sponsors** | **Source** |
| --- | --- | --- | --- |
| A Controlled, Randomised, Parallel Group Study Of The Efficacy And Safety Of Herpes Simplex Virus Thymidine Kinase Gene Therapy (Cerepro) with Subsequent Ganciclovir For The Treatment Of Patients With Operable High-Grade Glioma. | High-Grade Glioma | Trizell | TT |
| A Randomized, Double-Blind, Placebo-Controlled, Multi-Center, Phase 3 Study to Determine the Efficacy of TG-C in Subjects With Kellgren and Lawrence Grade (KLG) 2 or 3 Osteoarthritis of the Knee | Knee Osteoarthritis with Kellgren & Lawrence Grade 2 or 3 | Kolon TissueGene | TT |
| A Multicenter, Randomized, Placebo Controlled, Double-blind, Parallel, Phase III Clinical Trial to Evaluate the Efficacy and Safety of Invossa K Injection in Patients Diagnosed as Knee Osteoarthritis With Kellgren & Lawrence Grade 2 | Knee Osteoarthritis with Kellgren & Lawrence Grade 2 or 3 | Kolon Life Science | TT |
| Safety and Efficacy Study in Subjects With Leber Congenital Amaurosis | Leber Congenital Amaurosis due to RPE65 Mutations | Spark Therapeutics | CT |
| Efficacy Study of GS010 for Treatment of Vision Loss From 7 Months to 1 Year From Onset in LHON Due to the ND4 Mutation | Leber Hereditary Optic Neuropathy | GenSight Biologics | CT |
| Efficacy Study of GS010 for the Treatment of Vision Loss up to 6 Months From Onset in LHON Due to the ND4 Mutation | Leber Hereditary Optic Neuropathy | GenSight Biologics | CT |
| Efficacy and Safety Study of Bilateral Intravitreal Injection of GS010 for the Treatment of Vision Loss up to 1 Year From Onset in LHON Due to the ND4 Mutation | Leber Hereditary Optic Neuropathy | GenSight Biologics | CT |
| Continued on next page |  |  |  |

Table A1 – continued from previous page
| **Trial Title** | **Disease** | **Sponsors** | **Source** |
| --- | --- | --- | --- |
| Tisagenlecleucel Versus Blinatumomab or Inotuzumab for Adult Patients With Relapsed/Refractory B-cell Precursor Acute Lymphoblastic Leukemia: A Randomized Open Label, Multicenter, Phase III Trial | Leukemia (Acute Lymphoblastic) | Novartis | TT |
| Phase IIIb Study for Relapsed/Refractory Pediatric/Young Adult Acute Lymphoblastic Leukemia Patients to be Treated With CTL019 | Leukemia (Acute Lymphoblastic) | Novartis | TT |
| A Phase II/III Prospective, Open Label Study to Evaluate Safety and Efficacy of Intravenous Autologous CD19 CAR-T Cells for Relapsed/ Refractory B-Acute Lymphoblastic Leukemia | Leukemia (Acute Lymphoblastic) | Gaia Science | TT |
| A Randomized Phase II/III Study of $\alpha\beta$ T Cell-Depleted, Related, Haploidentical Hematopoietic Stem Cell Transplant (Haplo-HSCT) Plus Rivogenlecleucel vs. Haplo-HSCT Plus Post-Transplant Cyclophosphamide (PTCy) in Patients With AML or MDS | Leukemia (Acute Myelogenous) | Bellicum Pharmaceuticals | TT |
| Randomized, Registrational, Controlled Study of BPX-501 with Allogeneic Hematopoietic Stem Cells (Allo-HSCT) in Patients with Acute Myelogenous Leukemia | Leukemia (Acute Myelogenous) | Bellicum Pharmaceuticals | TT |
| TK008: Randomized Phase III Trial of Haploidentical HCT With or Without an Add Back Strategy of HSV-Tk Donor Lymphocytes in Patients With High Risk Acute Leukemia | Leukemia (Acute Myelogenous) | Molmed | TT |
| A Phase IIb/III Study of AST-VAC1 in Patients with Acute Myelogenous Leukemia (AML) | Leukemia (Acute Myelogenous) | Asterias/Lineage Cell Therapeutics | TT |
| A Study of Glybera for the Treatment of Lipoprotein Lipase (LPL) Deficiency | Lipoprotein Lipase Deficiency (LPLD) | uniQure | TT |

Table A1 – continued from previous page
| **Trial Title** | **Disease** | **Sponsors** | **Source** |
| --- | --- | --- | --- |
| A Study to Determine the Safety and Efficacy in Lipoprotein Lipase-Deficient Subjects After Intramuscular Administration of AMT-011, an Adeno-Associated Viral Vector Expressing Human Lipoprotein LipaseS447X. | Lipoprotein Lipase Deficiency (LPLD) | uniQure | TT |
| An Open-label Study to Assess the Efficacy and Safety of Alipogene Tiparvovec (AMT-011), Human LPL [S447X], Expressed by an Adeno-Associated Viral Vector After Intramuscular Administration in LPL-deficient Adult Subjects | Lipoprotein Lipase Deficiency (LPLD) | uniQure | TT |
| A Study of AMT-011 in Patients With LPL Deficiency | Lipoprotein Lipase Deficiency (LPLD) | uniQure | TT |
| A Phase III Trial of Glybera for Dyslipidemia | Lipoprotein Lipase Deficiency (LPLD) | uniQure | TT |
| Phase II/III study of Ad-IFNγ in Cutaneous T-cell lymphoma | Lymphoma | Transgene | TT |
| A Safety and Efficacy Study of Cryopreserved GSK2696274 for Treatment of Metachromatic Leukodystrophy (MLD) | Metachromatic Leukodystrophy | GlaxoSmithKline | CT |
| PV-10 Intralesional Injection vs Systemic Chemotherapy or Oncolytic Viral Therapy for Treatment of Locally Advanced Cutaneous Melanoma | Melanoma (Locally Advanced Cutaneous) | Provectus<br>Biopharmaceuticals | TT |
| A Phase Ib/III, Multicenter, Trial of Talimogene Laherparepvec in Combination With Pembrolizumab (MK-3475) for Treatment of Unresectable Stage IIIB to IVM1c Melanoma (MASTERKEY-265/KEYNOTE-034) | Melanoma (Metastatic) | Amgen/ Merck & Co./Merck Sharp & Dohme (MSD) | TT |
| Continued on next page |  |  |  |

Table A1 – continued from previous page
| **Trial Title** | **Disease** | **Sponsors** | **Source** |
| --- | --- | --- | --- |
| A Phase III Clinical Trial to Evaluate the Safety and Efficacy of Treatment With 2 mg Intralesional Allovectin-7 Compared to Dacarbazine (DTIC) or Temozolomide (TMZ) in Subjects With Recurrent Metastatic Melanoma;Allovectin-7 Immunotherapeutic for Metastatic Melanoma (AIMM). | Melanoma (Metastatic) | Brickell Biotech, AnGes | TT |
| A Randomized Phase III Clinical Trial to Evaluate the Efficacy and Safety of Treatment With OncoVEXGM-CSF Compared to Subcutaneously Administered GM-CSF in Melanoma Patients With Unresectable Stage IIIB, IIIC and IV Disease | Melanoma (Metastatic) | Amgen | TT |
| An Extension Protocol to Evaluate the Efficacy and Safety of Extended Use Treatment With OncoVEXGM-CSF for Eligible Melanoma Patients Participating in Study 005/05 | Melanoma (Metastatic) | Amgen | TT |
| A Controlled, Randomized Phase III Trial Comparing the Response to Dacarbazine With and Without Allovectin-7 in Patients With Metastatic Melanoma. | Melanoma (Metastatic) | Brickell Biotech | TT |
| Open-label, Single-arm, Multi-center Study of Intracerebral Administration of Adeno-associated Viral (AAV) Serotype rh.10 Carrying Human N-sulfoglucosamine Sulfohydrolase (SGSH) cDNA for Treatment of Mucopolysaccharidosis Type IIIA | Mucopolysaccharidosis Type IIIa | LYSOGENE | TT |
| Continued on next page |  |  |  |

Table A1 – continued from previous page
| **Trial Title** | **Disease** | **Sponsors** | **Source** |
| --- | --- | --- | --- |
| A Phase III, Single Arm, Multi-Center Study to Assess the Efficacy and Safety of Clarithromycin(Biaxin)-Lenalidomide-Low-Dose-Dexamethasone (BiRd) Combined With B-cell Muturation Antigen (BCMA)-Directed Chimeric Antigen Receptor (CAR) T-cell Therapy in Patients With Newly Diagnosed Multiple Myeloma | Multiple Myeloma (Newly Diagnosed) | Shanghai Unicar-Therapy Bio-medicine | TT |
| Clinical Trial of Recombinant Adenovirus-p53 (Gen-dicine) Combined with Radiotherapy in Nasopharyngeal Carcinoma Patients.; | Nasopharyngeal Carcinoma | Shenzhen SiBiono GeneTech Co. | TT |
| A Phase II/III, Multi-Center, Open-Label, Randomized Study to Compare the Effectiveness and Safety of Intralesional Administration of RPR/INGN 201 in Combination with Taxotere and Carboplatin and Radiotherapy Versus Taxotere and Carboplatin and Radiotherapy Alone in Patients with Locally Advanced Unresectable Non-Small Cell Lung Cancer (NSCLC) | NSCLC | Introgen Therapeutics | TT |
| A Phase IIB/III Randomized, Double-blind, Placebo Controlled Study Comparing First Line Therapy With or Without TG4010 Immunotherapy Product in Patients With Stage IV Non-Small Cell Lung Cancer (NSCLC) | NSCLC | Transgene | TT |
| Phase III multi-center, open, randomized clinical trial of percutaneous intratumoral injection of genetically engineered adenovirus (injection of H101), IL-2, TB hyperthermia and systemic chemotherapy in the treatment of advanced non-small cell lung cancer | NSCLC | Shanghai Sunway Biotech | TT |

Table A1 – continued from previous page
| **Trial Title** | **Disease** | **Sponsors** | **Source** |
| --- | --- | --- | --- |
| Phase III Study of Lucanix (Belagenpumatucel-L) in Advanced Non-small Cell Lung Cancer: An International Multicenter, Randomized, Double-blinded, Placebo-controlled Study of Lucanix Maintenance Therapy for Stages III/IV NSCLC Subjects Who Have Responded to or Have Stable Disease Following One Regimen of Front-line, Platinum-based Combination Chemotherapy; Survival, Tumor-free, Overall and Progression-free (STOP) | NSCLC Stage 3 | Activate Immunotherapy | TT |
| rAd-p53 Combined Chemotherapy Via Selective Arterial Cannula in The Treatment of Advanced Oral Cancer, A Randomized Controlled Trial | Oral Cancer (Advanced) | Shenzhen SiBiono GeneTech Co. | TT |
| A Randomized, Controlled, Double-Arm, Open-Label, Multi-Center Study of Ofranergene Obadenovec (VB-111) Combined With Paclitaxel vs. Paclitaxel Monotherapy for the Treatment of Recurrent Platinum-Resistant Ovarian Cancer | Ovarian Cancer (Platinum-Resistant) | Gynecologic Oncology Group (GOG)/ VBL Therapeutics | TT |
| A Phase II/III Trial of Chemotherapy Alone Versus Chemotherapy Plus SCH 58500 in Newly Diagnosed Stage III Ovarian and Primary Peritoneal Cancer Patients With Greater Than or Equal to 0.5 cm and Less Than or Equal to 2 cm Residual Disease Following Surgery | Ovarian Cancer, Primary Peritoneal Cavity Cancer | Merck & Co./Merck Sharp & Dohme (MSD) | TT |
| A Randomized, Phase II/III, Study of TNFerade Biologic With 5-FU and Radiation Therapy for First-Line Treatment of Unresectable Locally Advanced Pancreatic Cancer | Pancreatic Cancer (Locally Advanced) | Precigen | TT |

Table A1 – continued from previous page
| **Trial Title** | **Disease** | **Sponsors** | **Source** |
| --- | --- | --- | --- |
| Phase II/III Study of ProSavin for the Treatment of Parkinson's Disease | Parkinson's Disease | Oxford BioMedica | TT |
| Phase III Trial of CERE-120 for Parkinson's Disease | Parkinson's Disease | Sanofi/Sanofi Genzyme,<br>Sangamo Therapeutics | TT |
| A Randomized, Placebo-controlled Phase IIIa Pivotal Confirmatory Study to Evaluate Safety and Efficacy of VY-AADC in Patients with Parkinson's Disease | Parkinson's Disease | Neurocrine Biosciences | TT |
| A Randomized Double-Blind Placebo-Controlled Parallel Group Study of the Efficacy and Safety of XRP0038/NV1FGF on Amputation or Any Death in Critical Limb Ischemia Patients With Skin Lesions | Peripheral Artery Disease | Sanofi | TT |
| Efficiency, Safety and Portability of Neovasculgen | Peripheral Artery Disease | Human Stem Cell Institute, Russia | CT |
| Gene Therapy using Intramuscular Administration of AMG0001 in Patients with Peripheral Arterial Disease; | Peripheral Artery Disease | AnGes | TT |
| Hepatocyte Growth Factor to Improve Functioning in Peripheral Artery Disease: The HI-PAD Study; | Peripheral Artery Disease | Helixmith | TT |
| A phase III study of HGF Plasmid in Peripheral Arterial Disease (PAD) in the US | Peripheral Artery Disease | AnGes | TT |
| Phase 3 Study of Efficiency, Safety and Portability of Gene Therapy Drug Neovasculgen (DNA Encoding the 165-amino-acid Isoform of Human Vascular Endothelial Growth Factor (pCMV - VEGF165) for Peripheral Arterial Disease Complex Treatment | Peripheral Artery Disease | Human Stem Cells Institute | TT |
| Provenge (Sipuleucel-T) Active Cellular Immunotherapy Treatment of Metastatic Prostate Cancer After Failing Hormone Therapy | Prostate Cancer | Dendreon | CT |

Table A1 – continued from previous page
| Trial Title | Disease | Sponsors | Source |
| --- | --- | --- | --- |
| A Randomized, Controlled Trial of Replication-Competent Adenovirus-Mediated Suicide Gene Therapy in Combination With IMRT Versus IMRT Alone for the Treatment of Newly-Diagnosed Prostate Cancer With an Intermediate Risk Profile | Prostate Cancer (Localized) | Henry Ford Health System | TT |
| A Randomized Controlled Trial of ProstAtak as Adjuvant to Up-front Radiation Therapy For Localized Prostate Cancer | Prostate Cancer (Localized) | Candel Therapeutics | TT |
| A Phase III Randomized, Open-Label Study of CG1940 and CG8711 Versus Docetaxel and Estramustine in Patients with Metastatic Hormone-Refractory Prostate Cancer Who are Chemotherapy-Naive. | Prostate Cancer (Metastatic Hormone-Refractory) | ANI Pharmaceuticals, Takeda | TT |
| A Phase III Randomized, Open-Label Study of CG1940 and CG8711 Versus Docetaxel and Prednisone in Patients With Metastatic Hormone-Refractory Prostate Cancer Who Are Chemotherapy-Naive. | Prostate Cancer (Metastatic Hormone-Refractory) | ANI Pharmaceuticals, Takeda | TT |
| A Phase III Randomized, Open-Label Study of Docetaxel in Combination With CG1940 and CG8711 Versus Docetaxel and Prednisone in Taxane-Nave Patients With Metastatic Hormone-Refractory Prostate Cancer With Pain. | Prostate Cancer (Metastatic Hormone-Refractory) | ANI Pharmaceuticals, Takeda | TT |
| A Randomized Controlled Trial Of AdV-tk + Valacyclovir Administered During Active Surveillance For Newly Diagnosed Prostate Cancer | Prostate Cancer (Newly Diagnosed) | Candel Therapeutics | TT |
| An Open label,Randomized, Multi-Centered, Intra-Patient Controlled Phase III Study of FCX-007 in Patients with Recessive Dystrophic Epidermolysis Bullosa (RDEB) | Recessive Dystrophic Epidermolysis Bullosa | Fibrocell Science | TT |
| Continued on next page |  |  |  |

Table A1 – continued from previous page
| **Trial Title** | **Disease** | **Sponsors** | **Source** |
| --- | --- | --- | --- |
| VITAL: A Pivotal Phase 3 Study of EB-101 for the Treatment of Recessive Dystrophic Epidermolysis Bullosa (RDEB) (GENE TRANSFER) | Recessive Dystrophic Epidermolysis Bullosa | Stanford University Medical Center/ Abeona Therapeutics | TT |
| A Phase III, Randomized, Controlled, Double-Arm, Open-Label, Multi-center Study of VB-111 Combined With Bevacizumab vs. Bevacizumab Monotherapy in Patients With Recurrent Glioblastoma | Recurrent Glioblastoma | VBL Therapeutics | TT |
| A Phase II/III Randomized, Open-Label Study of Toca 511, a Retroviral Replicating Vector, Combined With Toca FC Versus Standard of Care in Subjects Undergoing Planned Resection for Recurrent Glioblastoma or Anaplastic Astrocytoma | Recurrent Glioblastoma | Tocagen | TT |
| A Randomized, Double-Blind, Placebo-Controlled, Parallel Group, Multicenter, Phase 3 Study to Evaluate the Safety and Efficacy of Ad5FGF-4 in Patients With Refractory Angina Due to Myocardial Ischemia; Ad5FGF-4 In Patients With Refractory Angina Due to Myocardial Ischemia (AFFIRM) | Refractory Angina due to Myocardial Ischemia (AFFIRM) | Gene Biotherapeutics/Angionetics | TT |
| A Phase III, Multicenter, Randomized, Open-label Study to Compare the Efficacy and Safety of bb2121 Versus Standard Triplet Regimens in Subjects With Relapsed and Refractory Multiple Myeloma (RRMM) (KarMMa-3) | Relapsed and Refractory Multiple Myeloma (RRMM) | Celgene | TT |
| A Single Global Phase 3 trial of RST-001 in Patients With Retinitis Pigmentosa (RP) | Retinitis Pigmentosa | Abbvie/Allergan | TT |
| A Phase II/III Expansion Study to Evaluate Safety and Efficacy of NSR-RPGR in Patients with a Diagnosis of X - Linked Retinitis Pigmentosa due to RPGR mutations | Retinitis Pigmentosa | NightstaRx | TT |
| Continued on next page |  |  |  |

Table A1 – continued from previous page
| **Trial Title** | **Disease** | **Sponsors** | **Source** |
| --- | --- | --- | --- |
| Phase 3 HGB-210 study of LentiGlobin in patients with SCD | Sickle Cell Anemia | bluebird bio | TT |
| Open-label, historical controlled study of AVXS-101 for treatment of spinal muscular atrophy | Spinal Muscular Atrophy | Novartis/AveXis | TT |
| A Multi-National Study of a One-Time Intrathecal Dose of AVXS-101 in Patients with Spinal Muscular Atrophy Types 1, 2, 3 | Spinal Muscular Atrophy | Novartis/AveXis | TT |
| A Global Study of a Single, One-Time Dose of AVXS-101 Delivered to Infants With Genetically Diagnosed and Pre-symptomatic Spinal Muscular Atrophy With Multiple Copies of SMN2 | Spinal Muscular Atrophy Type 1 | Novartis/AveXis | TT |
| European, Phase 3, Open-Label, Single-Arm, Single-Dose Gene Replacement Therapy Clinical Trial for Patients With Spinal Muscular Atrophy Type 1 With One or Two SMN2 Copies Delivering AVXS-101 by Intravenous Infusion | Spinal Muscular Atrophy Type 1 | Novartis/AveXis | TT |
| Phase 3, Open-Label, Single-Arm, Single-Dose Gene Replacement Therapy Clinical Trial for Patients With Spinal Muscular Atrophy Type 1 With One or Two SMN2 Copies Delivering AVXS-101 by Intravenous Infusion | Spinal Muscular Atrophy Type 1 | Novartis/AveXis | TT |
| A Phase Ib/III Multicenter, Randomized, Trial of Talimogene Laherparepvec in Combination With Pembrolizumab for the Treatment of Subjects With Recurrent or Metastatic Squamous Cell Carcinoma of the Head and Neck | Squamous Cell Cancer of Head and Neck or Esophagus | Amgen/ Merck & Co./Merck Sharp & Dohme (MSD) | TT |

Table A1 – continued from previous page
| **Trial Title** | **Disease** | **Sponsors** | **Source** |
| --- | --- | --- | --- |
| Phase III randomized clinical trial of intratumoral injection of E1B gene-deleted adenovirus (H101) combined with cisplatin-based chemotherapy in treating squamous cell cancer of head and neck or esophagus. | Squamous Cell Cancer of Head and Neck or Esophagus | Shanghai Sunway Biotech | TT |
| Phase III Randomized Study of Ad5CMV-p53 Gene Therapy (INGN 201) Versus Methotrexate in Patients With Refractory Squamous Cell Carcinoma of the Head and Neck (T301). | Squamous Cell Cancer of Head and Neck or Esophagus | Sanofi, Introgen Therapeutics | TT |
| A Phase III, Multi-Center, Open-Label, Randomized Study to Compare the Effectiveness and Safety of Intratumoral Administration of INGN 201 in Combination with Chemotherapy Versus Chemotherapy Alone in Patients with Squamous Cell Carcinoma of the Head and Neck (SCCHN) | Squamous Cell Cancer of Head and Neck or Esophagus | Introgen Therapeutics | TT |
| A Randomized, Controlled, Parallel Group, Multicenter Phase 3 Study to Evaluate the Efficacy and Safety of Ad5FGF-4 Using SPECT Myocardial Perfusion Imaging in Patients With Stable Angina Pectoris | Stable Angina | Gene Biotherapeutics/ Angionetics/ Gene Biotherapeutics | TT |
| A Randomized, Double Blind, Placebo Controlled, Parallel Group, Multicenter Study to Evaluate the Efficacy and Safety of Ad5FGF-4 in Female Patients With Stable Angina Pectoris Who Are Not Candidates for Revascularization; Angiogenesis in Women with Angina pectoris who are not candidates for Revascularization [AWARE] | Stable Angina | Gene Biotherapeutics/ Angionetics/ Gene Biotherapeutics | TT |
| Continued on next page |  |  |  |

Table A1 – continued from previous page
| **Trial Title** | **Disease** | **Sponsors** | **Source** |
| --- | --- | --- | --- |
| A Multinational Multicenter, Randomized, Double Blind, Placebo Controlled Study to Evaluate the Efficacy and Safety of Ad5FGF-4 in Patients With Stable Angina;(The Angiogenic Gene Therapy Trial - 4 [AGENT 4]). | Stable Angina | Bayer AG/Bayer HealthCare, Gene Biotherapeutics | TT |
| A Multicenter, Randomized, Double-Blind, Placebo Controlled Study to Evaluate the Efficacy and Safety of Ad5FGF-4 in Patients With Stable Angina (The Angiogenic Gene Therapy Trial - 3 [AGENT 3]) | Stable Angina | Bayer AG/Bayer HealthCare, Gene Biotherapeutics | TT |
| Multicentre, Randomized,Double Blind, Placebo Controlled Trial of Myocardial Angiogenesis Using VEGF165, Intramyocardial Gene Delivery in Patients With Severe Angina Pectoris | Stable Angina | Johnson & Johnson | TT |
| A Pivotal Study of NY-ESO-1 in Patients with Synovial Sarcoma including Myxoid Round Cell Liposarcoma | Synovial Sarcoma | GlaxoSmithKline/AdaptImmune | TT |

## A2 Disease-to-Therapeutic Area Mapping

As mentioned in the main paper, we show how the diseases are related to the therapeutic areas in the table below.

**Table A2:**
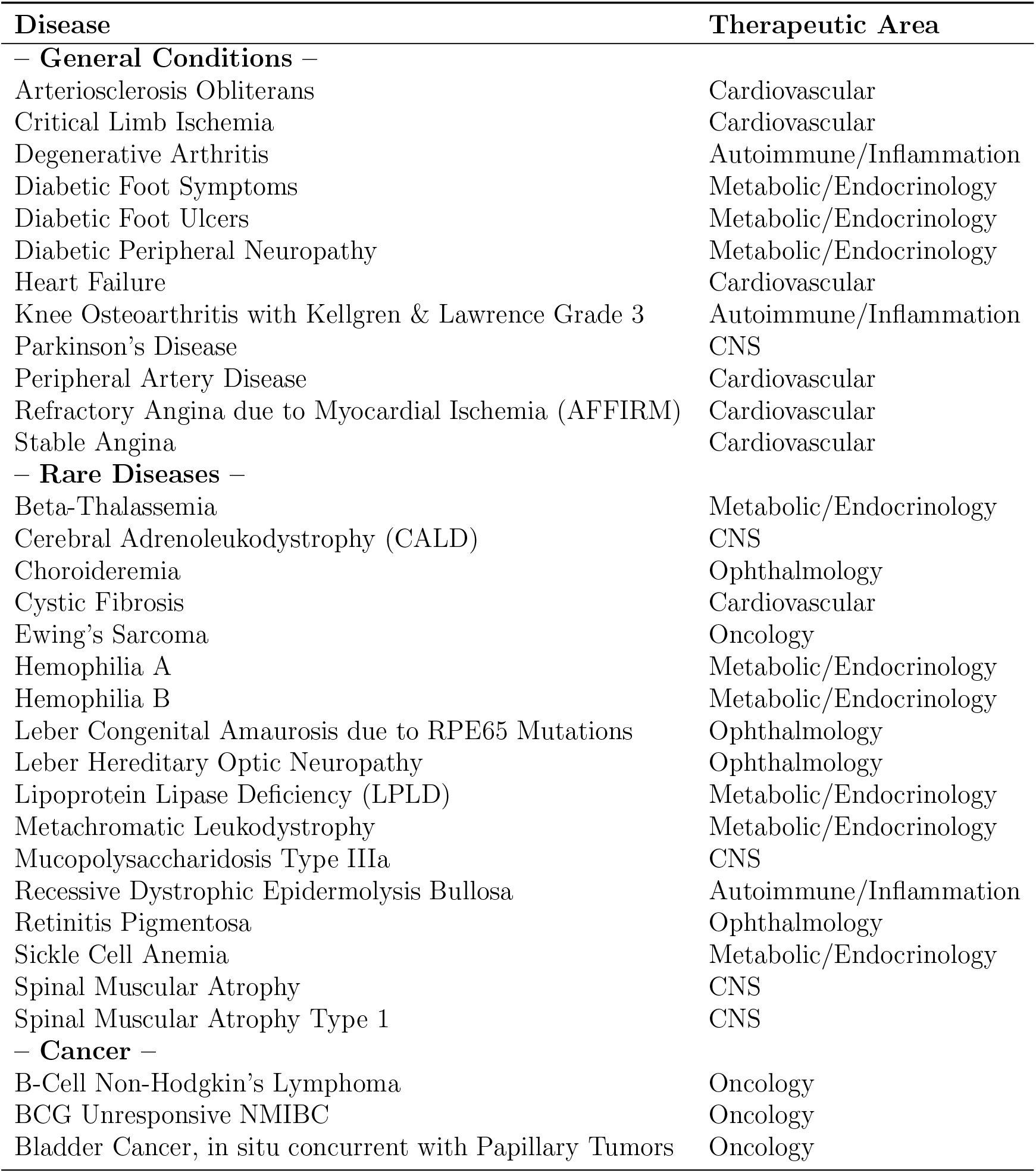
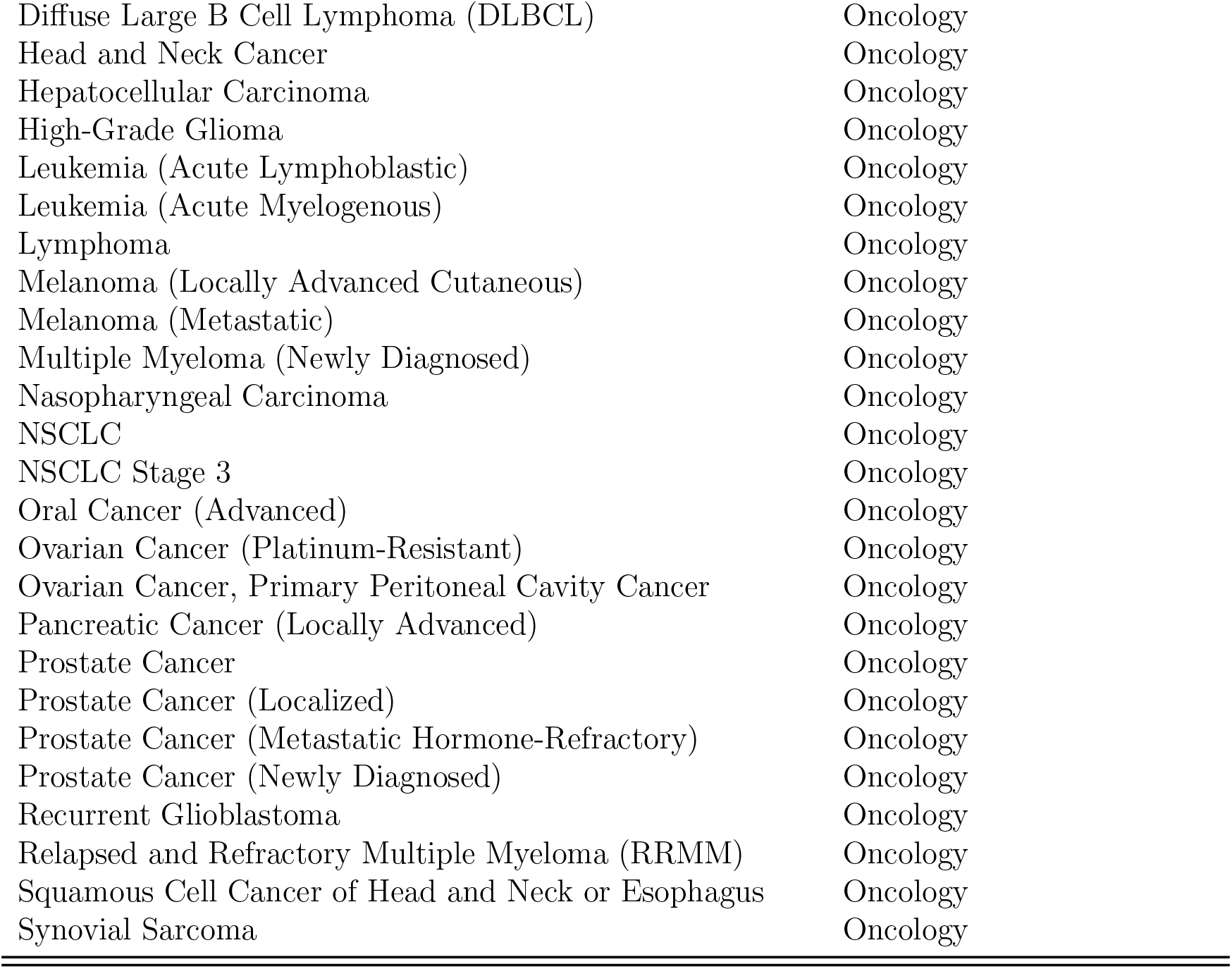
Diseases with ongoing gene therapy trials and their associated therapeutic areas.

## A3 Patient Population Estimation

We source the patient prevalence and incidence of the diseases from different sources. When necessary, we compute the prevalence from the incidence using Equation 1, or vice versa, using Equation 2. Our results are shown in Table A3. These numbers do not reflect the adjustments we make to NSC lung cancer, prostrate cancer and spinal muscular atrophy in order to minimize overlapping patient groups.

**Table A3:** Number of current patients and annual new patients for each disease. An asterisk (*) indicates that either the prevalence is computed from the incidence using Equation 1, or vice versa, using Equation 2.

| Disease | Current patients | New patients per year |
| --- | --- | --- |
| **– General Conditions –** |  |  |
| Arteriosclerosis Obliterans | [95]8500000 | *192100 |
| Critical Limb Ischemia | [85]975000 | [85]300000 |
| Degenerative Arthritis | [161]27000000 | *486000 |
| Diabetic Foot Ulcers | [130]2250000 | [129]112500 |
| Diabetic Peripheral Neuropathy | [69,96,168]9441480 | [69,96,168]467400 |
| Heart Failure | [107]5800000 | [107]812000 |
| Knee Osteoarthritis with Kellgren & Lawrence Grade 2 or/and 3 | *2929730 | [161]542000 |
| Parkinson’s Disease | [20]500000 | [20]50000 |
| Peripheral Artery Disease | [95]8500000 | *564400 |
| Refractory Angina due to Myocardial Ischemia (AF-FIRM) | [5]8200000 | [5,73]565000 |
| Stable Angina | [44]10000000 | [44]500000 |
| **– Rare Diseases –** |  |  |
| Beta-Thalassemia | [11]1000 | [137]3277 |
| Cerebral Adrenoleukodystrophy (CALD) | [66]411 | [66]37 |
| Choroideremia | [127]6554 | [127]77 |
| Cystic Fibrosis | [65]30000 | [65]1000 |
| Ewing’s Sarcoma | [13,14,61]15003 | [13,14,61]200 |
| Hemophilia A | [47]16000 | [47]360 |
| Hemophilia B | [47]4000 | [47]90 |
| Leber Congenital Amaurosis due to RPE65 Mutations | [79]187 | [79]19 |
| Leber Hereditary Optic Neuropathy | [27]6540 | [27]654 |
| Lipoprotein Lipase Deficiency (LPLD) | [75]328 | *33 |
| Metachromatic Leukodystrophy | [48,139]9333 | [48,139]771 |
| Mucopolysaccharidosis Type IIIa | [30]1638 | [30]39 |
| Recessive Dystrophic Epidermolysis Bullosa | [114]100 | *10 |

Table A3 – continued from previous page
| Disease | Current patients | New patients per year |
| --- | --- | --- |
| Retinitis Pigmentosa | [144]87387 | [144]8739 |
| Sickle Cell Anemia | [58]100000 | [59]58745 |
| Spinal Muscular Atrophy | [123]8526 | [123]290 |
| Spinal Muscular Atrophy Type 1 | [42]17500 | [82]500 |
| – **Cancer** – |  |  |
| B-Cell Non-Hodgkin’s Lymphoma | [9,10]694704 | [9,10]74200 |
| BCG Unresponsive NMIBC | [12,108,119,120]371933 | [12,108,119,120]42625 |
| Bladder Cancer, in situ concurrent with Papillary Tumors | [12]356720 | [12]41040 |
| Diffuse Large B Cell Lymphoma (DLBCL) | [1]257 | [1]18351 |
| Head and Neck Cancer | [50,53,54,55]134337 | [50,53,54,55]75275 |
| Hepatocellular Carcinoma | [15,103,105]11287 | [15,103,105]2032 |
| High-Grade Glioma | [3,8,49,51,147]87540 | [3,8,49,51,147]16334 |
| Leukemia (Acute Lymphoblastic) | [6]95764 | [6]5930 |
| Leukemia (Acute Myelogenous) | [7]61048 | [7]21450 |
| Lymphoma | [16,34]905678 | [16,34]82310 |
| Melanoma (Locally Advanced Cutaneous) | [17]107605 | [17]8683 |
| Melanoma (Metastatic) | [17]47824 | [17]3859 |
| Multiple Myeloma (Newly Diagnosed) | 0 | [2]32270 |
| Nasopharyngeal Carcinoma | [56]5390 | [56]327 |
| NSC Lung Cancer | [35,57]454469 | [57] 191646 |
| NSC Lung Cancer Stage 3 | [35,57,141]151490 | [57,141]63882 |
| Oral Cancer (Advanced) | [4]250000 | [4]53000 |
| Ovarian Cancer (Platinum-Resistant) | [36,37,38]141150 | [36,37,38]13956 |
| Ovarian Cancer, Primary Peritoneal Cavity Cancer | [40]2290 | [40]240 |
| Pancreatic Cancer (Locally Advanced) | [39,148]22066 | [39,148]17031 |
| Prostate Cancer | [18]3110403 | [18]174650 |
| Prostate Cancer (Localized) | [18]2395010 | [18]134481 |
| Prostate Cancer (Metastatic Hormone-Refractory) | [18]186624 | [18]10479 |
| Prostate Cancer (Newly Diagnosed) | 0 | [18]174650 |
| Recurrent Glioblastoma | [8,49,51,147]64127 | [8,49,51,147]12120 |
| Relapsed and Refractory Multiple Myeloma (RRMM) | [41,81]48840 | [41,81]16280 |
| Squamous Cell Cancer of Head and Neck or Esophagus | [50,53,54,55]120903 | [50,53,54,55]67747 |
| Synovial Sarcoma | [43,89]7282 | [89]655 |

## A4 Calibration of Survival Functions *D*_*alt*_(*x* − *a*)

We source either the survival or mortality rate from literature and use them to compute *λ*, the time parameter in the exponential survival function. We show our result in the table below.

**Table A4:** List of survival rate or mortality rate and *λ*, for each disease. An asterisk (*) under *λ* denotes that the disease does not affect mortality directly.

| Disease | $k$ years survival rate | | $k$ years mortality rate | | | $\lambda$ |
| --- | --- | --- | --- | --- | --- | --- |
| | $k = 5$ | $k = 10$ | $k = 1$ | $k = 5$ | $k = 10$ | |
| Arteriosclerosis Obliterans |  |  |  | [160]11.3 |  | 0.024 |
| Critical Limb Ischemia |  |  |  | [86]50 |  | 0.139 |
| Degenerative Arthritis |  | [163]82 |  |  |  | 0.020 |
| Diabetic Foot Ulcers |  |  |  | [146]49 |  | 0.135 |
| Diabetic Peripheral Neuropathy |  |  |  |  | [62]5 | 0.005 |
| Heart Failure |  |  |  | [52]42.3 |  | 0.110 |
| Knee Osteoarthritis with Kellgren & Lawrence Grade 2 | [155]7.5 |  |  |  |  | 0.518 |
| Knee Osteoarthritis with Kellgren & Lawrence Grade 3 | [155]7.5 |  |  |  |  | 0.518 |
| Parkinson's Disease | [165]40 |  |  |  |  | 0.174 |
| Peripheral Artery Disease |  |  |  | [76]33.2 |  | 0.081 |
| Refractory Angina due to Myocardial Ischemia (AF-FIRM) |  |  | [110]3.9 |  |  | 0.040 |
| Stable Angina | [111]90 |  |  |  |  | 0.021 |
| Beta-Thalassemia |  | [169]98.3 |  |  |  | 0.002 |
| Cerebral Adrenoleukodystrophy (CALD) | [143]55 |  |  |  |  | 0.120 |
| Choroideremia |  |  |  |  |  | * |
| Cystic Fibrosis |  |  |  |  | [121]28 | 0.033 |
| Ewing's Sarcoma | [131]70 |  |  |  |  | 0.071 |
| Hemophilia A |  |  |  |  | [140]9.7 | 0.010 |
| Hemophilia B |  |  |  |  | [140]9.7 | 0.010 |
| Leber Congenital Amaurosis due to RPE65 Mutations |  |  |  |  |  | * |

Table A4 – continued from previous page
| Disease | $k$ years survival rate | | $k$ years mortality rate | | | $\lambda$ |
| --- | --- | --- | --- | --- | --- | --- |
| | $k = 5$ | $k = 10$ | $k = 1$ | $k = 5$ | $k = 10$ | |
| Leber Hereditary Optic Neuropathy |  |  |  |  |  | * |
| Lipoprotein Lipase Deficiency (LPLD) |  |  |  |  |  | * |
| Metachromatic Leukodystrophy | [128]52 |  |  |  |  | 0.131 |
| Mucopolysaccharidosis Type III |  | [125]60 |  |  |  | 0.051 |
| Recessive Dystrophic Epidermolysis Bullosa |  |  |  |  |  | * |
| Retinitis Pigmentosa |  |  |  |  |  | * |
| Sickle Cell Anemia |  | [101]96 |  |  |  | 0.004 |
| Spinal Muscular Atrophy | [83]40 |  |  |  |  | 0.183 |
| Spinal Muscular Atrophy Type 1 | [170]10.13 |  |  |  |  | 0.458 |
| B-Cell Non-Hodgkin's Lymphoma | [31]72 |  |  |  |  | 0.066 |
| BCG Unresponsive Non-Muscle Invasive Bladder Cancer | [120]78 |  |  |  |  | 0.050 |
| Bladder Cancer, Transitional Cell Carcinoma | [21]95.8 |  |  |  |  | 0.009 |
| Diffuse Large B-Cell Lymphoma (DLBCL) | [26]63.2 |  |  |  |  | 0.092 |
| Head and Neck Cancer | [22]64 |  |  |  |  | 0.089 |
| Hepatocellular Carcinoma | [105]10 |  |  |  |  | 0.461 |
| High-Grade Glioma | [113]9.87 |  |  |  |  | 0.463 |
| Leukemia (Acute Lymphoblastic) | [6]68.8 |  |  |  |  | 0.075 |
| Leukemia (Acute Myelogenous) | [7]28.7 |  |  |  |  | 0.250 |
| Lymphoma | [29]72 |  |  |  |  | 0.066 |
| Melanoma (Locally Advanced Cutaneous) | [28]64 |  |  |  |  | 0.089 |
| Melanoma (Metastatic) | [28]23 |  |  |  |  | 0.294 |
| Multiple Myeloma (Newly Diagnosed) | [60]52 |  |  |  |  | 0.131 |
| Nasopharyngeal Carcinoma | [32]72 |  |  |  |  | 0.066 |
| NSCLC | [45]23 |  |  |  |  | 0.294 |
| NSCLC (Stage 3) | [45]33 |  |  |  |  | 0.222 |
| Oral Cancer (Advanced) | [22]39.1 |  |  |  |  | 0.188 |

Table A4 – continued from previous page
| Disease | $k$ years survival rate | | $k$ years mortality rate | | | $\lambda$ |
| --- | --- | --- | --- | --- | --- | --- |
| | $k = 5$ | $k = 10$ | $k = 1$ | $k = 5$ | $k = 10$ | |
| Ovarian Cancer (Platinum-Resistant) | [162] 1.9 |  |  |  |  | 0.793 |
| Ovarian Cancer, Primary Peritoneal Cavity Cancer | [23] 47.6 |  |  |  |  | 0.148 |
| Pancreatic Cancer (Locally Advanced) | [24] 12.4 |  |  |  |  | 0.417 |
| Prostate Cancer | [25] 98 |  |  |  |  | 0.004 |
| Prostate Cancer (Localized) | [25] 98 |  |  |  |  | 0.004 |
| Prostate Cancer (Metastatic Hormone Refractory) | [25] 30.5 |  |  |  |  | 0.237 |
| Prostate Cancer (Newly Diagnosed) | [25] 95.1 |  |  |  |  | 0.010 |
| Recurrent Glioblastoma | [?] 10 |  |  |  |  | 0.461 |
| Relapsed and Refractory Multiple Myeloma (RRMM) | [46] 9.92 |  |  |  |  | 0.462 |
| Squamous Cell Cancer of Head and Neck or Esophagus | [22] 64 |  |  |  |  | 0.089 |
| Synovial Sarcoma | [33] 55 |  |  |  |  | 0.120 |

## A5 Calibration of Age Distribution *A*(*x*)

As mentioned in the main paper, our optimization program produces triangular age distributions that conforms to data, have wider support compared to fitting uniform distributions and, avoids sharp changes in the probability density. We illustrate some examples that compare triangle distributions with the uniform distributions with the same average age.

**Figure A1:**
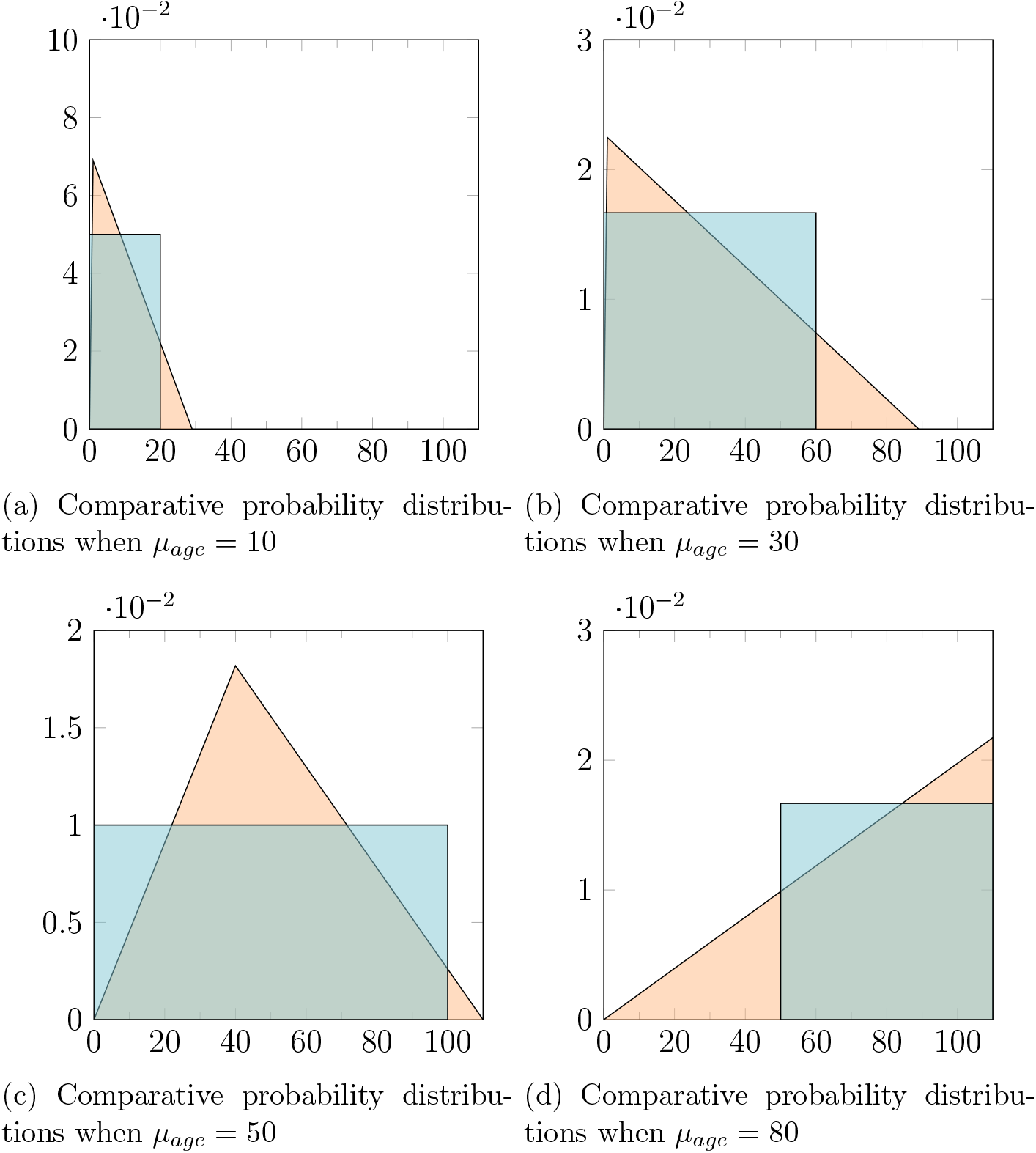
Age distributions given various mean ages, *μ*_*age*_. The red triangles represent the solutions obtained by our optimization program, while the blue rectangles represent the solutions given by an uniform distribution. The distributions from the optimization program have a wider base of support and avoid sharp changes in density.

## A6 Quality of Life Estimation

The results of our literature search and estimation for the change in QoL for each disease is shown in the table below.

**Table A5:** Table of disease scores (*ζ*), estimated quality of life values before treatment 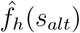, after treatment 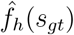, and the change in quality of life (ΔQoL). Asterisks (*) indicate that the values are interpolated. Cancers are not included, as we assume that the gains in survival dominate the gains in QoL.

| Non-Cancer Disease | $\zeta$ | $\hat{f}_h(s_{alt})$ | $\Delta\text{QoL}$ | $\hat{f}_h(s_{gt})$ |
| --- | --- | --- | --- | --- |
| Arteriosclerosis Obliterans | 1 | *0.775 | *0.075 | *0.850 |
| Beta-Thalassemia | 3 | <sup>[151]</sup>0.870 | *0.166 | *1.000 |
| Cerebral Adrenoleukodystrophy (CALD) | 5 | *0.654 | *0.257 | *0.911 |
| Choroideremia | 3 | *0.715 | *0.166 | *0.881 |
| Critical Limb Ischemia | 4 | *0.684 | *0.212 | *0.896 |
| Cystic Fibrosis | 3 | <sup>[64]</sup>0.671 | *0.166 | *0.837 |
| Degenerative Arthritis | 3 | *0.715 | *0.166 | *0.881 |
| Diabetic Foot Ulcers | 3 | <sup>[154]</sup>0.703 | <sup>[154]</sup>0.258 | <sup>[154]</sup>0.961 |
| Diabetic Peripheral Neuropathy | 2 | <sup>[153]</sup>0.630 | <sup>[153]</sup>0.180 | <sup>[153]</sup>0.810 |
| Ewing's Sarcoma | 2 | <sup>[145]</sup>0.690 | *0.121 | *0.811 |
| Heart Failure | 4 | *0.684 | *0.212 | *0.896 |
| Hemophilia A | 5 | <sup>[77]</sup>0.750 | *0.257 | *1.000 |
| Hemophilia B | 5 | <sup>[77]</sup>0.700 | *0.257 | *0.957 |
| Knee Osteoarthritis, Kellgren & Lawrence Grade 2 | 2 | <sup>[134]</sup>0.900 | <sup>[134]</sup>0.042 | *0.942 |
| Knee Osteoarthritis, Kellgren & Lawrence Grade 3 | 2 | <sup>[134]</sup>0.900 | <sup>[134]</sup>0.048 | *0.948 |
| Leber Congenital Amaurosis (RPE65 Mutations) | 3 | *0.715 | *0.166 | *0.881 |
| Leber Hereditary Optic Neuropathy | 3 | *0.715 | *0.166 | *0.881 |
| Lipoprotein Lipase Deficiency | 4 | *0.684 | *0.212 | *0.896 |
| Lysosomal Storage Disease | 5 | <sup>[100]</sup>0.640 | *0.257 | *0.897 |
| Mucopolysaccharidosis Type IIIa | 2 | <sup>[109]</sup>0.582 | <sup>[109]</sup>0.264 | <sup>[109]</sup>0.846 |
| Osteoarthritis | 2 | <sup>[158]</sup>0.900 | <sup>[158]</sup>0.040 | *0.940 |
| Parkinson's Disease | 4 | <sup>[87,152]</sup>0.700 | <sup>[87]</sup>0.150 | <sup>[87]</sup>0.850 |
| Peripheral Artery Disease | 2 | <sup>[112]</sup>0.660 | <sup>[112]</sup>0.060 | <sup>[112]</sup>0.720 |
| Recessive Dystrophic Epidermolysis Bullosa | 4 | <sup>[118]</sup>0.590 | *0.212 | *0.802 |
| Refractory Angina due to Myocardial Ischemia | 2 | <sup>[106]</sup>0.600 | *0.121 | *0.721 |
| Retinitis Pigmentosa | 3 | <sup>[150]</sup>0.770 | *0.166 | *0.936 |
| Sickle Cell Anemia | 3 | <sup>[94]</sup>0.732 | <sup>[94]</sup>0.198 | <sup>[94]</sup>0.930 |
| Spinal Muscular Atrophy<sup>6</sup> | 5 | 0.520 | *0.257 | *0.777 |
| Spinal Muscular Atrophy Type 1 | 5 | <sup>[172]</sup>0.520 | *0.257 | *0.777 |
| Stable Angina | 2 | <sup>[122,167]</sup>0.750 | <sup>[167]</sup>0.150 | <sup>[167]</sup>0.900 |

<sup>6</sup>We are unable to find QoL values for SMA only and assume that they are the same as SMA Type 1.

## A7 Simulation Convergence Criteria

Let *X*_*k*_ be the results of the *k*-th simulation. *X*_*k*_ has a true mean of *μ* and variance *σ*^2^. Let the mean of the Monte Carlo simulations over *n* runs be 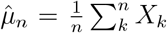. Then, by Linde-berg–Lévy’s Central Limit Theorem, 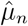 converges in distribution to a normal distribution with mean *μ* and variance of *nσ*^2^. The 95 percent confidence interval for *μ* is given by:

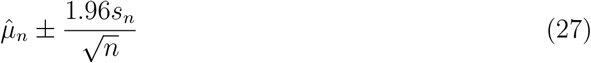

where *s*_*n*_ is the sample variance of {*X*_1_,, *X*_*n*_}.

Since we are using 1-by-T vectors, we investigated the error in our simulation by dividing the half-range of the confidence interval in each time-step by 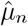 before taking the maximum across the time series. As can be seen from Figure A2, we should expect the simulated mean to be within 1.89% of the true mean 95% of the time with 1,000,000 iterations.

**Figure A2:**
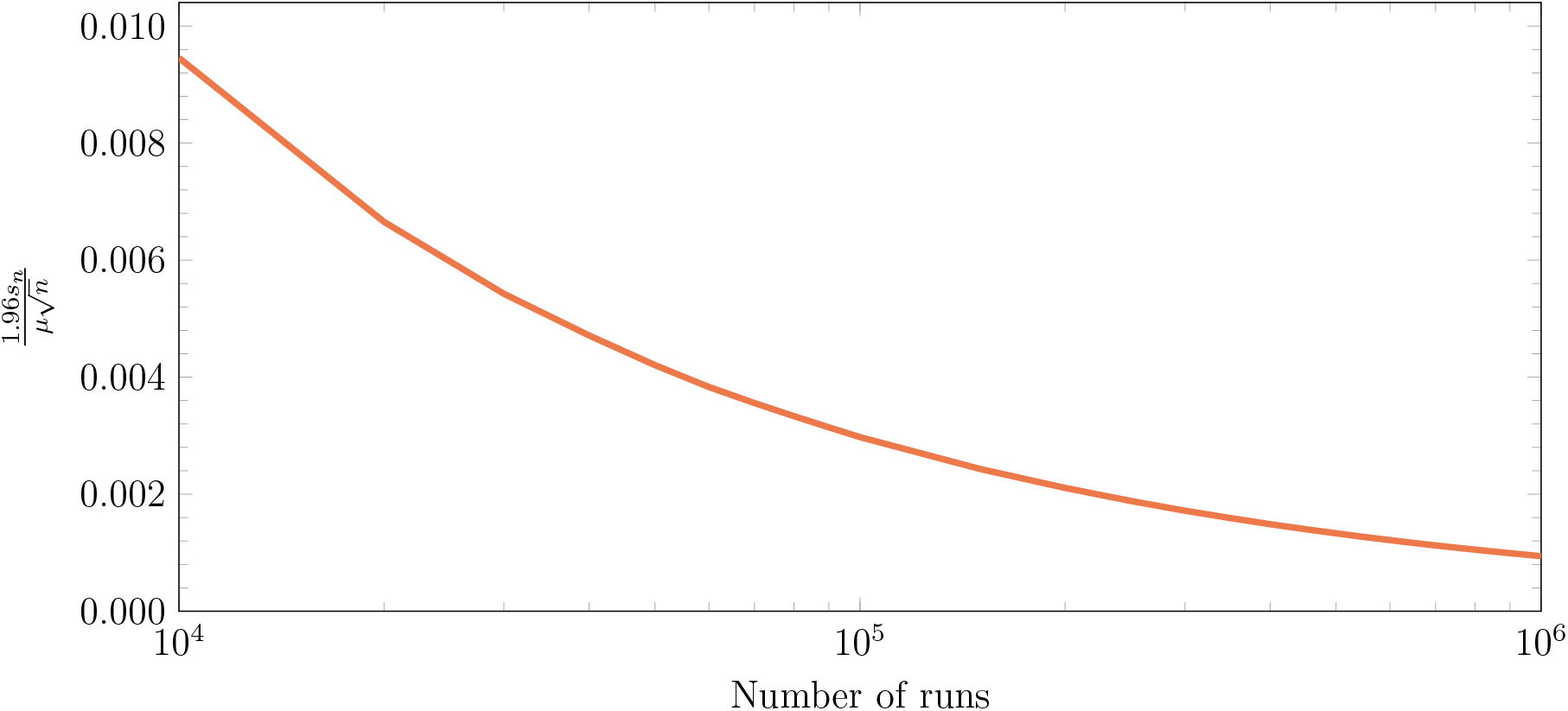
Plot of ^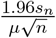^ against the number of iterations of simulations of the cost.

## A8 Pseudo-Code and Implementation Details

### Pseudo-code

We perform a Monte Carlo simulation to determine the total number of patients undergoing gene therapy and the cost of these gene therapies at specific points in time. The sequence of computations for each iteration of the simulation is detailed in Algorithm 1.

#### Algorithm 1: Pseudocode for one iteration of the simulation.

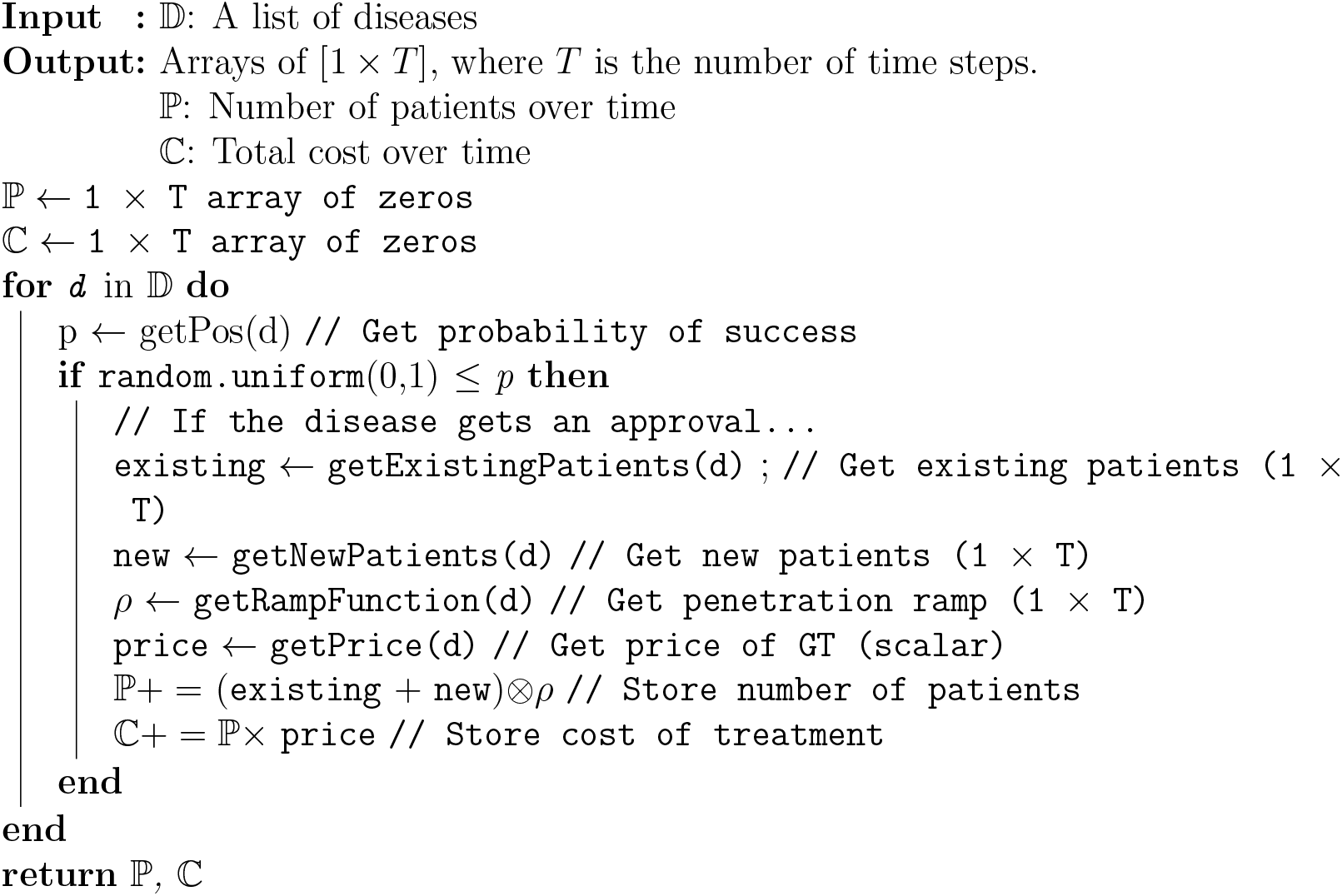

### Implementation

All the equations are discretized for computation from their continuous forms. When solving the integrals using the trapezoidal rule to obtain ΔQALY, we use strip widths of 1 year across a range from 0 to 110 years old, the resolution offered by the life tables. When simulating the number of patients and the cost over time, we use steps of 1 month.

Our codes are implemented on Python 3.6 backed by Numpy. Our vectorized implementation averages 6.120ms per iteration over 1,000,000 runs on a single thread of an Intel Xeon Gold 5120, clocked at 2.20GHz with 20GB of RAM. We attempted to use PyTorch to speed up the computations using a GPU, but it ran more slowly than a single-threaded CPU. We determined this took place for two reasons. First, generating random numbers must be sequential, since PyTorch delegates it solely to the CPU, which limits the amount of parallelization that can be achieved, as dictated by Amdahl’s law. Second, because our computations require a large amount of data from different sources, they must be batched due to the GPU’s limited RAM. The constant movement of data through the PCIe bridge, however, turns out to be a massive bottleneck to the overall speed.

## A9 Visualization of the Cost over Time

In this section, we visualize how the monthly cost of treating patients with gene therapy will be affected by changes to the variables. The results are summarized in the tornado chart presented in the main paper.

**Figure A3:**
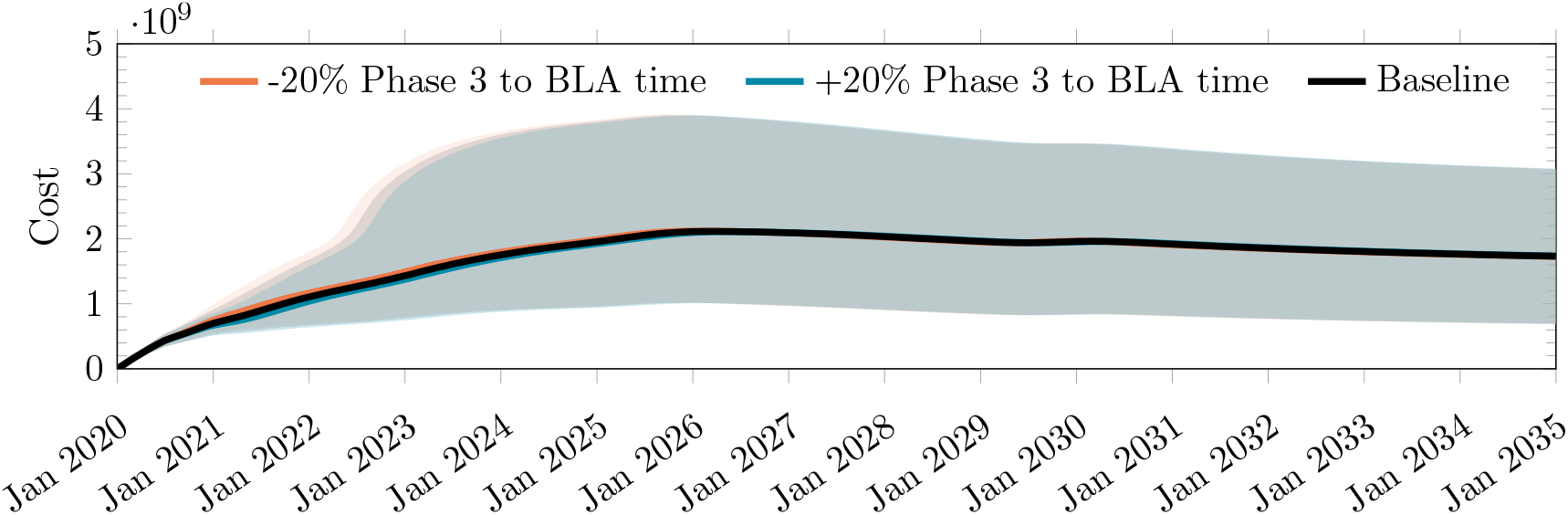
Impact on monthly cost of treating patients given a ±20% change in the time from phase 3 to BLA.

**Figure A4:**
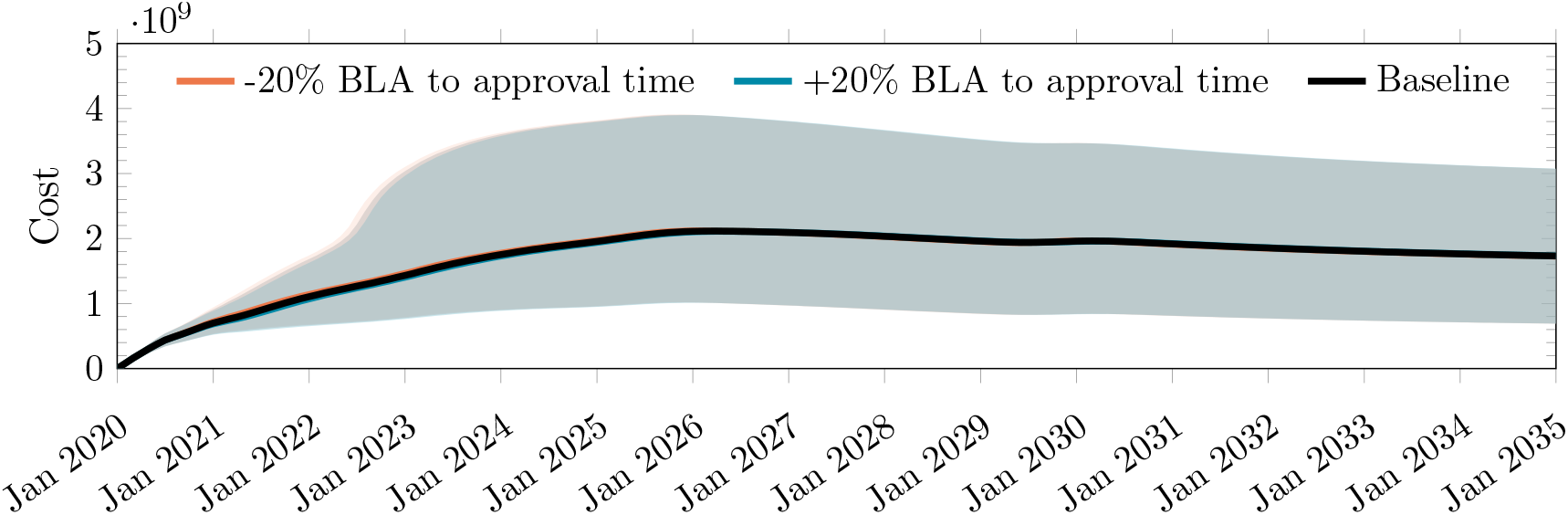
Impact on monthly cost of treating patients given a ±20% change in the time from BLA to approval.

**Figure A5:**
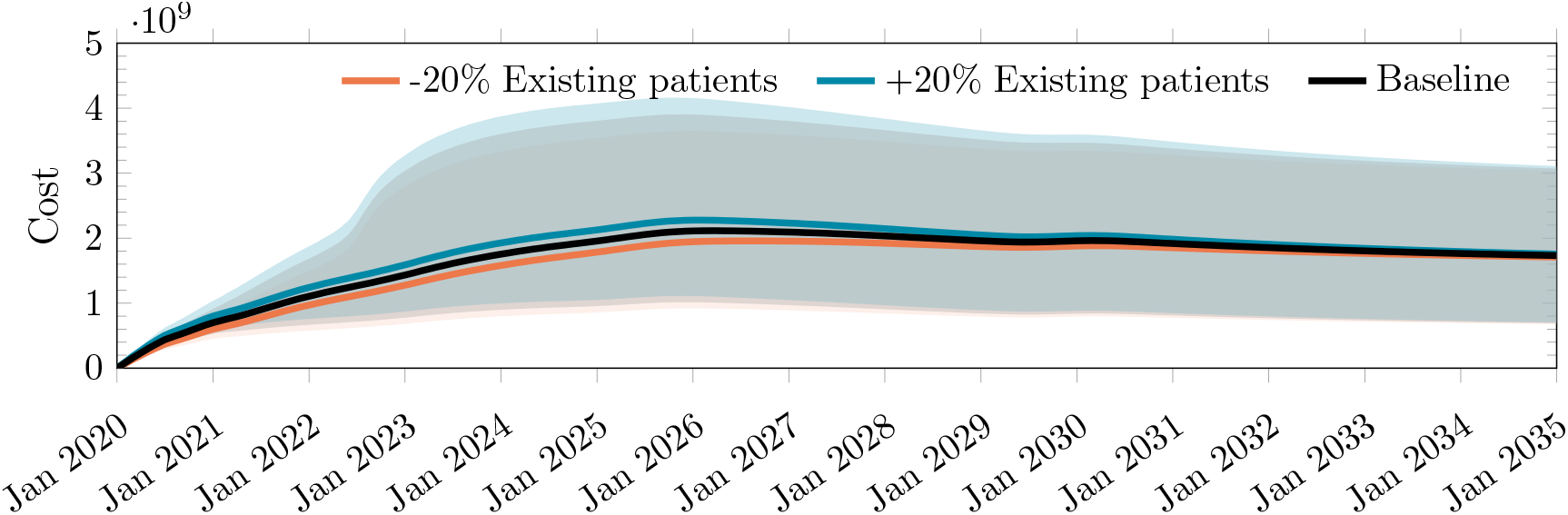
Impact on monthly cost of treating patients given a±20% change in the number of existing patients.

**Figure A6:**
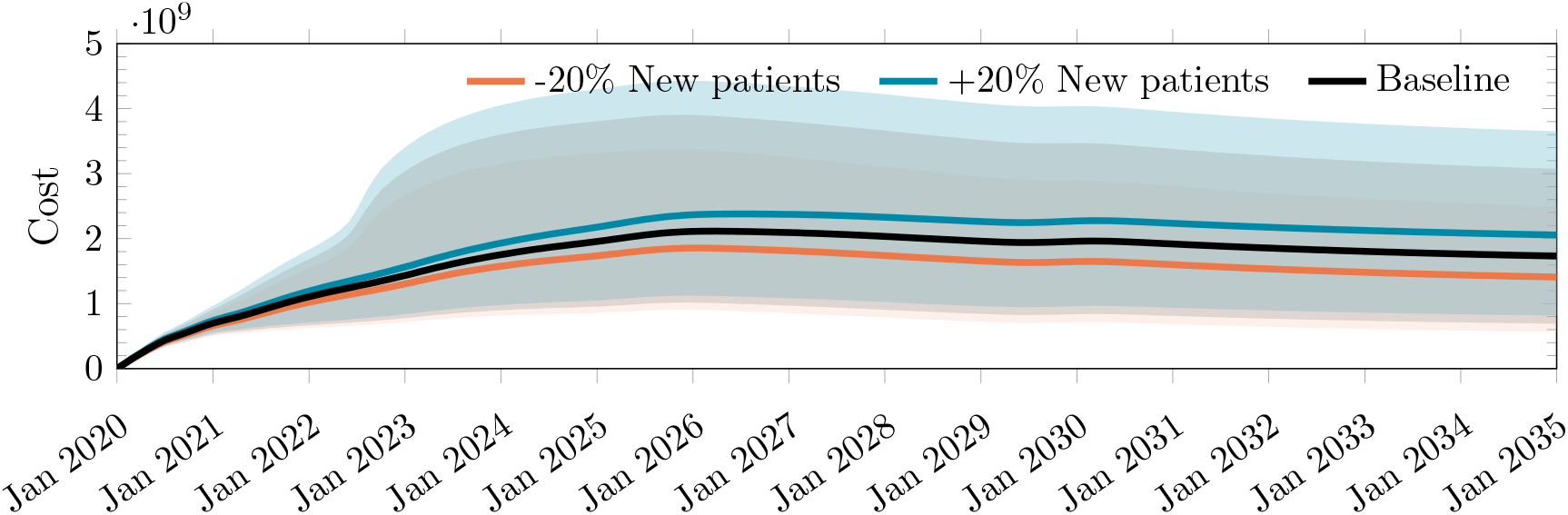
Impact on monthly cost of treating patients given a ±20% change in the number of new patients.

**Figure A7:**
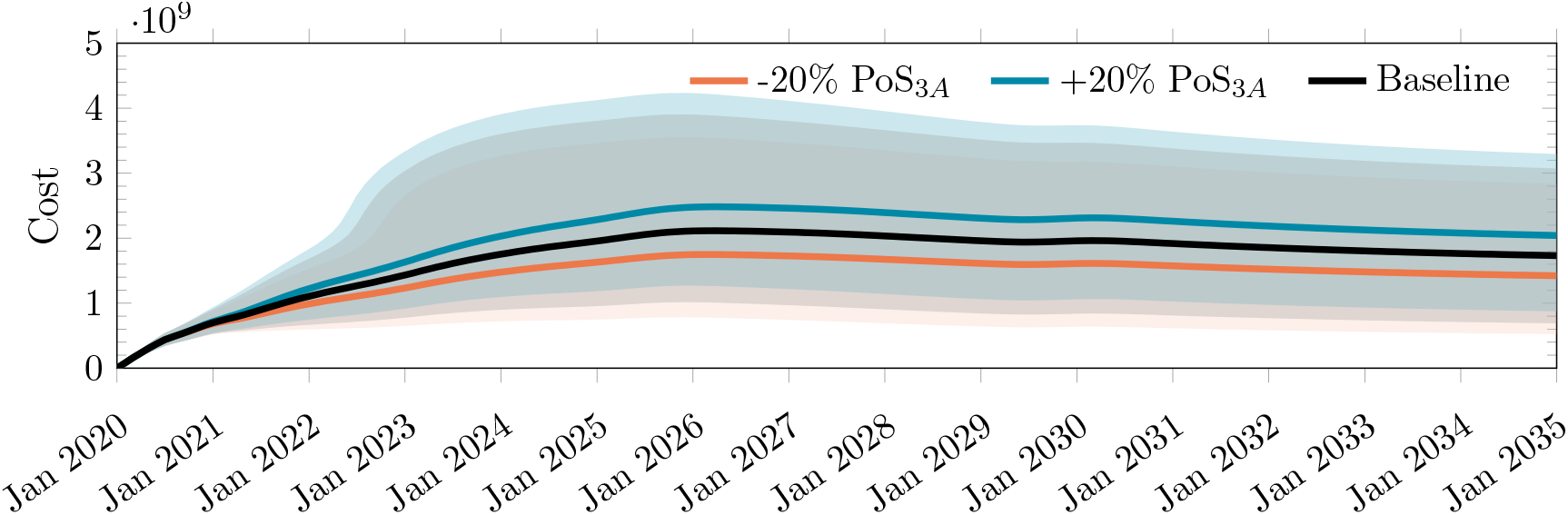
Impact on monthly cost of treating patients given a ±20% change in the PoS_3*A*_.

**Figure A8:**
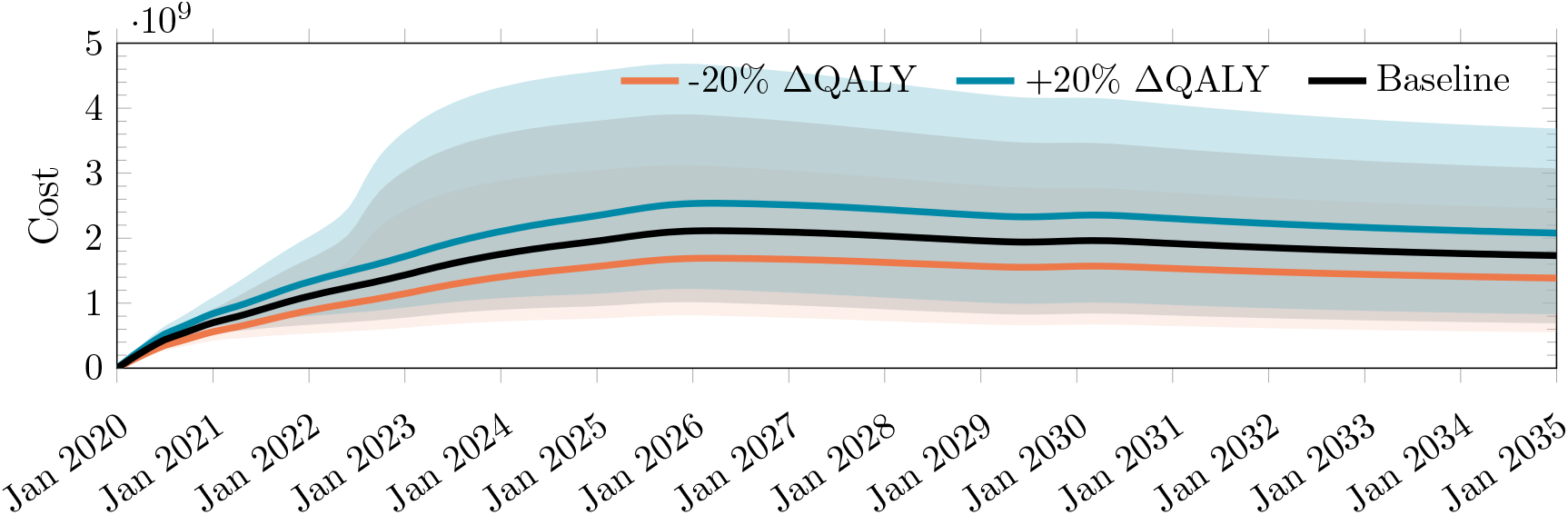
Impact on monthly cost of treating patients given a ±20% change in ΔQALY gained.

**Figure A9:**
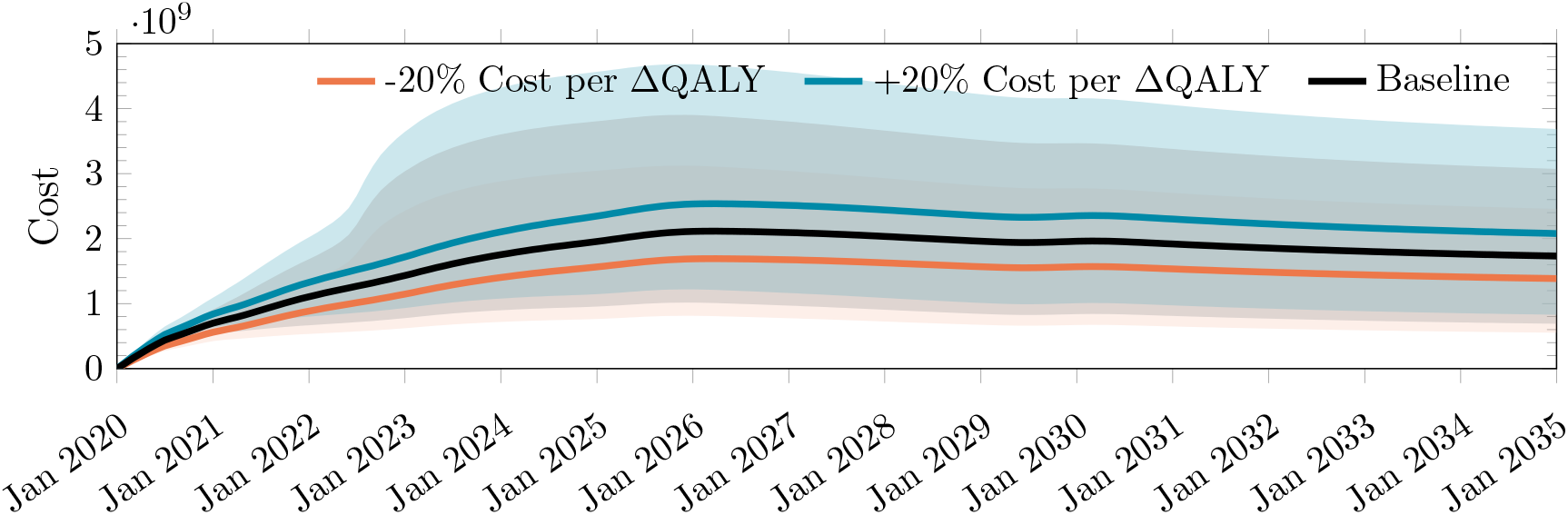
Impact on monthly cost of treating patients given a ±20% change in the cost per ΔQALY.

**Figure A10:**
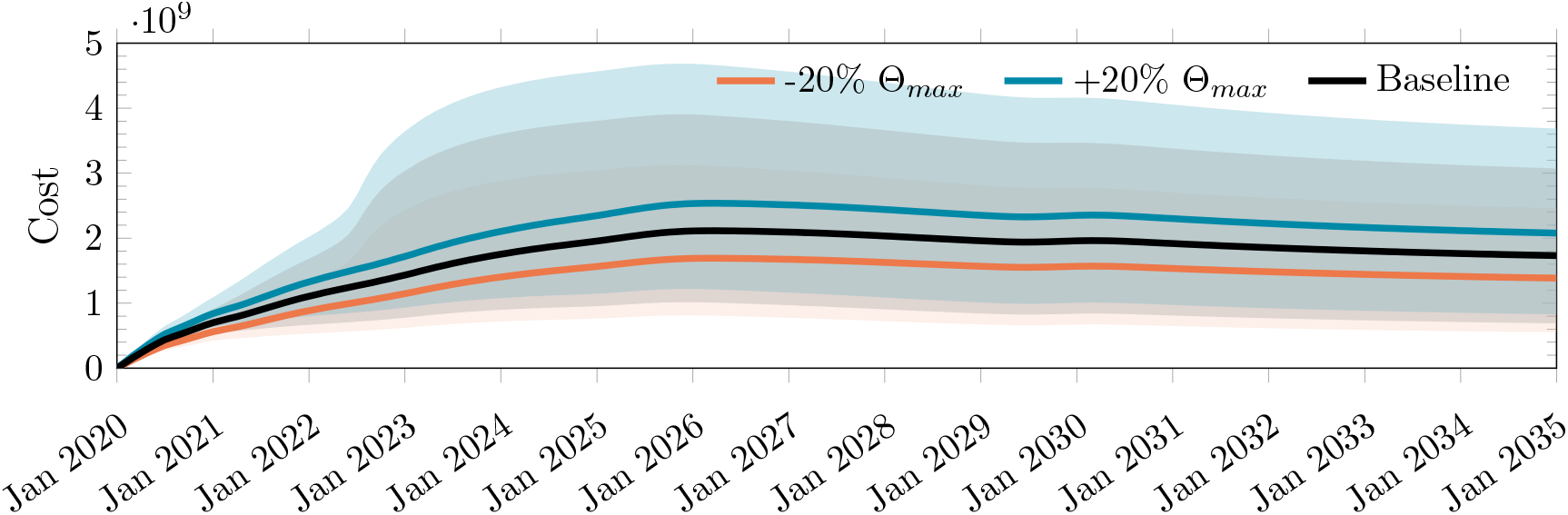
Impact on monthly cost of treating patients given a ±20% change in Θ_*max*_.

**Figure A11:**
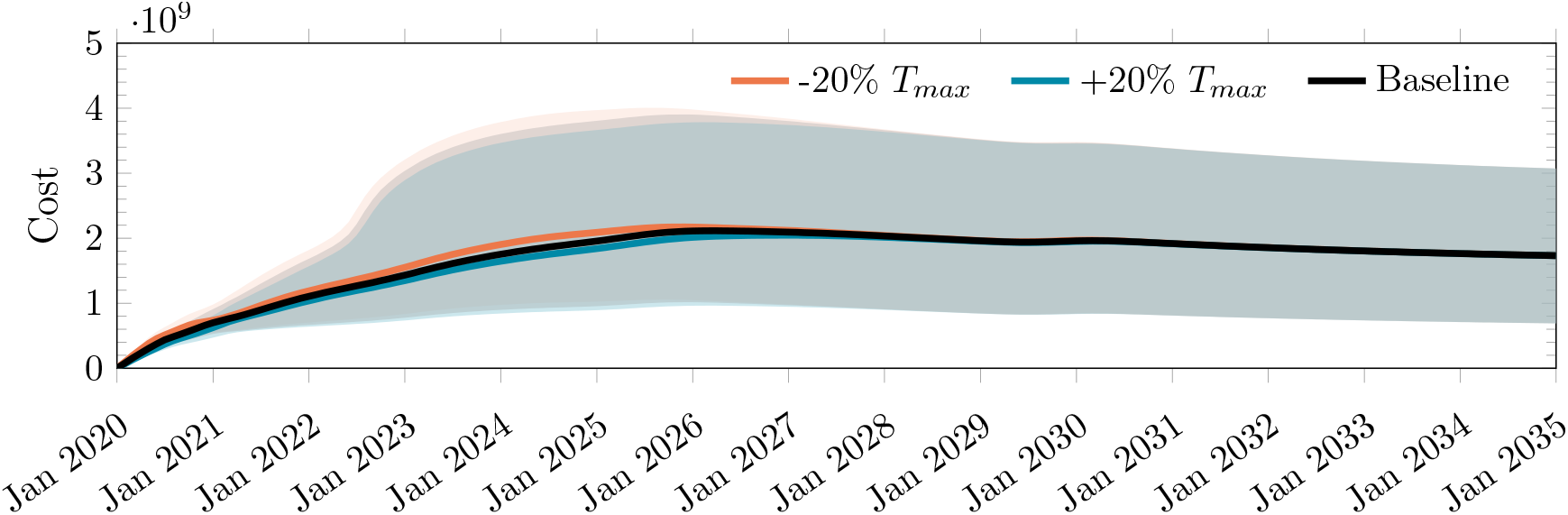
Impact on monthly cost of treating patients given a ±20% change in *T*_*max*_.

## Footnotes

1 Compassionate use, also known as ‘expanded access’, refers to the administration of investigational treatments outside of the clinical trial to treat patients with serious or immediately life-threatening diseases, or conditions when there are no comparable or satisfactory alternative treatment options.

2 We classify Ewing’s Sarcoma—a rare form of cancer—as a rare disease instead of cancer.

3 It must be noted that several pathways, such as the priority review program, are exceptions to this rule. The assumption of perfect correlation still holds if only one trial is required for regulatory review.

4 ICER provides a range of ΔQALY estimates corresponding to different age groups. We have considered the distribution of ages to produce a weighted average estimate.

5 The spending estimates for Medicaid do not take into account the 23.1% drug rebate that it is expected to receive [98].

6 We are unable to find QoL values for SMA only and assume that they are the same as SMA Type 1.

